# Genome-wide association study of over 40,000 bipolar disorder cases provides new insights into the underlying biology

**DOI:** 10.1101/2020.09.17.20187054

**Authors:** Niamh Mullins, Andreas J. Forstner, Kevin S. O’Connell, Brandon Coombes, Jonathan R. I. Coleman, Zhen Qiao, Thomas D. Als, Tim B. Bigdeli, Sigrid Børte, Julien Bryois, Alexander W. Charney, Ole Kristian Drange, Michael J. Gandal, Saskia P. Hagenaars, Masashi Ikeda, Nolan Kamitaki, Minsoo Kim, Kristi Krebs, Georgia Panagiotaropoulou, Brian M. Schilder, Laura G. Sloofman, Stacy Steinberg, Vassily Trubetskoy, Bendik S. Winsvold, Hong-Hee Won, Liliya Abramova, Kristina Adorjan, Esben Agerbo, Mariam Al Eissa, Diego Albani, Ney Alliey-Rodriguez, Adebayo Anjorin, Verneri Antilla, Anastasia Antoniou, Swapnil Awasthi, Ji Hyun Baek, Marie Bækvad-Hansen, Nicholas Bass, Michael Bauer, Eva C. Beins, Sarah E. Bergen, Armin Birner, Carsten Bøcker Pedersen, Erlend Bøen, Marco P. Boks, Rosa Bosch, Murielle Brum, Ben M. Brumpton, Nathalie Brunkhorst-Kanaan, Monika Budde, Jonas Bybjerg-Grauholm, William Byerley, Murray Cairns, Miquel Casas, Pablo Cervantes, Toni-Kim Clarke, Cristiana Cruceanu, Alfredo Cuellar-Barboza, Julie Cunningham, David Curtis, Piotr M. Czerski, Anders M. Dale, Nina Dalkner, Friederike S. David, Franziska Degenhardt, Srdjan Djurovic, Amanda L. Dobbyn, Athanassios Douzenis, Torbjørn Elvsåshagen, Valentina Escott-Price, I. Nicol Ferrier, Alessia Fiorentino, Tatiana M. Foroud, Liz Forty, Josef Frank, Oleksandr Frei, Nelson B. Freimer, Louise Frisén, Katrin Gade, Julie Garnham, Joel Gelernter, Marianne Giørtz Pedersen, Ian R. Gizer, Scott D. Gordon, Katherine Gordon-Smith, Tiffany A. Greenwood, Jakob Grove, José Guzman-Parra, Kyooseob Ha, Magnus Haraldsson, Martin Hautzinger, Urs Heilbronner, Dennis Hellgren, Stefan Herms, Per Hoffmann, Peter A. Holmans, Laura Huckins, Stéphane Jamain, Jessica S. Johnson, Janos L. Kalman, Yoichiro Kamatani, James L. Kennedy, Sarah Kittel-Schneider, James A. Knowles, Manolis Kogevinas, Maria Koromina, Thorsten M. Kranz, Henry R. Kranzler, Michiaki Kubo, Ralph Kupka, Steven A. Kushner, Catharina Lavebratt, Jacob Lawrence, Markus Leber, Heon-Jeong Lee, Phil H. Lee, Shawn E. Levy, Catrin Lewis, Calwing Liao, Susanne Lucae, Martin Lundberg, Donald J. MacIntyre, Sigurdur H. Magnusson, Wolfgang Maier, Adam Maihofer, Dolores Malaspina, Eirini Maratou, Lina Martinsson, Manuel Mattheisen, Steven A. McCarroll, Nathaniel W. McGregor, Peter McGuffin, James D. McKay, Helena Medeiros, Sarah E. Medland, Vincent Millischer, Grant W. Montgomery, Jennifer L. Moran, Derek W. Morris, Thomas W. Mühleisen, Niamh O’Brien, Claire O’Donovan, Loes M. Olde Loohuis, Lilijana Oruc, Sergi Papiol, Antonio F. Pardiñas, Amy Perry, Andrea Pfennig, Evgenia Porichi, James B. Potash, Digby Quested, Towfique Raj, Mark H. Rapaport, J. Raymond DePaulo, Eline J. Regeer, John P. Rice, Fabio Rivas, Margarita Rivera, Julian Roth, Panos Roussos, Douglas M. Ruderfer, Cristina Sánchez-Mora, Eva C. Schulte, Fanny Senner, Sally Sharp, Paul D. Shilling, Engilbert Sigurdsson, Lea Sirignano, Claire Slaney, Olav B. Smeland, Daniel J. Smith, Janet L. Sobell, Christine Søholm Hansen, Maria Soler Artigas, Anne T. Spijker, Dan J. Stein, John S. Strauss, Beata Świątkowska, Chikashi Terao, Thorgeir E. Thorgeirsson, Claudio Toma, Paul Tooney, Evangelia-Eirini Tsermpini, Marquis P. Vawter, Helmut Vedder, James T. R. Walters, Stephanie H. Witt, Simon Xi, Wei Xu, Jessica Mei Kay Yang, Allan H. Young, Hannah Young, Peter P. Zandi, Hang Zhou, Lea Zillich, HUNT All-In Psychiatry, Rolf Adolfsson, Ingrid Agartz, Martin Alda, Lars Alfredsson, Gulja Babadjanova, Lena Backlund, Bernhard T. Baune, Frank Bellivier, Susanne Bengesser, Wade H. Berrettini, Douglas H. R. Blackwood, Michael Boehnke, Anders D. Børglum, Gerome Breen, Vaughan J. Carr, Stanley Catts, Aiden Corvin, Nicholas Craddock, Udo Dannlowski, Dimitris Dikeos, Tõnu Esko, Bruno Etain, Panagiotis Ferentinos, Mark Frye, Janice M. Fullerton, Micha Gawlik, Elliot S. Gershon, Fernando S. Goes, Melissa J. Green, Maria Grigoroiu-Serbanescu, Joanna Hauser, Frans Henskens, Jan Hillert, Kyung Sue Hong, David M. Hougaard, Christina M. Hultman, Kristian Hveem, Nakao Iwata, Assen V. Jablensky, Ian Jones, Lisa A. Jones, René S. Kahn, John R. Kelsoe, George Kirov, Mikael Landén, Marion Leboyer, Cathryn M. Lewis, Qingqin S. Li, Jolanta Lissowska, Christine Lochner, Carmel Loughland, Nicholas G. Martin, Carol A. Mathews, Fermin Mayoral, Susan L. McElroy, Andrew M. McIntosh, Francis J. McMahon, Ingrid Melle, Patricia Michie, Lili Milani, Philip B. Mitchell, Gunnar Morken, Ole Mors, Preben Bo Mortensen, Bryan Mowry, Bertram Müller-Myhsok, Richard M. Myers, Benjamin M. Neale, Caroline M. Nievergelt, Merete Nordentoft, Markus M. Nöthen, Michael C. O’Donovan, Ketil J. Oedegaard, Tomas Olsson, Michael J. Owen, Sara A. Paciga, Chris Pantelis, Carlos Pato, Michele T. Pato, George P. Patrinos, Roy H. Perlis, Danielle Posthuma, Josep Antoni Ramos-Quiroga, Andreas Reif, Eva Z. Reininghaus, Marta Ribasés, Marcella Rietschel, Stephan Ripke, Guy A. Rouleau, Takeo Saito, Ulrich Schall, Martin Schalling, Peter R. Schofield, Thomas G. Schulze, Laura J. Scott, Rodney J. Scott, Alessandro Serretti, Cynthia Shannon Weickert, Jordan W. Smoller, Hreinn Stefansson, Kari Stefansson, Eystein Stordal, Fabian Streit, Patrick F. Sullivan, Gustavo Turecki, Arne E. Vaaler, Eduard Vieta, John B. Vincent, Irwin D. Waldman, Thomas W. Weickert, Thomas Werge, Naomi R. Wray, John-Anker Zwart, Joanna M. Biernacka, John I. Nurnberger, Sven Cichon, Howard J. Edenberg, Eli A. Stahl, Andrew McQuillin, Arianna Di Florio, Roel A. Ophoff, Ole A. Andreassen

## Abstract

Bipolar disorder (BD) is a heritable mental illness with complex etiology. We performed a genome-wide association study (GWAS) of 41,917 BD cases and 371,549 controls of European ancestry, which identified 64 associated genomic loci. BD risk alleles were enriched in genes in synaptic signaling pathways and brain-expressed genes, particularly those with high specificity of expression in neurons of the prefrontal cortex and hippocampus. Significant signal enrichment was found in genes encoding targets of antipsychotics, calcium channel blockers, antiepileptics and anesthetics. Integrating eQTL data implicated 15 genes robustly linked to BD via gene expression, encoding druggable targets such as HTR6, MCHR1, DCLK3 and FURIN. Analyses of BD subtypes indicated high but imperfect genetic correlation between BD type I and II and identified additional associated loci. Together, these results advance our understanding of the biological etiology of BD, identify novel therapeutic leads and prioritize genes for functional follow-up studies.

## Introduction

Bipolar disorder (BD) is a complex mental disorder characterized by recurrent episodes of (hypo)mania and depression. It is a common condition affecting an estimated 40 to 50 million people worldwide^1^. This, combined with the typical onset in young adulthood, an often chronic course, and increased risk of suicide^2^, make BD a major public health concern and a major cause of global disability^1^. Clinically, BD is classified into two main subtypes: bipolar I disorder, in which manic episodes typically alternate with depressive episodes, and bipolar II disorder, characterized by the occurrence of at least one hypomanic and one depressive episode^3^. These subtypes have a lifetime prevalence of ∼1% each in the population^4,5^.

Family and molecular genetic studies provide convincing evidence that BD is a multifactorial disorder, with genetic and environmental factors contributing to its development^6^. On the basis of twin and family studies, the heritability of BD is estimated at 60-85%^7,8^. Genome-wide association studies (GWAS)^9–23^ have led to valuable insights into the genetic etiology of BD. The largest such study has been conducted by the Psychiatric Genomics Consortium (PGC), in which genome-wide SNP data from 29,764 BD patients and 169,118 controls were analyzed and 30 genome-wide significant loci were identified (PGC2)^24^. SNP-based heritability (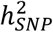) estimation using the same data, suggested that common genetic variants genome-wide explain ∼20% of BD’s phenotypic variance^24^. Polygenic risk scores generated from the results of this study explained ∼4% of phenotypic variance in independent samples. Across the genome, genetic associations with BD converged on specific biological pathways including regulation of insulin secretion^25,26^, retrograde endocannabinoid signaling^24^, glutamate receptor signaling^27^ and calcium channel activity^9^.

Despite this considerable progress, only a fraction of the genetic etiology of BD has been identified and the specific biological mechanisms underlying the development of the disorder are still unknown. In the present study, we report the results of the third GWAS meta-analysis of the PGC Bipolar Disorder Working Group, comprising 41,917 patients with BD and 371,549 controls. These results confirm and expand on many previously reported findings, identify novel therapeutic leads and prioritize genes for functional follow-up studies^28,29^. Thus, our results further illuminate the biological etiology of BD.

## Results

### GWAS results

A GWAS meta-analysis was conducted of 57 BD cohorts collected in Europe, North America and Australia (Table S1), totaling 41,917 BD cases and 371,549 controls of European descent (Effective N = 101,962, see online methods). For 52 cohorts, individual-level genotype and phenotype data were shared with the PGC and cases met international consensus criteria (DSM-IV, ICD-9 or ICD-10) for lifetime BD, established using structured diagnostic interviews, clinician-administered checklists or medical record review. BD GWAS summary statistics were received for five external cohorts (iPSYCH^30^, deCODE genetics^31^, Estonian Biobank^32^, Trøndelag Health Study (HUNT)^33^ and UK Biobank^34^), in which most cases were ascertained using ICD codes. The GWAS meta-analysis identified 64 independent loci associated with BD at genome-wide significance (P < 5E-08) (Figure 1, Table 1, Table S2). Using LD Score regression (LDSC)^35^ the 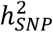of BD was estimated to be 18.6% (SE=0.008, P=5.1E-132) on the liability scale, assuming a BD population prevalence of 2%, and 15.6% (SE=0.006, P=5.0E-132) assuming a population prevalence of 1% (Table S3). The genomic inflation factor (λ_GC_) was 1.38 and the LD Score regression (LDSC) intercept was 1.04 (SE=0.01, P=2.5E-04)(Supplementary Figure 1). While the intercept has frequently been used as an indicator of confounding from population stratification, it can rise above 1 with increased sample size and heritability. The attenuation ratio - (LDSC intercept - 1)/(mean of association chi-square statistics - 1) - which is not subject to these limitations, was 0.06 (SE=0.02), indicating that the majority of inflation of the GWAS test statistics was due to polygenicity^35,36^. Of the 64 genome-wide significant loci, 33 are novel discoveries (ie. loci not overlapping with any locus previously reported as genome-wide significant for BD). Novel loci include the major histocompatibility complex (MHC) and loci previously reaching genome-wide significance for other psychiatric disorders, including 10 for schizophrenia, 4 for major depression and 3 for childhood-onset psychiatric disorders or problematic alcohol use (Table 1).

**Table 1:** Genome-wide siginificiant loci for bipolar disorder from meta-analysis of 41,917 cases and 371,549 controls.

| Locus | CHR | BP | SNP | P | OR | SE | A1/A2 | A1 freq in controls | Previous report<sup>A</sup> for BD (citation) | Name for novel locus<sup>+</sup> | Previous report<sup>A</sup> for psychiatric disorders |
| --- | --- | --- | --- | --- | --- | --- | --- | --- | --- | --- | --- |
| 1 | 1 | 61105668 | rs2126180 | **1.6E-09** | 1.058 | 0.009 | A/G | 0.457 |  | LINC01748 |  |
| 2 | 1 | 163745389 | rs10737496 | **7.2E-09** | 1.056 | 0.009 | C/T | 0.444 |  | NUF2 | CDG |
| 3* | 2 | 97416153 | rs4619651 | **4.8E-11** | 1.068 | 0.010 | G/A | 0.670 | LMAN2L (PGC2) |  | CDG |
| 4 | 2 | 166152389 | rs17183814 | **2.7E-08** | 1.108 | 0.019 | G/A | 0.924 | SCN2A (PGC2) |  |  |
| 5 | 2 | 169481837 | rs13417268 | **2.1E-08** | 1.064 | 0.011 | C/G | 0.758 |  | CERS6 |  |
| 6 | 2 | 193738336 | rs2011302 | **4.3E-08** | 1.055 | 0.010 | A/T | 0.377 |  | PCGEM1 | CDG |
| 7 | 2 | 194437889 | rs2719164 | **4.9E-08** | 1.053 | 0.010 | A/G | 0.564 | intergenic (PGC2) |  | CDG |
| 8* | 3 | 36856030 | rs9834970 | **6.6E-19** | 1.087 | 0.009 | C/T | 0.481 | TRANK1 (PGC2) |  | SCZ, CDG |
| 9* | 3 | 52626443 | rs2336147 | **3.6E-13** | 1.070 | 0.009 | T/C | 0.498 | ITIH1 (PGC2) |  | SCZ, CDG |
| 10 | 3 | 70488788 | rs115694474 | **2.4E-08** | 1.068 | 0.012 | T/A | 0.799 |  | MDFIC2 |  |
| 11 | 3 | 107757060 | rs696366 | **4.5E-08** | 1.053 | 0.009 | C/A | 0.550 | CD47 (PGC2) |  |  |
| 12* | 4 | 123076007 | rs112481526 | **1.9E-09** | 1.065 | 0.011 | G/A | 0.256 |  | KIAA1109 | MD |
| 13* | 5 | 7542911 | rs28565152 | **2.0E-09** | 1.070 | 0.011 | A/G | 0.238 | ADCY2 (PGC2) |  |  |
| 14* | 5 | 78849505 | rs6865469 | **1.7E-08** | 1.060 | 0.010 | T/G | 0.274 |  | HOMER1 |  |
| 15 | 5 | 80961069 | rs6887473 | **8.8E-09** | 1.062 | 0.011 | G/A | 0.739 | SSBP2 (PGC2) |  |  |
| 16* | 5 | 137712121 | rs10043984 | **3.7E-08** | 1.062 | 0.011 | T/C | 0.236 |  | KDM3B | CDG |
| 17 | 5 | 169289206 | rs10866641 | **2.8E-11** | 1.065 | 0.009 | T/C | 0.575 |  | DOCK2 |  |
| 18* | 6 | 26463575 | rs13195402 | **5.8E-15** | 1.146 | 0.018 | G/T | 0.919 |  | MHC | MD, SCZ, CDG, MOOD |
| 19* | 6 | 98565211 | rs1487445 | **1.5E-15** | 1.078 | 0.009 | T/C | 0.487 | POU3F2 (PGC2) |  | CDG |
| 20 | 6 | 152793572 | rs4331993 | **2.0E-08** | 1.056 | 0.010 | A/T | 0.382 | SYNE1 (Green, 2013) |  |  |
| 21* | 6 | 166995260 | rs10455979 | **4.2E-09** | 1.057 | 0.010 | G/C | 0.500 | RPS6KA2 (PGC2) |  |  |
| 22* | 7 | 2020995 | rs12668848 | **1.9E-09** | 1.059 | 0.010 | G/A | 0.575 | MAD1L1 (Hou, 2016, Ikeda, 2017) |  | MD, SCZ, CDG |
| 23* | 7 | 11871787 | rs113779084 | **1.4E-13** | 1.079 | 0.010 | A/G | 0.299 | THSD7A (PGC2) |  |  |
| 24* | 7 | 21492589 | rs6954854 | **5.9E-10** | 1.060 | 0.009 | G/A | 0.425 |  | SP4 |  |
| 25 | 7 | 24647222 | rs12672003 | **2.7E-09** | 1.096 | 0.016 | G/A | 0.113 |  | MPP6 | SCZ, CDG, MOOD |
| 26 | 7 | 105043229 | rs11764361 | **3.5E-09** | 1.063 | 0.010 | A/G | 0.668 | SRPK2 (PGC2) |  | SCZ, ASD, CDG |
| 27 | 7 | 131870597 | rs6946056 | **3.7E-08** | 1.055 | 0.010 | C/A | 0.623 |  | PLXNA4 |  |
| 28 | 7 | 140676153 | rs10255167 | **1.6E-08** | 1.068 | 0.012 | A/G | 0.778 | MRPS33 (PGC2) |  | CDG |
| 29* | 8 | 9763581 | rs62489493 | **2.6E-11** | 1.094 | 0.014 | G/C | 0.128 |  | miR124-1 | SCZ, ALC, ASD |
| 30* | 8 | 10226355 | rs3088186 | **2.1E-08** | 1.058 | 0.010 | T/C | 0.287 |  | MSRA | SCZ, ALC, ASD |
| 31 | 8 | 34152492 | rs2953928 | **6.3E-09** | 1.124 | 0.020 | A/G | 0.067 |  | RP1-84O15.2 (lincRNA) | SCZ, ADHD, CDG |
| 32* | 8 | 144993377 | rs6992333 | **1.6E-09** | 1.062 | 0.010 | G/A | 0.410 |  | PLEC |  |
| 33 | 9 | 37090538 | rs10973201 | **2.5E-08** | 1.101 | 0.017 | C/T | 0.110 |  | ZCCHC7 | MD, CDG, MOOD |
| 34* | 9 | 141066490 | rs62581014 | **2.8E-08** | 1.067 | 0.012 | T/C | 0.366 |  | TUBBP5 |  |
| 35* | 10 | 18751103 | rs1998820 | **4.1E-08** | 1.087 | 0.015 | T/A | 0.886 |  | CACNB2 | SCZ, CDG |
| 36* | 10 | 62322034 | rs10994415 | **1.1E-11** | 1.125 | 0.017 | C/T | 0.082 | ANK3 (PGC2) |  |  |
| 37 | 10 | 64525135 | rs10761661 | **4.7E-08** | 1.053 | 0.009 | T/C | 0.472 |  | ADO |  |
| 38* | 10 | 111648659 | rs2273738 | **1.6E-11** | 1.096 | 0.014 | T/C | 0.135 | ADD3 (Charney, 2017, PGC2) |  |  |
| 39* | 11 | 61618608 | rs174592 | **9.9E-14** | 1.074 | 0.010 | G/A | 0.395 | FADS2 (PGC2) |  | MD, CDG, MOOD |
| 40 | 11 | 64009879 | rs4672 | **3.4E-09** | 1.107 | 0.017 | A/G | 0.083 |  | FKBP2 |  |
| 41* | 11 | 65848738 | rs475805 | **2.0E-09** | 1.070 | 0.011 | A/G | 0.767 | PACS1 (PGC2) |  |  |
| 42* | 11 | 66324583 | rs678397 | **5.5E-09** | 1.056 | 0.009 | T/C | 0.457 | PC (PGC1, PGC2) |  |  |
| 43* | 11 | 70517927 | rs12575685 | **1.2E-10** | 1.067 | 0.010 | A/G | 0.327 | SHANK2 (PGC2) |  | MD |
| 44 | 11 | 79092527 | rs12289486 | **3.3E-08** | 1.086 | 0.015 | T/C | 0.115 | ODZ4 (PGC1) |  |  |
| 45* | 12 | 2348844 | rs11062170 | **1.9E-15** | 1.081 | 0.010 | C/G | 0.333 | CACNA1C (PGC2) |  | SCZ, CDG, MOOD |
| 46 | 13 | 113869045 | rs35306827 | **3.6E-09** | 1.068 | 0.011 | G/A | 0.775 |  | CUL4A |  |
| 47 | 14 | 99719219 | rs2693698 | **2.0E-08** | 1.055 | 0.009 | G/A | 0.551 |  | BCL11B | SCZ, CDG |
| 48* | 15 | 38973793 | rs35958438 | **3.8E-08** | 1.066 | 0.012 | G/A | 0.772 |  | C15orf53 | CDG |
| 49* | 15 | 42904904 | rs4447398 | **2.6E-09** | 1.086 | 0.014 | A/C | 0.131 | STARD9 (PGC2) |  |  |
| 50 | 15 | 83531774 | rs62011709 | **1.4E-08** | 1.064 | 0.011 | T/A | 0.747 |  | HOMER2 | SCZ |
| 51* | 15 | 85149575 | rs748455 | **5.0E-11** | 1.070 | 0.010 | T/C | 0.719 | ZNF592 (PGC2) |  | SCZ, CDG |
| 52 | 15 | 91426560 | rs4702 | **3.5E-09** | 1.059 | 0.010 | G/A | 0.446 |  | FURIN | SCZ, CDG |
| 53 | 16 | 9230816 | rs28455634 | **2.6E-10** | 1.065 | 0.010 | G/A | 0.620 |  | C16orf72 |  |
| 54 | 16 | 9926348 | rs7199910 | **1.7E-08** | 1.057 | 0.010 | G/T | 0.312 | GRIN2A (PGC2) |  | SCZ, CDG |
| 55 | 16 | 89632725 | rs12932628 | **6.7E-09** | 1.058 | 0.010 | T/G | 0.487 |  | RPL13 |  |
| 56 | 17 | 1835482 | rs4790841 | **3.1E-08** | 1.075 | 0.013 | T/C | 0.151 |  | RTN4RL1 |  |
| 57 | 17 | 38129841 | rs11870683 | **2.8E-08** | 1.059 | 0.010 | T/A | 0.650 | ERBB2 (Hou, 2016) |  |  |
| 58 | 17 | 38220432 | rs61554907 | **1.6E-08** | 1.091 | 0.015 | T/G | 0.124 | ERBB2 (Hou, 2016) |  |  |
| 59* | 17 | 42191893 | rs228768 | **2.8E-10** | 1.067 | 0.010 | G/T | 0.294 | HDAC5 (PGC2) |  |  |
| 60* | 20 | 43682551 | rs67712855 | **4.2E-11** | 1.070 | 0.010 | T/G | 0.687 | STK4 (PGC2) |  |  |
| 61* | 20 | 43944323 | rs6032110 | **1.0E-09** | 1.059 | 0.009 | A/G | 0.512 | WFDC12 (PGC2) |  |  |
| 62* | 20 | 48033127 | rs237460 | **4.3E-09** | 1.057 | 0.009 | T/C | 0.412 |  | KCNB1 | CDG |
| 63 | 20 | 60865815 | rs13044225 | **8.5E-09** | 1.056 | 0.010 | G/A | 0.440 |  | OSBPL2 |  |
| 64 | 22 | 41153879 | rs5758064 | **2.0E-08** | 1.054 | 0.009 | T/C | 0.523 |  | SLC25A17 | MD, SCZ, CDG, MOOD |

CHR, chromosome; BP, GRCh37 basepair position; SNP, single nucleotide polymorphism; OR, odds ratio; SE, standard error, A1, tested allele; A2, other allele; freq, frequency; BD, bipolar disorder; CDG, Cross-disorder GWAS of the Psychiatric Genomics Consortium; MD, major depression; SCZ, schizophrenia; MOOD, mood disorders; ASD, Autism Spectrum Disorder; ALC, Alcohol use disorder or problematic alcohol use; ADHD, attention deficit/hyperactivity disorder. \*Locus overlaps with genome-wide significant locus for bipolar I disorder. <sup>A</sup>Previous report refers to previous association of a SNP in the locus with the psychiatric disorder at genome-wide significance. PGC1 = PMID 21926972, PGC2 = PMID 31043756, Hou, 2016 = PMID 27329760, Ikeda, 2017 = PMID:28115744, Green, 2013 = PMID 22565781. Charney, 2017 = PMID 28072414. <sup>+</sup>Novel loci are named using the nearest gene to the index SNP.

**Figure 1:**
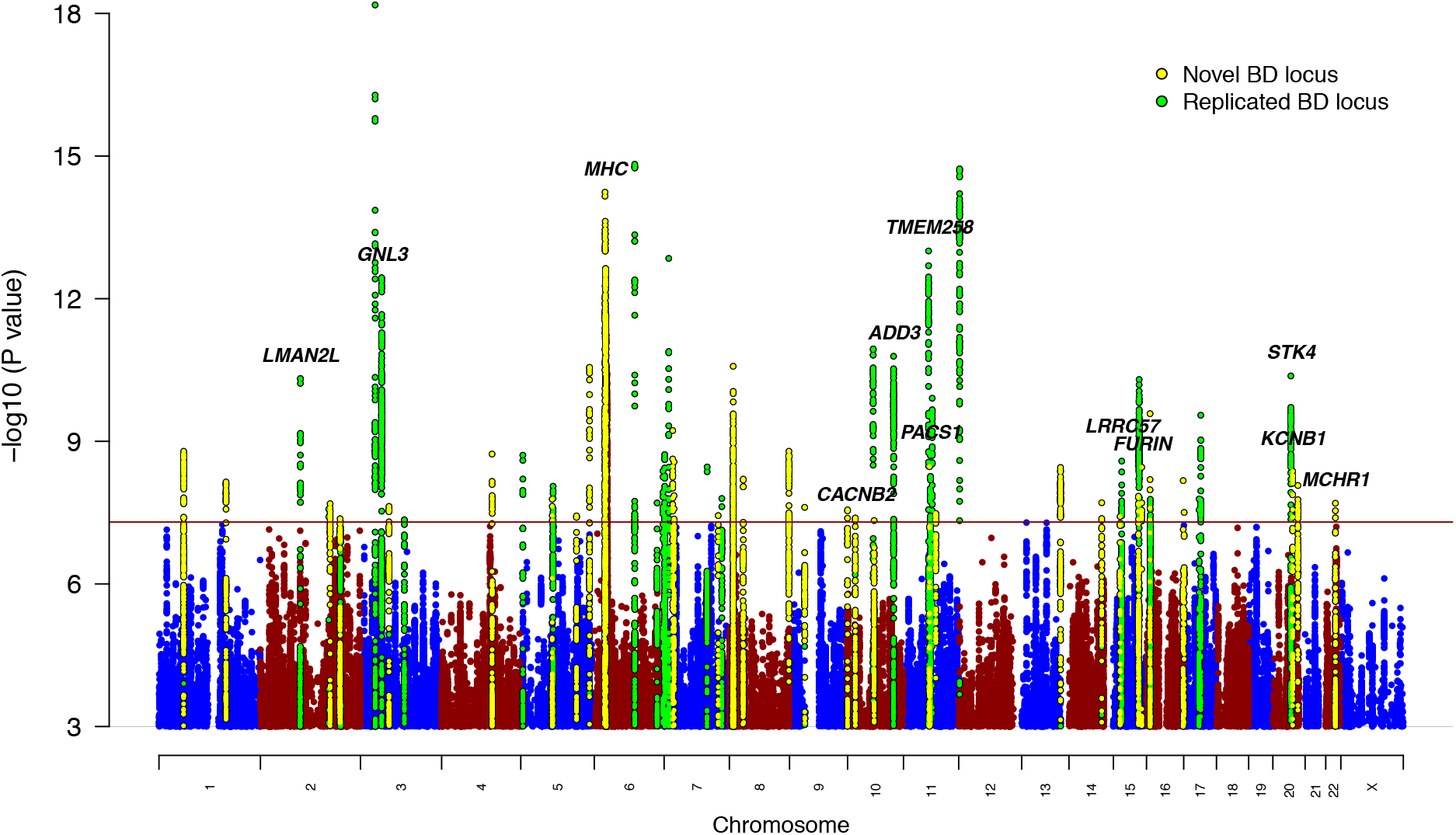
Manhattan plot of genome-wide association meta-analysis of 41,917 bipolar disorder cases and 371,549 controls. The *x-*axis shows genomic position (chromosomes 1-22 and X) and the *y-*axis shows statistical significance as –log_10_(P value). P values are two-sided and based on an inverse variance weighted fixed effects meta-analysis. The red line shows the genome-wide significance threshold (P<5E-08). SNPs in genome-wide significant loci are colored green for loci previously associated with bipolar disorder (BD) and yellow for novel associations from this study. The genes labeled are those prioritized by integrative eQTL analyses or notable genes in novel loci (*MHC, CACNB2, KCNB1*).

### Enrichment analyses

Genome-wide analyses using MAGMA^37^ indicated significant enrichment of BD associations in 161 genes (Table S4) and 4 gene sets, related to synaptic signaling (Table S5). The BD association signal was enriched amongst genes expressed in different brain tissues (Table S6), especially genes with high specificity of gene expression in neurons (both excitatory and inhibitory) versus other cell types, within cortical and subcortical brain regions in mice (Supplementary Figure 2)^38^. In human brain samples, signal enrichment was also observed in hippocampal pyramidal neurons and interneurons of the prefrontal cortex and hippocampus, compared with other cell types (Supplementary Figure 2).

**Figure 2:**
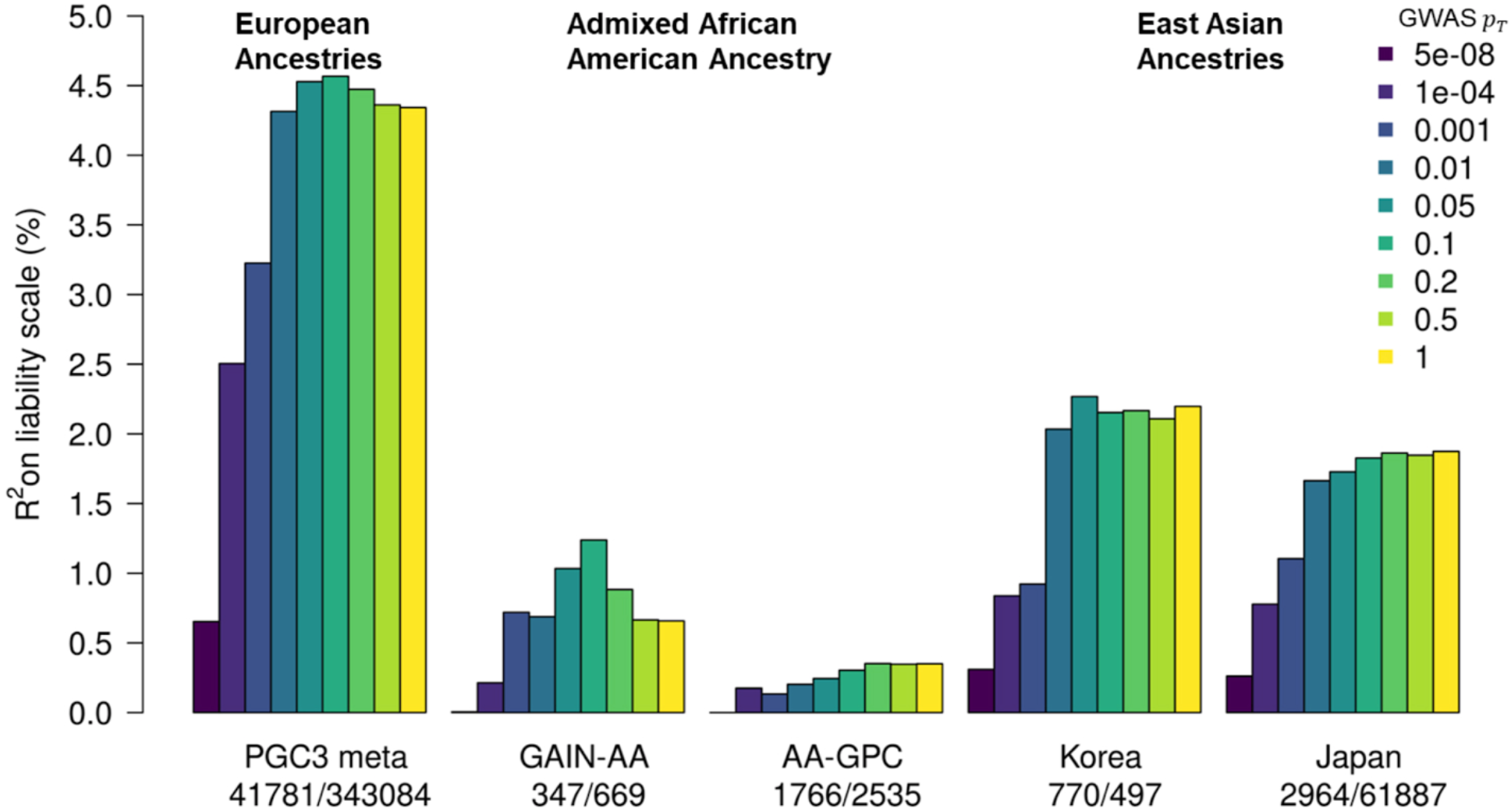
Phenotypic variance in bipolar disorder explained by polygenic risk scores. Variance explained is presented on the liability scale, assuming a 2% population prevalence of bipolar disorder. For European ancestries, the results shown are the weighted mean R^2^ values across all 57 cohorts in the PGC3 meta-analysis, weighted by the effective N per cohort. The numbers of cases and controls are shown from left to right under the barplot for each study. GWAS p_T_ - the color of the bars represents the P value threshold used to select SNPs from the discovery GWAS. GAIN-AA - Genetic Association Information Network African American cohort, AA-GPC - African American Genomic Psychiatry Cohort.

In a gene-set analysis of the targets of individual drugs (from the Drug-Gene Interaction Database DGIdb v.2^39^ and the Psychoactive Drug Screening Database Ki DB^40^), the targets of the calcium channel blockers mibefradil and nisoldipine were significantly enriched (Table S7). Grouping drugs according to their Anatomical Therapeutic Chemical (ATC) classes^41^, there was significant enrichment in the targets of four broad drug classes (Table S8): psycholeptics (drugs with a calming effect on behavior) (especially hypnotics and sedatives, antipsychotics and anxiolytics), calcium channel blockers, antiepileptics and (general) anesthetics. (Table S8).

### eQTL integrative analyses

A transcriptome-wide association study (TWAS) was conducted using FUSION^42^ and eQTL data from the PsychENCODE Consortium (1,321 brain samples)^43^. BD-associated alleles significantly influenced expression of 77 genes in the brain (Table S9, Supplementary Figure 3). These genes encompassed 40 distinct regions. TWAS fine-mapping was performed using FOCUS^44^ to model the correlation among the TWAS signals and prioritize the most likely causal gene(s) in each region. Within the 90%-credible set, FOCUS prioritised 22 genes with a posterior inclusion probability (PIP) > 0.9 (encompassing 20 distinct regions) and 32 genes with a PIP > 0.7 (29 distinct regions) (Table S10).

**Figure 3:**
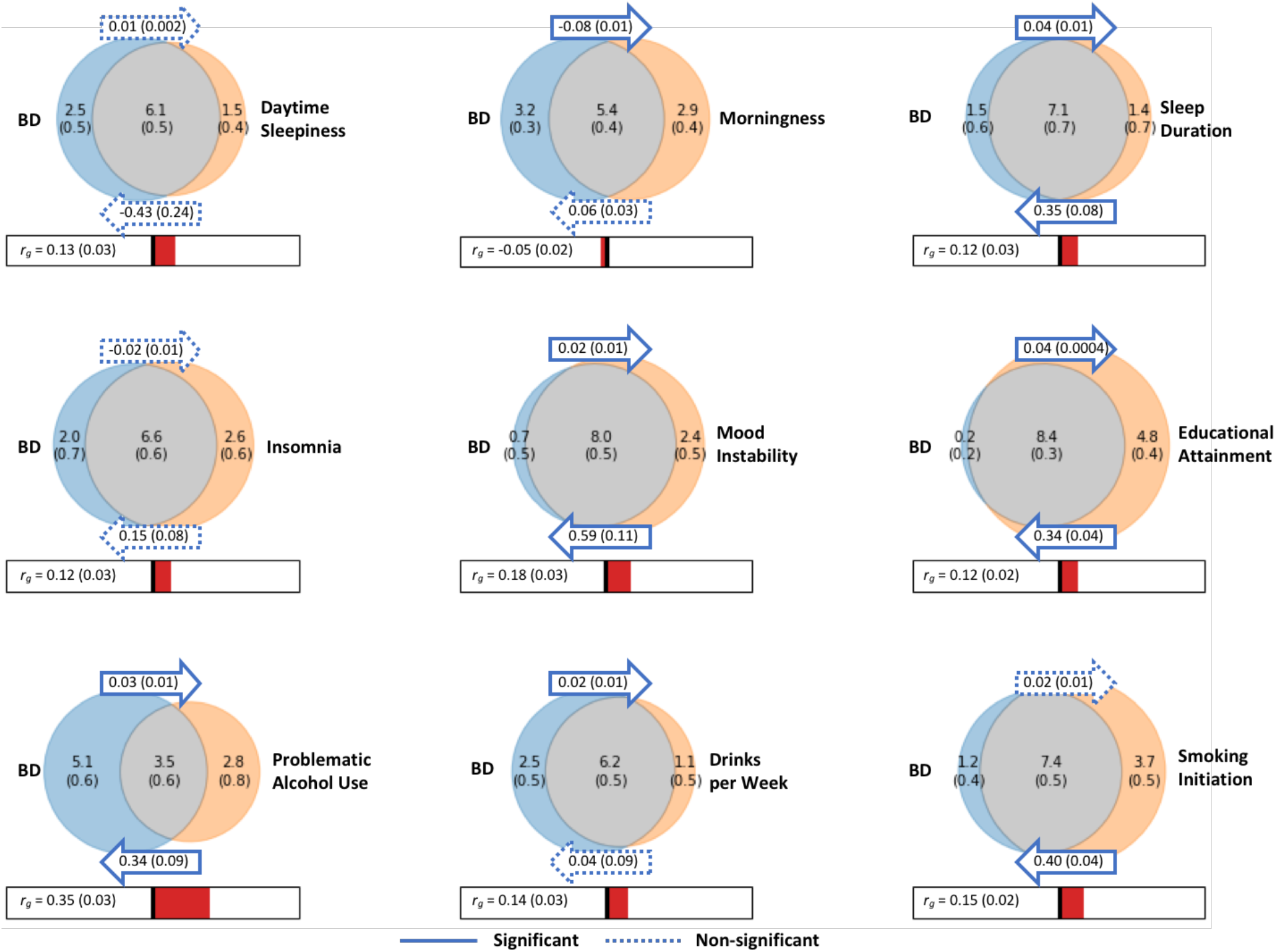
Relationships between bipolar disorder and modifiable risk factors based on genetic correlations, generalized summary statistics-based Mendelian randomization and bivariate gaussian mixture modeling. Venn diagrams depict MiXeR results of the estimated number of influencing variants shared between bipolar disorder (BD) and each trait of interest (grey), unique to BD (blue) and unique to the trait of interest (orange). The number of influencing variants and standard error are shown in thousands. The size of the circles reflects the polygenicity of each trait, with larger circles corresponding to greater polygenicity. The estimated genetic correlation (r_g_) between BD and each trait of interest and standard error from LD Score regression is shown below the corresponding Venn diagram, with an accompanying scale (−1 to +1). The arrows above and below the Venn diagrams indicate the results of generalized summary statistics-based Mendelian randomization (GSMR) of BD on the trait of interest, and the trait of interest on BD, respectively. The GSMR effect size and standard error is shown inside the corresponding arrow. Solid arrows indicate a significant relationship between the exposure and the outcome, after correction for multiple comparisons (P<1.47E-03) and dashed arrows indicate a non-significant relationship.

Summary data-based Mendelian randomization (SMR)^45,46^ was used to identify putative causal relationships between SNPs and BD via gene expression by integrating the BD GWAS results with brain eQTL summary statistics from the PsychENCODE^43^ Consortium and blood eQTL summary statistics from the eQTLGen Consortium (31,684 whole blood samples)^47^. The eQTLGen results represent the largest existing eQTL study and provide independent eQTL data. Of the 32 genes fine-mapped with PIP > 0.7, 15 were significantly associated with BD in the SMR analyses and passed the HEIDI (heterogeneity in dependent instruments) test^45,46^, suggesting that their effect on BD is mediated via gene expression in the brain and/or blood (Table S11). The genes located in genome-wide significant loci are labeled in Figure 1. Other significant genes included *HTR6, DCLK3, HAPLN4* and *PACSIN2*.

### MHC locus

Variants within and distal to the major histocompatibility complex (MHC) locus were associated with BD at genome-wide significance. The most highly associated SNP was rs13195402, 3.2 megabases distal to any *HLA* gene or the complement component 4 (*C4*) genes (Supplementary Figure 4). Imputation of *C4* alleles using SNP data uncovered no association between the five most common structural forms of the *C4A/C4B* locus (BS, AL, AL-BS, AL-BL, and AL-AL) and BD, either before or after conditioning on rs13195402 (Supplementary Figure 5). While genetically predicted *C4A* expression initially showed a weak association with BD, this association was non-significant after controlling for rs13195402 (Supplementary Figure 6).

### Polygenic risk scoring

The performance of polygenic risk scores (PRS) based on these GWAS results was assessed by excluding cohorts in turn from the meta-analysis to create independent test samples. PRS explained ∼4.57% of phenotypic variance in BD on the liability scale (at GWAS P value threshold (p_T_) < 0.1, BD population prevalence 2%), based on the weighted mean R^2^ across cohorts (Figure 2, Table S12). This corresponds to a weighted mean area under the curve (AUC) of 65%. Results per cohort and per wave of recruitment to the PGC are in Tables S12-S13 and Supplementary Figure 7. At p_T_ < 0.1, individuals in the top 10% of BD PRS had an odds ratio of 3.5 (95% CI 1.7-7.3) of being affected with the disorder compared with individuals in the middle decile (based on the weighted mean OR across PGC cohorts), and an odds ratio of 9.3 (95% CI 1.7-49.3) compared with individuals in the lowest decile. The generalizability of PRS from this meta-analysis was examined in several non-European cohorts. PRS explained up to 2.3% and 1.9% of variance in BD in two East Asian samples, and 1.2% and 0.4% in two admixed African American samples (Figure 2, Table S14). The variance explained by the PRS increased in every cohort with increasing sample size of the PGC BD European discovery sample (Supplementary Figure 8, Table S14).

### Genetic architecture of BD and other traits

The genome-wide genetic correlation (r_g_) of BD with a range of diseases and traits was assessed on LD Hub^48^. After correction for multiple testing, BD showed significant r_g_ with 16 traits among 255 tested from published GWAS (Table S15). Genetic correlation was positive with all psychiatric disorders assessed, particularly schizophrenia (r_g_ = 0.68) and major depression (r_g_=0.44), and to a lesser degree anorexia, attention deficit/hyperactivity disorder and autism spectrum disorder (r_g_≈ 0.2). We found evidence of positive r_g_ between BD and smoking initiation, cigarettes per day, problematic alcohol use and drinks per week (Figure 3). BD was also positively genetically correlated with measures of sleep quality (daytime sleepiness, insomnia, sleep duration) (Figure 3). Among 514 traits measured in the general population of the UK Biobank, there was significant r_g_ between BD and many psychiatric-relevant traits or symptoms, dissatisfaction with interpersonal relationships, poorer overall health rating and feelings of loneliness or isolation (Table S16).

Bivariate gaussian mixture models were applied to the GWAS summary statistics for BD and other complex traits using the MiXeR tool^49,50^ to estimate the number of variants influencing each trait that explain 90% of 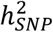 and their overlap between traits. MiXeR estimated that approximately 8.6 k (SE=0.2 k) variants influence BD, which is similar to the estimate for schizophrenia (9.7 k, SE=0.2 k) and somewhat lower than that for major depression (12.3 k, SE=0.6 k) (Table S17, Supplementary Figure 9). When considering the number of shared loci as a proportion of the total polygenicity of each trait, the vast majority of loci influencing BD were also estimated to influence major depression (97%) and schizophrenia (96%) (Table S17, Supplementary Figure 9). Interestingly, within these shared components, the variants that influenced both BD and schizophrenia had high concordance in direction of effect (80%, SE=2%), while the portion of concordant variants between BD and MDD was only 69% (SE=1%) (Table S17).

### Genetic and causal relationships between BD and modifiable risk factors

Ten traits associated with BD from clinical and epidemiological studies were investigated in detail for genetic and potentially causal relationships with BD via LDSC^35^, generalized summary statistics-based Mendelian randomization (GSMR)^51^ and bivariate gaussian mixture modeling^49^. BD has been strongly linked with sleep disturbances^52^, alcohol use^53^ and smoking^54^, higher educational attainment^55,56^ and mood instability^57^. Most of these traits had modest but significant genetic correlations with BD (r_g_ −0.05-0.35) (Figure 3). Examining the effects of these traits on BD via GSMR, smoking initiation was associated with BD, corresponding to an OR of 1.49 (95% CI 1.38-1.61) for developing the disorder (P=1.74E-22) (Figure 3). Testing the effect of BD on the traits, BD was significantly associated with reduced likelihood of being a morning person and increased number of drinks per week (P<1.47E-03) (Figure 3). Positive bi-directional relationships were identified between BD and longer sleep duration, problematic alcohol use, educational attainment (EA) and mood instability (Figure 3). Notably, the instrumental variables for mood instability were selected from a GWAS conducted in the general population, excluding individuals with psychiatric disorders^58^. For all of the aforementioned BD-trait relationships, the effect size estimates from GSMR were consistent with those calculated using the inverse variance weighted regression method, and there was no evidence of bias from horizontal pleiotropy. Full MR results are in Tables S18-19. Bivariate gaussian mixture modeling using MiXeR, indicated large proportions of variants influencing both BD and all other traits tested, particularly educational attainment, where approximately 98% of variants influencing BD were estimated to also influence EA. While cigarettes per day was a trait of interest, MiXeR could not model these data due to low polygenicity and heritability, and the effect of cigarettes per day on BD was inconsistent between MR methods, suggesting a violation of MR assumptions (Tables S18-20).

### BD subtypes

We conducted GWAS meta-analyses of bipolar I disorder (BD I) (25,060 cases, 449,978 controls) and bipolar II disorder (BD II) (6,781 cases, 364,075 controls). The BD I analysis identified 44 genome-wide significant loci, 31 of which overlapped with genome-wide significant loci from the main BD GWAS (Table 1, Table S21). The remaining 13 genome-wide significant loci for BD I all had P < 4.0E-05 in the main BD GWAS. One genome-wide significant locus was identified in the GWAS meta-analysis of BD II and had a P < 1.1E-04 in the main GWAS of BD (Table S21). The 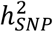 estimates on the liability scale for BD I and BD II were 20.9% (SE=0.009, P=1.0E-111) and 11.6% (SE=0.01, P = 3.9E-15), respectively, assuming a 1% population prevalence of each subtype. These heritability values are significantly different from each other (P=2.4E-25, block jackknife). The genetic correlation between BD I and BD II was 0.85 (SE=0.05, P = 2.88E-54), which is significantly different from 1 (P=1.6E-03). The genetic correlation of BD I with schizophrenia (r_g_=0.66, SE=0.02) was higher than that of BD II (r_g_=0.54 SE=0.05), whereas major depression was more strongly genetically correlated with BD II (r_g_=0.66, SE=0.05) than with BD I (r_g_=0.34, SE=0.03) (Table S22).

## Discussion

In a GWAS of 41,917 BD cases, we identify 64 associated genomic loci, 33 of which are novel discoveries. With a 1.5-fold increase in effective sample size compared with the PGC2 BD GWAS, this study more than doubled the number of associated loci, representing an inflection point in the rate of risk variant discovery. We observed consistent replication of known BD loci, including 28/30 loci from the PGC2 GWAS^24^ and several implicated by other BD GWAS^15,16,17^, including a study of East Asian cases^59^.

The 33 novel loci discovered here encompass genes of expected biological relevance to BD, such as the ion channels *CACNB2* and *KCNB1*. Amongst the 64 BD loci, 17 have previously been implicated in GWAS of schizophrenia^60^, and seven in GWAS of major depression^61^, representing the first overlap of genome-wide significant loci between the mood disorders. For these genome-wide significant loci shared across disorders, 17/17 and 5/7 of the BD index SNPs had the same direction of effect on schizophrenia and major depression respectively (Table S23). More generally, 50/64 and 62/64 BD loci had a consistent direction of effect on major depression and schizophrenia respectively, considerably greater than chance (P<1E-05, binomial test). Bivariate gaussian mixture modeling estimated that across the entire genome, almost all variants influencing BD also influence schizophrenia and major depression, albeit with variable effects^62^. SNPs in and around the MHC locus reached genome-wide significance for BD for the first time. However, unlike in schizophrenia, we found no influence of *C4* structural alleles or gene expression^63^. Rather the association was driven by variation outside the classical MHC locus, with the index SNP (rs13195402) being a missense variant in *BTN2A1*, a brain-expressed gene^64^ encoding a plasma membrane protein.

The genetic correlation of BD with other psychiatric disorders was consistent with previous reports^65,66^. Our results also corroborate previous genetic and clinical evidence of associations between BD and sleep disturbances^67^, problematic alcohol use^68^ and smoking^69^. While the genome-wide genetic correlations with these traits were modest (r_g_ −0.05-0.35), MiXeR estimated that for all traits, more than 55% of trait-influencing variants also influence BD (Figure 3). Taken together, these results point to shared biology as one possible explanation for the high prevalence of substance use in BD. However, excluding genetic variants associated with both traits, MR analyses suggested that smoking is also a putatively “causal” risk factor for BD, while BD has no effect on smoking, consistent with a previous report^70^. [We use the word “causal” with caution here as we consider MR an exploratory analysis to identify potentially modifiable risk factors which warrant more detailed investigations to understand their complex relationship with BD.] In contrast, MR indicated that BD had bi-directional “causal” relationships with problematic alcohol use, longer sleep duration and mood instability. Insights into the relationship of such behavioral correlates with BD may have future impact on clinical decision making in the prophylaxis or management of the disorder. Higher educational attainment has previously been associated with BD in epidemiological studies^55,56^, while lower educational attainment has been associated with schizophrenia and major depression^71,72^. Here, educational attainment had a significant positive effect on risk of BD and vice versa. Interestingly, MiXeR estimated that almost all variants that influence BD also influence educational attainment. The substantial genetic overlap observed between BD and the other phenotypes suggests that many variants likely influence multiple phenotypes which may be differentiated by phenotype-specific effect size distributions among the shared influencing variants.

The integration of eQTL data with our GWAS results yielded 15 high-confidence genes for which there was converging evidence that their association with BD is mediated via gene expression. Amongst these were *HTR6*, encoding a serotonin receptor targeted by antipsychotics and antidepressants^73^ and *MCHR1* (melanin-concentrating hormone receptor 1), encoding a target of the antipsychotic haloperidol^73^. We note that for both of these genes, their top eQTLs have opposite directions of effect on gene expression in the brain and blood, possibly playing a role in the tissue-specific gene regulation influencing BD^74^. BD was associated with decreased expression of *FURIN*, a gene with a neurodevelopmental role which has already been the subject of functional genomics experiments in neuronal cells, following its association with schizophrenia in GWAS^75^. The top association in our GWAS was in the *TRANK1* locus on chromosome 3, which has previously been implicated in BD^12,18,59^. Although BD-associated SNPs in this locus are known to regulate *TRANK1* expression^76^, our eQTL analyses support a stronger but correlated regulation of *DCLK3*, located 87 kb upstream of *TRANK1*^*43,77*^. Both *FURIN* and *DCLK3* also encode druggable proteins (although they are not targets for any current psychiatric medications)^73,78^. These eQTL results provide promising BD candidate genes for functional follow-up experiments^29^. While several of these are in genome-wide significant loci, many are not the closest gene to the index SNP, highlighting the value of probing underlying molecular mechanisms to prioritize the most likely causal genes in the loci.

GWAS signals were enriched in the gene targets of existing BD pharmacological agents, such as antipsychotics, mood stabilizers, and antiepileptics. However, enrichment was also found in the targets of calcium channel blockers used to treat hypertension and GABA-receptor targeting anesthetics (Table S8). Calcium channel antagonists have long been investigated for the treatment of BD, without becoming an established therapeutic approach, and there is evidence that some antiepileptics have calcium channel-inhibiting effects^79,80^. These results underscore the opportunity for repurposing some classes of drugs, particularly calcium channel antagonists, as potential BD treatments^81^.

BD associations were enriched in gene sets involving neuronal parts and synaptic signaling. Neuronal and synaptic pathways have been described in cross-disorder GWAS of multiple psychiatric disorders including BD^82–84^. Dysregulation of such pathways has also been suggested by previous functional and animal studies^85^. Analysis of single-cell gene expression data revealed enrichment in genes with high specificity of gene expression in neurons (both excitatory and inhibitory), of many brain regions, in particular the cortex and hippocampus. These findings are similar to those reported in GWAS data of schizophrenia^86^ and major depressive disorder^38^.

PRS for BD explained on average 4.57% of phenotypic variance (liability scale) across European cohorts, although this varied in different waves of the BD GWAS, ranging from 6.6% in the PGC1 cohorts to 2.9% in the External biobank studies (Supplementary Figure 7, Table S12). These results are in line with the 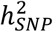 of BD per wave, which ranged from 24.6% (SE=0.01) in PGC1 to 11.9% (SE=0.01) in External studies (Table S3). Some variability in 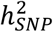 estimates may arise from the inclusion of cases from population biobanks, who may have more heterogeneous clinical presentations or less severe illness than BD patients ascertained via inpatient or outpatient psychiatric clinics. Across the waves of clinically ascertained samples within the PGC, 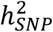 and the R^2^ of PRS also varied, likely reflecting clinical and genetic heterogeneity in the type of BD cases ascertained; the PGC1 cohorts consisted mostly of BD I cases^9^, known to be the most heritable of the BD subtypes^11,24^, while later waves included more individuals with BD II^24^. Overall, the 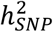 of BD calculated from the meta-analysis summary statistics was 18% on the liability scale, a decrease of ∼2% compared with the PGC2 GWAS^24^, which may be due to the addition of cohorts with lower 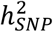 estimates and heterogeneity between cohorts (Table S3). However, despite differences in 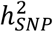 and R^2^ of PRS per wave, the genetic correlation of BD between all waves was high (weighted mean r_g_=0.94, SE=0.03), supporting our rationale for combining cases with different BD subtypes or ascertainment to increase power for discovery of risk variants. In Europeans, individuals in the top 10% of PRS had an OR of 3.5 for BD, compared with individuals with average PRS (middle decile), which translates into a modest absolute lifetime risk of the disorder (7% based on PRS alone). While PRS are invaluable tools in research settings, the current BD PRS lack sufficient power to separate individuals into clinically meaningful risk categories, and therefore have no clinical utility at present^87,88^. PRS from this European BD meta-analysis yield higher R^2^ values in diverse ancestry samples than PRS based on any currently available BD GWAS within the same ancestry^59^. However, performance still greatly lags behind that in Europeans, with ∼2% variance explained in East Asian samples and substantially less in admixed African American samples, likely due to differences in allele frequencies and LD structures, consistent with previous studies^89,90^. There is a pressing need for more and larger studies in other ancestry groups to ensure that any future clinical utility is broadly applicable. Exploiting the differences in LD structure between diverse ancestry samples will also assist in the fine-mapping of risk loci for BD.

Our analyses confirmed that BD is a highly polygenic disorder, with an estimated 8.6 k variants explaining 90% of its 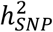. Hence, many more SNPs than those identified here are expected to account for the common variant architecture underlying BD. This GWAS marks an inflection point in risk variant discovery and we expect that from this point forward, the addition of more samples will lead to a dramatic increase in genetic findings. Nevertheless, fewer genome-wide significant loci have been identified in BD than in a schizophrenia GWAS of comparable sample size^60^. This may be due to the clinical and genetic heterogeneity that exists in BD.

Our GWAS of subtypes BD I and BD II identified additional associated loci. Consistent with previous findings^24^, our analysis showed that the two subtypes were highly but imperfectly genetically correlated (r_g_=0.85), and that BD I is more genetically correlated with schizophrenia, while BD II has stronger genetic correlation with major depression. The subtypes are sufficiently similar to justify joint analysis as BD, but are not identical in their genetic composition, and as such contribute to the genetic heterogeneity of BD^91^. We identified thirteen loci passing genome-wide significance for BD I, and one for BD II, which did not reach significance in the main BD GWAS, further illustrating the partially differing genetic composition of the two subtypes. Understanding the shared and distinct genetic components of BD subtypes and symptoms requires detailed phenotyping efforts in large cohorts and is an important area for future psychiatric genetics research.

In summary, these new data advance our understanding of the biological etiology of BD and prioritize a set of candidate genes for functional follow-up experiments. Several lines of evidence converge on the involvement of calcium channel signaling, providing a promising avenue for future therapeutic development.

## Data Availability

GWAS summary statistics are publicly available on the PGC website (https://www.med.unc.edu/pgc/results-and-downloads). Individual-level data are accessible through collaborative analysis proposals to the Bipolar Disorder Working Group of the PGC (https://www.med.unc.edu/pgc/shared-methods/how-to/). This study included some publicly available datasets accessed through dbGaP (PGC bundle phs001254.v1.p1) and the Haplotype Reference Consortium reference panel v1.0 (http://www.haplotype-reference-consortium.org/home). Databases used: Drug-Gene Interaction Database DGIdb v.2 https://www.dgidb.org Psychoactive Drug Screening Database Ki DB https://pdsp.unc.edu/databases/kidb.php DrugBank 5.0 www.drugbank.ca LDHub http://ldsc.broadinstitute.org FUMA https://fuma.ctglab.nl

## Acknowledgements

We thank the participants who donated their time, life experiences and DNA to this research and the clinical and scientific teams that worked with them. We are deeply indebted to the investigators who comprise the PGC. The PGC has received major funding from the US National Institute of Mental Health (PGC3: U01 MH109528; PGC2: U01 MH094421; PGC1: U01 MH085520). Statistical analyses were carried out on the NL Genetic Cluster Computer (http://www.geneticcluster.org) hosted by SURFsara and the Mount Sinai high performance computing cluster (http://hpc.mssm.edu), which is supported by the Office of Research Infrastructure of the National Institutes of Health under award numbers S10OD018522 and S10OD026880. The content is solely the responsibility of the authors and does not necessarily represent the official views of the National Institutes of Health. Full acknowledgements are included in the Supplementary Note.

## Author contributions

### Writing group

N.M., A.J.F., K.S.O’C., B.C., J.R.I.C., J.M.B., J.I.N., S.Cichon, H.J.E., E.A.S., A.McQuillin, A.D.F., R.A.O., O.A.A.

### PGC BD PI group

A.J.F., M.I., H-H.W., D.C., R.A., I.A., M.A., L.Alfredsson, G.Babadjanova, L.B., B.T.B., F.B., S.Bengesser, W.H.B., D.H.R.B., M.Boehnke, A.D.B., G.Breen, V.J.C., S.Catts, A.C., N.C., U.D., D.D., T.Esko, B.E., P.F., M.F., J.M.F., M.G., E.S.G., F.S.G., M.J.Green, M.G-S., J.Hauser, F.H., J.Hillert, K.S.H., D.M.H., C.M.H., K.Hveem, N.I., A.V.J., I.J., L.A.J., R.S.K., J.R.K., G.K., M.Landén, M.Leboyer, C.M.L., Q.S.L., J.Lissowska, C.Lochner, C.Loughland, N.G.M., C.A.M., F.M., S.L.M., A.M.M., F.J.M., I.M., P.Michie, L.Milani, P.B.Mitchell, G.M., O.M., P.B.Mortensen, B.M., B.M-M., R.M.M., B.M.N., C.M.N., M.N., M.M.N., M.C.O’D., K.J.O., T.O., M.J.O., S.A.P., C.Pantelis, C.Pato, M.T.P., G.P.P., R.H.P., D.P., J.A.R-Q., A.R., E.Z.R., M.Ribasés, M.Rietschel, S.R., G.A.R., T.S., U.S., M.S., P.R.S., T.G.S., L.J.S., R.J.S., A.S., C.S.W., J.W.S., H.S., K.S., E.Stordal, F.Streit, P.F.S., G.T., A.E.V., E.V., J.B.V., I.D.W., T.W.W., T.W., N.R.W., J-A.Z., J.M.B., J.I.N., S.Cichon, H.J.E., E.A.S., A.McQuillin, A.D.F., R.A.O., O.A.A.

### Bioinformatics

N.M., A.J.F., J.R.I.C., S.Børte, M.J.Gandal, M.Kim, B.M.S., L.G.S., B.S.W., H-H.W., N.A-R., S.E.B., B.M.B., V.E-P., S.H., P.A.H., Y.K., M.Koromina, M.Kubo, M.Leber, P.H.L., C.Liao, L.M.O.L., T.R., P.R., P.D.S., M.S.A., C.Terao, T.E.T., S.X., H.Y., P.P.Z., S.Bengesser, G.Breen, P.F., E.S.G., Q.S.L., G.A.R., H.S., T.W., E.A.S.

### Clinical

O.K.D., M.I., L.Abramova, K.A., E.A., N.A-R., A.Anjorin, A.Antoniou, J.H.B., N.B., M.Bauer, A.B., C.B.P., E.B., M.P.B., R.B., M.Brum, N.B-K., M.Budde, W.B., M.Cairns, M.Casas, P.C., A.C-B., D.C., P.M.C., N.D., A.D., T.Elvsåshagen, L.Forty, L.Frisén, K.G., J.Garnham, M.G.P., I.R.G., K.G-S., J.Grove, J.G-P., K.Ha, M.Haraldsson, M.Hautzinger, U.H., D.H., J.L.Kalman, J.L.Kennedy, S.K-S., M.Kogevinas, T.M.K., R.K., S.A.K., J.L., H-J.L., C.Lewis, S.L., M.Lundberg, D.J.M., W.M., D.M., L.Martinsson, M.M., P.McGuffin, H.M., V.M., C.O’D., L.O., S.P., A.Perry, A.Pfennig, E.P., J.B.P., D.Q., M.H.R., J.R.D., E.J.R., J.P.R., F.R., J.R., E.C.S., F.Senner, E.Sigurdsson, L.S., C.S., O.B.S., D.J.Smith, J.L.S., A.T.S., J.S.S., B.S., P.T., M.P.V., H.V., A.H.Y., L.Z., HUNT All-In Psychiatry, R.A., I.A., M.A., G.Babadjanova, L.B., B.T.B., F.B., S.Bengesser, D.H.R.B., A.D.B., A.C., N.C., U.D., D.D., B.E., P.F., M.F., M.G., E.S.G., F.S.G., M.J.Green, M.G-S., J.Hauser, K.S.H., N.I., I.J., L.A.J., R.S.K., G.K., M.Landén, C.M.L., J.Lissowska, N.G.M., C.A.M., F.M., S.L.M., A.M.M., I.M., P.B.Mitchell, G.M., O.M., P.B.Mortensen, M.C.O’D., K.J.O., M.J.O., C.Pato, M.T.P., R.H.P., J.A.R-Q., A.R., E.Z.R., M.Rietschel, T.S., T.G.S., A.S., C.S.W., J.W.S., E.Stordal, F.Streit, A.E.V., E.V., J.B.V., I.D.W., T.W.W., T.W., J.I.N., A.McQuillin, A.D.F.

### Genomic assays/data generation

A.J.F., M.I., E.A., M.A.E., D.A., M.B-H., E.C.B., C.B.P., J.B-G., M.Cairns, T-K.C., C.C., J.C., F.S.D., F.D., S.D., A.F., J.F., N.B.F., J.Gelernter, M.G.P., P.H., S.J., Y.K., H.R.K., M.Kubo, S.E.L., C.Liao, E.M., N.W.M., J.D.M., G.W.M., J.L.M., D.W.M., T.W.M., N.O’B., M.Rivera, C.S-M., S.Sharp, C.S.H., C.Terao, C.Toma, E-E.T., S.H.W., HUNT All-In Psychiatry, G.Breen, A.C., T.Esko, J.M.F., E.S.G., D.M.H., N.I., F.J.M., L.Milani, R.M.M., M.M.N., M.Ribasés, G.A.R., T.S., G.T., S.Cichon

### Obtained funding for BD samples

M.I., M.Cairns, I.N.F., L.Frisén, S.J., Y.K., J.A.K., M.Kubo, C.Lavebratt, S.L., D.M., P.McGuffin, G.W.M., J.B.P., M.H.R., J.R.D., D.J.Stein, J.S.S., C.Terao, A.H.Y., P.P.Z., M.A., L.Alfredsson, L.B., B.T.B., F.B., W.H.B., M.Boehnke, A.D.B., G.Breen, A.C., N.C., B.E., M.F., J.M.F., E.S.G., M.J.Green, M.G-S., K.S.H., K.Hveem, N.I., I.J., L.A.J., M.Landén, M.Leboyer, N.G.M., F.J.M., P.B.Mitchell, O.M., P.B.Mortensen, B.M.N., M.N., M.M.N., M.C.O’D., T.O., M.J.O., C.Pato, M.T.P., G.P.P., M.Rietschel, G.A.R., T.S., M.S., P.R.S., T.G.S., C.S.W., J.W.S., G.T., J.B.V., T.W.W., T.W., J.M.B., J.I.N., H.J.E., R.A.O.

### Statistical analysis

N.M., K.S.O’C., B.C., J.R.I.C., Z.Q., T.D.A., T.B.B., S.Børte, J.B., A.W.C., O.K.D., M.J.Gandal, S.P.H., N.K., M.Kim, K.K., G.P., B.M.S., L.G.S., S.Steinberg, V.T., B.S.W., H-H.W., V.A., S.A., S.E.B., B.M.B., A.M.D., A.L.D., V.E-P., T.M.F., O.F., S.D.G., T.A.G., J.Grove, P.A.H., L.H., J.S.J., Y.K., M.Kubo, C.Lavebratt, M.Leber, P.H.L., S.H.M., A.Maihofer, M.M., S.A.M., S.E.M., L.M.O.L., A.F.P., T.R., P.R., D.M.R., O.B.S., C.Terao, T.E.T., J.T.R.W., W.X., J.M.K.Y., H.Y., P.P.Z., H.Z., A.D.B., G.Breen, E.S.G., F.S.G., Q.S.L., B.M-M., C.M.N., D.P., S.R., H.S., P.F.S., T.W., N.R.W., J.M.B., E.A.S.

Kevin S. O’Connell, Brandon Coombes, Jonathan R. I. Coleman, and Zhen Qiao contributed equally to this work and should be regarded as joint second authors.

## Methods

### Sample description

The meta-analysis sample comprises 57 cohorts collected in Europe, North America and Australia, totaling 41,917 BD cases and 371,549 controls of European descent (Table S1). The total effective N, equivalent to an equal number of cases and controls in each cohort (4*Ncases*Ncontrols/(Ncases+Ncontrols)), is 101,962. For 52 cohorts, individual-level genotype and phenotype data were shared with the PGC. Cohorts have been added to the PGC in five waves (PGC1^9^, PGC2^24^, PGC PsychChip, PGC3 and External Studies); all cohorts from previous PGC BD GWAS were included. The source and inclusion/exclusion criteria for cases and controls for each cohort, are described in the Supplementary Note. Cases were required to meet international consensus criteria (DSM-IV, ICD-9 or ICD-10) for a lifetime diagnosis of BD, established using structured diagnostic instruments from assessments by trained interviewers, clinician-administered checklists or medical record review. In most cohorts, controls were screened for the absence of lifetime psychiatric disorders and randomly selected from the population. For five cohorts (iPSYCH^30^, deCODE genetics^31^, Estonian Biobank^32^, Trøndelag Health Study (HUNT)^33^ and UK Biobank^34^), GWAS summary statistics for BD were shared with the PGC. In these cohorts, BD cases were ascertained using ICD codes or self-report during a nurse interview, and the majority of controls were screened for the absence of psychiatric disorders via ICD codes. Follow-up analyses included four non-European BD case-control cohorts, two from East Asia (Japan^59^ and Korea^92^), and two admixed African American cohorts^22,93^, providing a total of 5,847 cases and 65,588 controls. These BD cases were ascertained using international consensus criteria (DSM-IV)^22,93^ through psychiatric interviews (Supplementary Note).

### Genotyping, quality control and imputation

For 52 cohorts internal to the PGC, genotyping was performed following local protocols and genotypes were called using standard genotype calling softwares from commercial sources (Affymetrix and Illumina). Subsequently, standardized quality control, imputation and statistical analyses were performed centrally using RICOPILI (Rapid Imputation for COnsortias PIpeLIne) (version 2018_Nov_23.001)^94^, separately for each cohort. Briefly, the quality control parameters for retaining SNPs and subjects were: SNP missingness < 0.05 (before sample removal), subject missingness < 0.02, autosomal heterozygosity deviation (F_het_ < 0.2), SNP missingness < 0.02 (after sample removal), difference in SNP missingness between cases and controls < 0.02, SNP Hardy-Weinberg equilibrium (P > 10E-10 in psychiatric cases and P > 10E-06 in controls). Relatedness was calculated across cohorts using identity by descent and one of each pair of related individuals (pi_hat > 0.2) was excluded. Principal components (PCs) were generated using genotyped SNPs in each cohort separately using EIGENSTRAT v6.1.4^95^. Based on visual inspection of plots of PCs for each dataset (which were all of European descent according to self-report/clinical data), we excluded samples to obtain more clearly homogeneous datasets. Genotype imputation was performed using the pre-phasing/ imputation stepwise approach implemented in Eagle v2.3.5^96^ and Minimac3^97^ to the Haplotype Reference Consortium (HRC) reference panel v1.0^98^. Data on the X chromosome were available for 50 cohorts internal to the PGC and one external cohort (HUNT), and the X chromosome was imputed to the HRC reference panel in males and females separately within each cohort. The five external cohorts were processed by the collaborating research teams using comparable procedures and imputed to the HRC or a custom reference panel as appropriate. Full details of the genotyping, quality control and imputation for each of these cohorts are available in the Supplementary Note. Identical individuals between PGC cohorts and the Estonian Biobank and UK Biobank cohorts were detected using genotype-based checksums (https://personal.broadinstitute.org/sripke/share_links/zpXkV8INxUg9bayDpLToG4g58TMtjN_PGC_SCZ_w3.0718d.76) and removed from PGC cohorts.

### Genome-wide association study

For PGC cohorts, GWAS were conducted within each cohort using an additive logistic regression model in PLINK v1.90^99^, covarying for PCs 1-5 and any others as required. Association analyses of the X chromosome were conducted in males and females separately using the same procedures, with males coded as 0 or 2 for 0 or 1 copies of the reference allele. Results from males and females were then meta-analyzed within each cohort. For external cohorts, GWAS were conducted by the collaborating research teams using comparable procedures (Supplementary Note). To control test statistic inflation at SNPs with low minor allele frequency (MAF) in small cohorts, SNPs were retained only if cohort MAF was > 1% and minor allele count was > 10 in either cases or controls (whichever had smaller N). There was no evidence of stratification artifacts or uncontrolled inflation of test statistics in the results from any cohort (λ_GC_ 0.97-1.05)(Table S1). Meta-analysis of GWAS summary statistics was conducted using an inverse variance-weighted fixed effects model in METAL (version 2011-03-25)^100^ across 57 cohorts for the autosomes (41,917 BD cases and 371,549 controls) and 51 cohorts for the X chromosome (35,691 BD cases and 96,731 controls). A genome-wide significant locus was defined as the region around a SNP with P < 5E-08, with linkage disequilibrium (LD) r^2^ > 0.1, within a 3000 kilobase (kb) window. Regional association plots and forest plots of the index SNP for all genome-wide significant loci are presented in Supplementary Data 1 and 2 respectively.

### Overlap of loci with other psychiatric disorders

Genome-wide significant loci for BD were assessed for overlap with genome-wide significant loci for other psychiatric disorders, using the largest available GWAS results for major depression^61^, schizophrenia^60^, attention deficit/hyperactivity disorder^101^, post-traumatic stress disorder^102^, lifetime anxiety disorder^103^, Tourette’s Syndrome^104^, anorexia nervosa^105^, alcohol use disorder or problematic alcohol use^68^, autism spectrum disorder^106^, mood disorders^91^ and the cross-disorder GWAS of the Psychiatric Genomics Consortium^66^. The boundaries of the genome-wide significant loci were calculated in the original publications. Overlap of loci was calculated using bedtools v2.29.2^107^.

### Enrichment analyses

P values quantifying the degree of association of genes and gene sets with BD were calculated using MAGMA v1.08^37^, implemented in FUMA v1.3.6a^64,108^. Gene-based tests were performed for 19,576 genes (Bonferroni-corrected *P* value threshold = 2.55E-06). A total of 11,858 curated gene sets including at least 10 genes from MSigDB V7.0 were tested for association with BD (Bonferroni-corrected P value threshold = 4.22E-06). Competitive gene-set tests were conducted correcting for gene size, variant density and LD within and between genes. Tissue-set enrichment analyses were also performed using MAGMA implemented in FUMA, to test for enrichment of association signal in genes expressed in 54 tissue types from GTEx V8 (Bonferroni-corrected P value threshold = 9.26E-04)^64,108^.

For single-cell enrichment analyses, publicly available single-cell RNA-seq data were compiled from five studies of the adult human and mouse brain^86,109–112^. The mean expression for each gene in each cell type was computed from the single-cell expression data (if not provided). For the Zeisel dataset^109^, we used the mean expression at level 4 (39 cell types from 19 regions for the mouse nervous system). For the Saunders dataset^110^, we computed the mean expression of the different classes in each of the 9 different brain regions sampled (88 cell types in total). We filtered out any genes with non-unique names, genes not expressed in any cell types, non-protein coding genes, and, for mouse datasets, genes that had no expert curated 1:1 orthologs between mouse and human (Mouse Genome Informatics, The Jackson laboratory, version 11/22/2016, http://www.informatics.jax.org/downloads/reports/index.html#homology), resulting in 16,472 genes. Gene expression was then scaled to a total of 1 million UMIs (unique molecular identifiers) (or transcript per million (TPM)) for each cell type/tissue. Using a previously described method^38^, a metric of gene expression specificity was calculated by dividing the expression of each gene in each cell type by the total expression of that gene in all cell types, leading to values ranging from 0 to 1 for each gene (0 meaning that the gene is not expressed in that cell type and 1 meaning that all of the expression of the gene is in that cell type). We then selected the top 10% most specific genes for each cell type/tissue for enrichment analysis. MAGMA v1.08^37^ was used to test gene-set enrichment using GWAS summary statistics, covarying for gene size, gene density, mean sample size for tested SNPs per gene, the inverse of the minor allele counts per gene and the log of these metrics. We excluded any SNPs with INFO score <0.6, with MAF < 1% or with estimated odds ratio > 25 or smaller than 1/25, as well as SNPs located in the MHC region (chr6:25-34 Mb). We set a window of 35 kb upstream to 10 kb downstream of the gene coordinates to compute gene-level association statistics and used the European reference panel from the phase 3 of the 1000 genomes project as the reference population^113^. We then used MAGMA to test whether the 10% most specific genes (with an expression of at least 1 TPM or 1 UMI per million) for each cell type/tissue were associated with BD. The P value threshold for significance was P < 9.1E-03, representing a 5% false discovery rate (FDR) across datasets.

Further gene-set analyses were performed restricted to genes targeted by drugs, assessing individual drugs and grouping drugs with similar actions. This approach has been described previously^41^. Gene-level and gene-set analyses were performed in MAGMA v1.08^37^. Gene boundaries were defined using build 37 reference data from the NCBI, available on the MAGMA website (https://ctg.cncr.nl/software/magma), extended 35kb upstream and 10kb downstream to include regulatory regions outside of the transcribed region. Gene-level association statistics were defined as the aggregate of the mean and the lowest variant-level P value within the gene boundary, converted to a Z-value. Gene sets were defined comprising the targets of each drug in the Drug-Gene Interaction database DGIdb v.2^39^ and in the Psychoactive Drug Screening Database Ki DB^40^, both downloaded in June 2016^41^. Analyses were performed using competitive gene-set analyses in MAGMA. Results from the drug-set analysis were then grouped according to the Anatomical Therapeutic Chemical class of the drug^41^. Only drug classes with at least 10 valid drug gene sets within them were analyzed. Drug-class analysis was performed using enrichment curves. All drug gene sets were ranked by their association in the drug set analysis, and then for a given drug class an enrichment curve was drawn scoring a “hit” if the drug gene set was within the class, or a “miss” if it was outside of the class. The area under the curve was calculated, and a p-value for this calculated as the Wilcoxon Mann-Whitney test comparing drug gene sets within the class to drug gene sets outside of the class^41^. Multiple testing was controlled using a Bonferroni-corrected significance threshold of P < 5.60E-05 for drug-set analysis and P < 7.93E-04 for drug-class analysis, accounting for 893 drug-sets and 63 drug classes tested.

### eQTL integrative analysis

A transcriptome-wide association study (TWAS) was conducted using the precomputed gene expression weights from PsychENCODE data (1,321 brain samples)^43^, available online with the FUSION software^42^. For genes with significant *cis*-SNP heritability (13,435 genes), FUSION software (vOct 1, 2019) was used to test whether SNPs influencing gene expression are also associated with BD (Bonferroni-corrected P value threshold < 3.72E-06). For regions including a TWAS significant gene, TWAS fine-mapping of the region was conducted using FOCUS (fine-mapping of causal gene sets, v0.6.10)^44^. Regions were defined using the correlation matrix of predicted effects on gene expression around TWAS significant genes^44^. A posterior inclusion probability (PIP) was assigned to each gene for being causal for the observed TWAS association signal. Based on the PIP of each gene and a null model, whereby no gene in the region is causal for the TWAS signal, the 90%-credible gene set for each region was computed^44^.

Summary data-based Mendelian randomization (SMR) (v1.03)^45,46^ was applied to further investigate putative causal relationships between SNPs and BD via gene expression. SMR was performed using eQTL summary statistics from the eQTLGen (31,684 blood samples)^47^ and PsychENCODE^43^ consortia. SMR analysis is limited to transcripts with at least one significant *cis*-eQTL (P < 5E-08) in each dataset (15,610 in eQTLGen; 10,871 in PsychENCODE). The Bonferroni-corrected significance threshold was P < 3.20E-06 and P < 4.60E-06 for eQTLGen and PsychENCODE respectively. The significance threshold for the HEIDI test (heterogeneity in dependent instruments) was *P*_HEIDI_ ≥ 0.01^46^. While the results of TWAS and SMR indicate an association between BD and gene expression, a non-significant HEIDI test additionally indicates either a direct causal role or a pleiotropic effect of the BD-associated SNPs on gene expression.

### Complement component 4 (C4) imputation

To investigate the major histocompatibility complex (MHC; chr6:24-34 Mb on hg19), the alleles of complement component 4 genes (*C4A* and *C4B*) were imputed in 47 PGC cohorts for which individual-level genotype data were accessible, totaling 32,749 BD cases and 53,370 controls. The imputation reference panel comprised 2,530 reference haplotypes of MHC SNPs and *C4* alleles, generated using a sample of 1,265 individuals with whole-genome sequence data, from the Genomic Psychiatry cohort^114^. Briefly, imputation of *C4* as a multi-allelic variant was performed using Beagle v4.1^115,116^, using SNPs from the MHC region that were also in the haplotype reference panel. Within the Beagle pipeline, the reference panel was first converted to bref format. We used the conform-gt tool to perform strand-flipping and filtering of specific SNPs for which strand remained ambiguous. Beagle was run using default parameters with two key exceptions: we used the GRCh37 PLINK recombination map, and we set the output to include genotype probability (i.e., GP field in VCF) for correct downstream probabilistic estimation of *C4A* and *C4B* joint dosages. The output consisted of dosage estimates for each of the common *C4* structural haplotypes for each individual. The five most common structural forms of the *C4A/C4B* locus (BS, AL, AL-BS, AL-BL, and AL-AL) could be inferred with reasonably high accuracy (generally 0.70 < *r2* < 1.00). The imputed *C4* alleles were tested for association with BD in a joint logistic regression that included (i) terms for dosages of the five most common *C4* structural haplotypes (AL-BS, AL-BL, AL-AL, BS, and AL), (ii) rs13195402 genotype (top lead SNP in the MHC) and (iii) PCs as per the GWAS. The genetically regulated expression of *C4A* was predicted from the imputed *C4* alleles using a model previously described^63^. Predicted *C4A* expression was tested for association with BD in a joint logistic regression that included (i) predicted *C4A* expression, (ii) rs13195402 genotype (top lead SNP in the MHC) and (iii) PCs as per the GWAS.

### Polygenic risk scoring

PRS from our GWAS meta-analysis were tested for association with BD in individual cohorts, using a discovery GWAS where the target cohort was left out of the meta-analysis. Briefly, the GWAS results from each discovery GWAS were pruned for LD using the P value informed clumping method in PLINK v1.90^99^(r^2^ 0.1 within a 500 kb window) based on the LD structure of the HRC reference panel^98^. Subsets of SNPs were selected from the results below nine increasingly liberal P value thresholds (p_T_) (5E-08, 1E-04, 1E-03, 0.01, 0.05, 0.1, 0.2, 0.5, 1). Sets of alleles, weighted by their log odds ratios from the discovery GWAS, were summed into PRS for each individual in the target datasets, using PLINK v1.90 implemented via RICOPILI^94,99^. PRS were tested for association with BD in the target dataset using logistic regression, covarying for PCs as per the GWAS in each cohort. PRS were tested in the external cohorts by the collaborating research teams using comparable procedures. The variance explained by the PRS (R^2^) was converted to the liability scale to account for the proportion of cases in each target dataset, using a BD population prevalence of 2% and 1%^117^. The weighted average R^2^ values were calculated using the effective N for each cohort. The odds ratios for BD for individuals in the top decile of PRS compared with those in the lowest decile and middle decile were calculated in the 52 datasets internal to the PGC. To assess cross-ancestry performance, PRS generated from the meta-analysis results were tested for association with BD using similar methods in a Japanese sample^59^, a Korean sample^92^ and two admixed African American samples. Full details of the QC, imputation and analysis of these samples are in the Supplementary Note.

### LD score regression

LD Score regression (LDSC)^35^ was used to estimate the 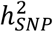 of BD from GWAS summary statistics. 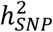 was converted to the liability scale, using a lifetime BD prevalence of 2% and 1%. LDSC bivariate genetic correlations attributable to genome-wide SNPs (*r*_g_) were estimated with 255 human diseases and traits from published GWAS and 514 GWAS of phenotypes in the UK Biobank from LD Hub^48^. Adjusting for the number of traits tested, the Bonferroni-corrected P value thresholds were P < 1.96E-04 and P < 9.73E-05 respectively.

### MiXeR

We applied causal mixture models^49,118^ to the GWAS summary statistics, using MiXeR v1.3. MiXeR provides univariate estimates of the proportion of non-null SNPs (“polygenicity”) and the variance of effect sizes of non-null SNPs (“discoverability”) in each phenotype. For each SNP, *i*, univariate MiXeR models its additive genetic effect of allele substitution, *β*_*i*_, as a point-normal mixture, 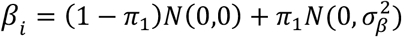, where *π*_1_ represents the proportion of non-null SNPs (‘polygenicity’) and 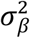 represents variance of effect sizes of non-null SNPs (‘discoverability’). Then, for each SNP, *j*, MiXeR incorporates LD information and allele frequencies for M=9,997,231 SNPs extracted from 1000 Genomes Phase3 data to estimate the expected probability distribution of the signed test statistic, 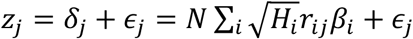, where *N* is sample size, *H*_*i*_ indicates heterozygosity of i-th SNP, *r*_*i j*_ indicates allelic correlation between i-th and j-th SNPs, and 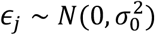 is the residual variance. Further, the three parameters, 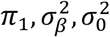, are fitted by direct maximization of the likelihood function. The optimization is based on a set of approximately 600,000 SNPs, obtained by selecting a random set of 2,000,000 SNPs with minor allele frequency of 5% or higher, followed by LD pruning procedure at LD r2=0.8 threshold. The random SNP selection and full optimization procedure are repeated 20 times to obtain mean and standard errors of model parameters. The log-likelihood figures show individual curves for each of the 20 runs, each shifted vertically so that best log-likelihood point is shown at zero ordinate. The total number of trait influencing variants is estimated as *Mπ*_1_, where M=9,997,231 gives the number of SNPs in the reference panel. MiXeR Venn diagrams report the effective number of influencing variants, ηMπ_1, where η is a fixed number, η=0.319, which gives the faction of influencing variants contributing to 90% of trait’s heritability (with rationale for this adjustment being that the remaining 68.1% of influencing variants are small and cumulatively explain only 10% of trait’s heritability). Phenotypic variance explained on average by an influencing genetic variant is calculated as 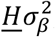, where 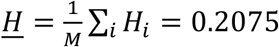 is the average heterozygosity across SNPs in the reference panel. Under the assumptions of the MiXeR model, SNP-heritability is then calculated as 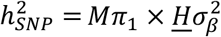

In the cross-trait analysis, MiXeR models additive genetic effects as a mixture of four components, representing null SNPs in both traits (*π*_0_); SNPs with a specific effect on the first and on the second trait (*π*_1_ and *π*_2_, respectively); and SNPs with non-zero effect on both traits (*π*_12_). In the last component, MiXeR models variance-covariance matrix as 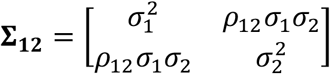 where *ρ*_12_ indicates correlation of effect sizes within the shared component, and 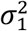 and 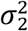 correspond to the discoverability parameter estimated in the univariate analysis of the two traits. These components are then plotted in Venn diagrams. After fitting parameters of the model, the Dice coefficient of polygenic overlap is then calculated as 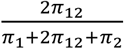, and genetic correlation is calculated as 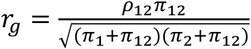.Fraction of influencing variants with concordant effect direction is calculated as twice the multivariate normal CDF at point (0, 0) for the bivariate normal distribution with zero mean and variance-covariance matrix Σ_12_. All code is available online (https://github.com/precimed/mixer).

### Mendelian randomization

Seventeen traits associated with BD in clinical or epidemiological studies were selected for Mendelian randomization (MR) to dissect their relationship with BD (Supplementary Note). Bi-directional generalized summary statistics-based MR (GSMR)^51^ analyses were performed between BD and the traits of interest using GWAS summary statistics, implemented in GCTA software (v1.93.1f beta). The instrumental variables (IVs) were selected by a clumping procedure internal to the GSMR software with parameters: *--gwas-thresh 5e-8 --clump-r2 0*.*01*. Traits with less than 10 IVs available were excluded from the GSMR analyses to avoid conducting underpowered tests^51^, resulting in 10 traits tested (Bonferroni-corrected P value threshold < 2.5E-03). The HEIDI-outlier test (heterogeneity in dependent instruments) was applied to test for horizontal pleiotropy (*P*_HEIDI_ < 0.01)^51^. For comparison, the MR analyses were also performed using the inverse variance weighted regression method, implemented via the TwoSampleMR R package, using the IVs selected by GSMR^119,120^. To further investigate horizontal pleiotropy, the MR Egger intercept test was conducted using the TwoSampleMR package^119,120^ and MR-PRESSO software was used to perform the Global Test and Distortion Test^121^.

### BD subtypes

GWAS meta-analyses were conducted for BD I (25,060 cases, 449,978 controls from 55 cohorts, effective N = 64,802) and BD II (6,781 cases, 364,075 controls from 31 cohorts, effective N = 22,560) (Table S1) using the same procedures described for the main GWAS. BD subtypes were defined based on international consensus criteria (DSM-IV, ICD-9 or ICD-10), established using structured diagnostic instruments from assessments by trained interviewers, clinician-administered checklists or medical record review. In the external biobank cohorts, BD subtypes were defined using ICD codes (Supplementary Note). LDSC^35^ was used to estimate the 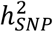 of each subtype, and the genetic correlation between the subtypes. The difference between the LDSC 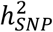 estimates for BD I and BD II was tested for deviation from 0 using the block jackknife^122^. The LDSC genetic correlation (*r*_g_) was tested for difference from 1 by calculating a chi-square statistic corresponding to the estimated *r*_g_ as [(*r*_g_ − 1)/ SE]^2^.

### Code availability

All software used is publicly available at the URLs or references cited.

