## Supplementary Note for "Genome-wide association study of over 40,000 bipolar disorder cases provides new insights into the underlying biology"

**CONTENTS**


Supplementary Figures **2**

Supplementary Note **14**

- Sample descriptions 14
- Quality control, imputation and analysis of cohorts external to the PGC 29
- Sample descriptions and polygenic risk scoring in non-European cohorts 30
- Selection of traits for Mendelian randomization 32
- Full acknowledgements 35
- Funding Sources 38
- Consortium authors and affiliations 45
- References for Supplementary Note 46

**Supplementary Figures**

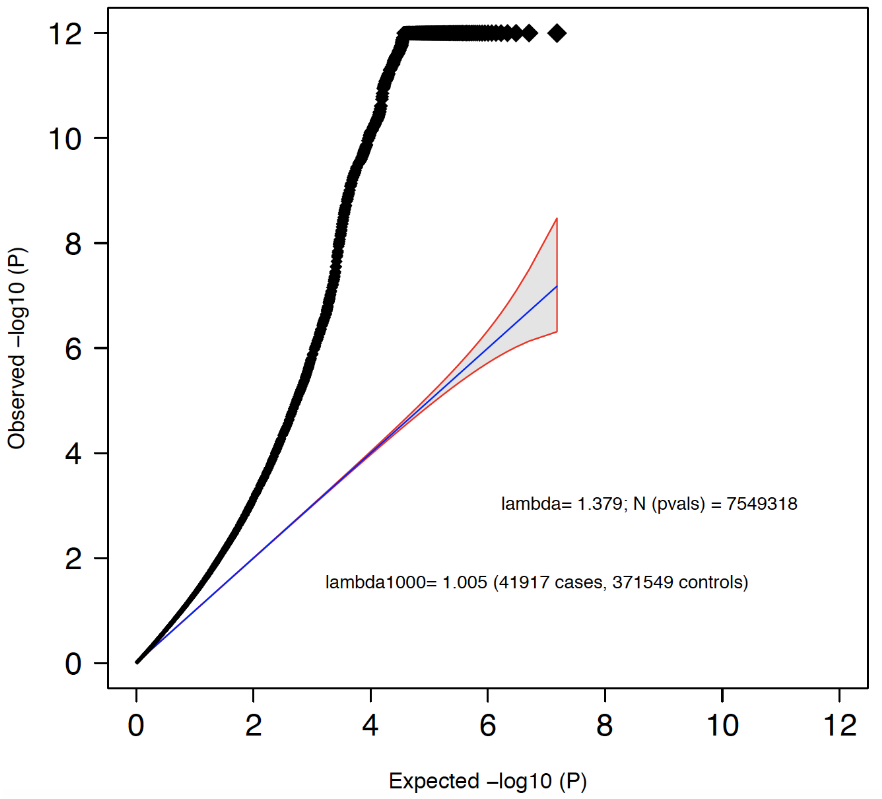


**Supplementary Figure 1: Quantile-quantile plot of association test results from genome-wide association meta-analysis of bipolar disorder**

The *y* axis is truncated at P=1E-12. SNPs plotted have a minor allele frequency >= 1% and an imputation INFO score >= 0.6. Observed results are based on an inverse variance weighted fixed effects meta-analysis of 41,917 bipolar disorder cases and 371,549 controls. P values are uncorrected and two-sided.

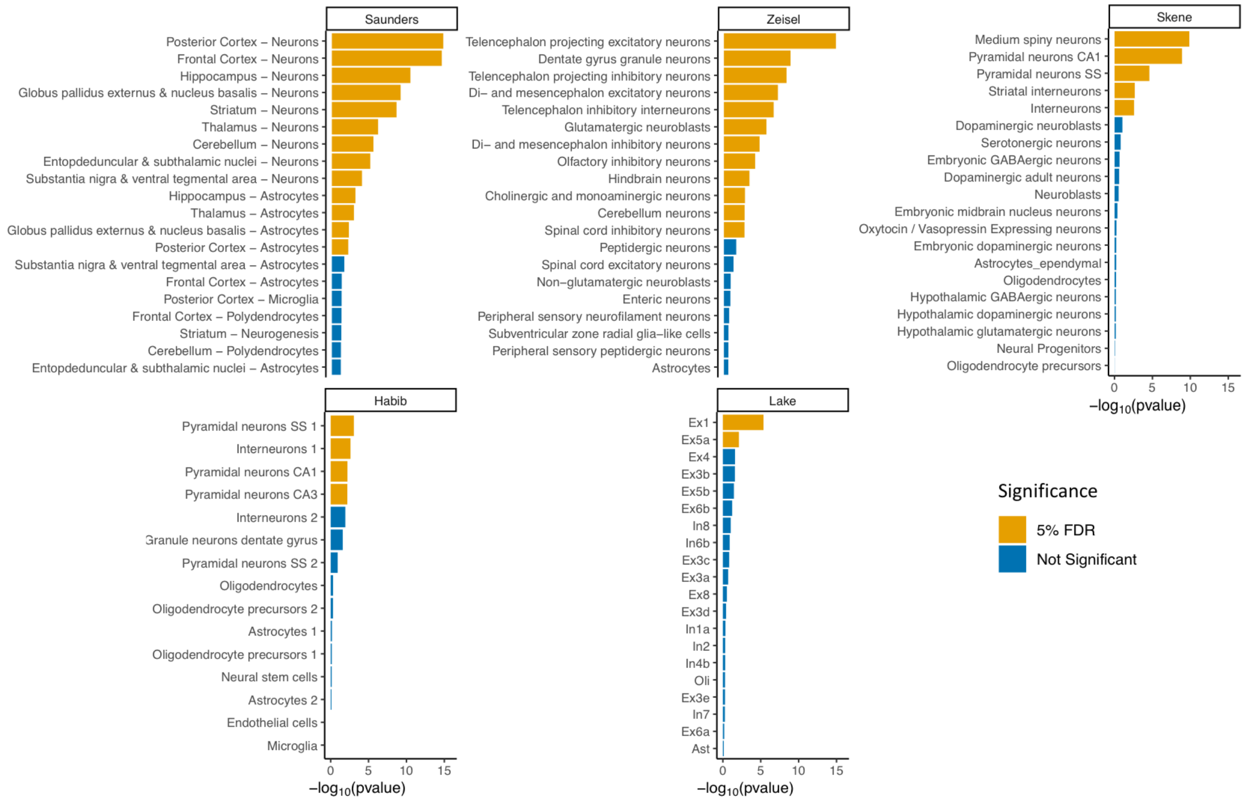


**Supplementary Figure 2: Top brain cell types enriched for bipolar disorder association signal**

The significance level (-$\log_{10}$(p value)) of MAGMA is shown for the top 20 most associated cell types in diverse datasets. The genes tested for each cell type are the top 10% of genes most specifically expressed in that cell type compared with all other cell types in the dataset. The color indicates whether the cell type is enriched for BD association signal at a 5% false discovery rate (FDR) across datasets. P values are based on a linear regression and are uncorrected and one-sided. The Zeisel, Saunders and Skene datasets are derived from mouse samples, while the Habib and Lake datasets are derived from human samples. SS - somato-sensory cortex, Ex - excitatory, In - inhibitory, Oli - oligodendrocyte. The numbers after the cell types refer to the cluster of cells with a similar gene expression profile, defined using clustering algorithms in the original publications.

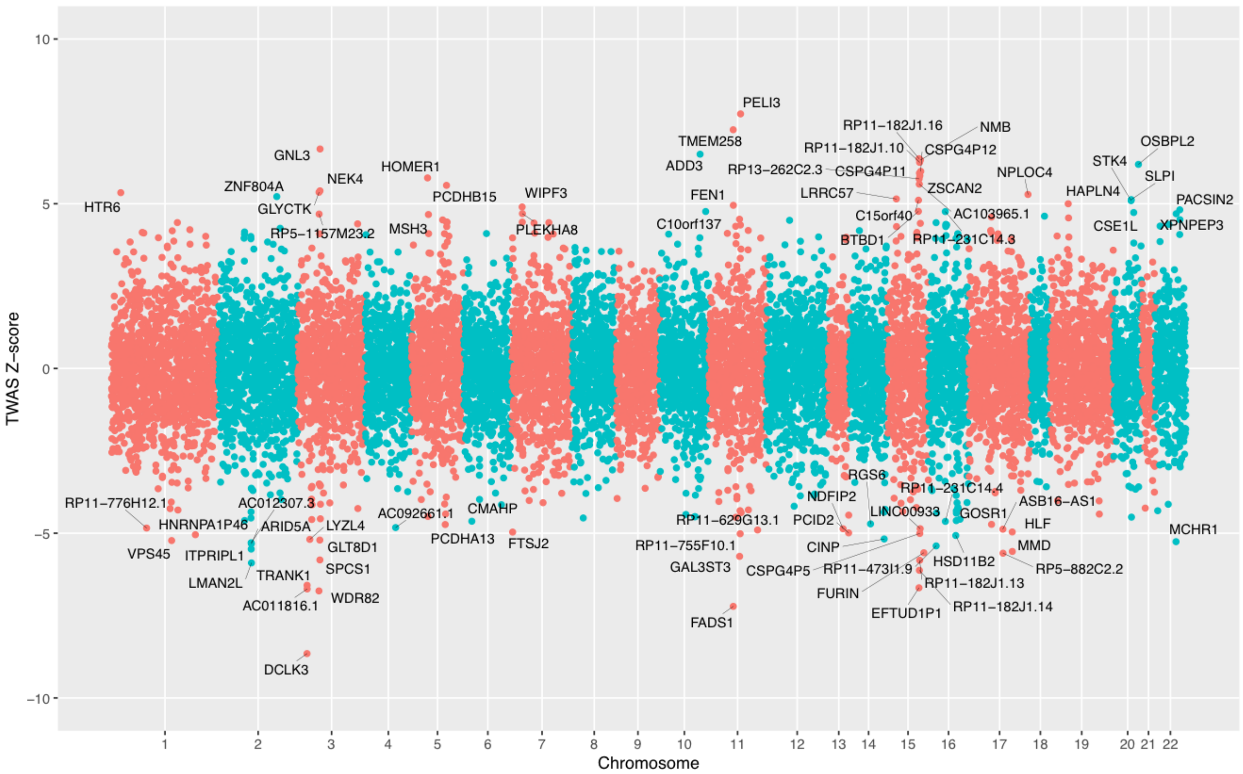


**Supplementary Figure 3: Results of transcriptome-wide association study of bipolar disorder performed using FUSION and eQTL data from the PsychENCODE Consortium**

Genes which are labeled passed the Bonferroni corrected significance threshold of P < 3.72E-06, adjusting for 13,435 genes tested. Association results are based on two-sided tests conducted using least absolute shrinkage and selection operator (lasso), bayesian sparse linear mixed model (bslmm), elastic net or best linear unbiased prediction (blup) models. TWAS Z-score - direction of effect of bipolar disorder risk alleles on predicted gene expression level.

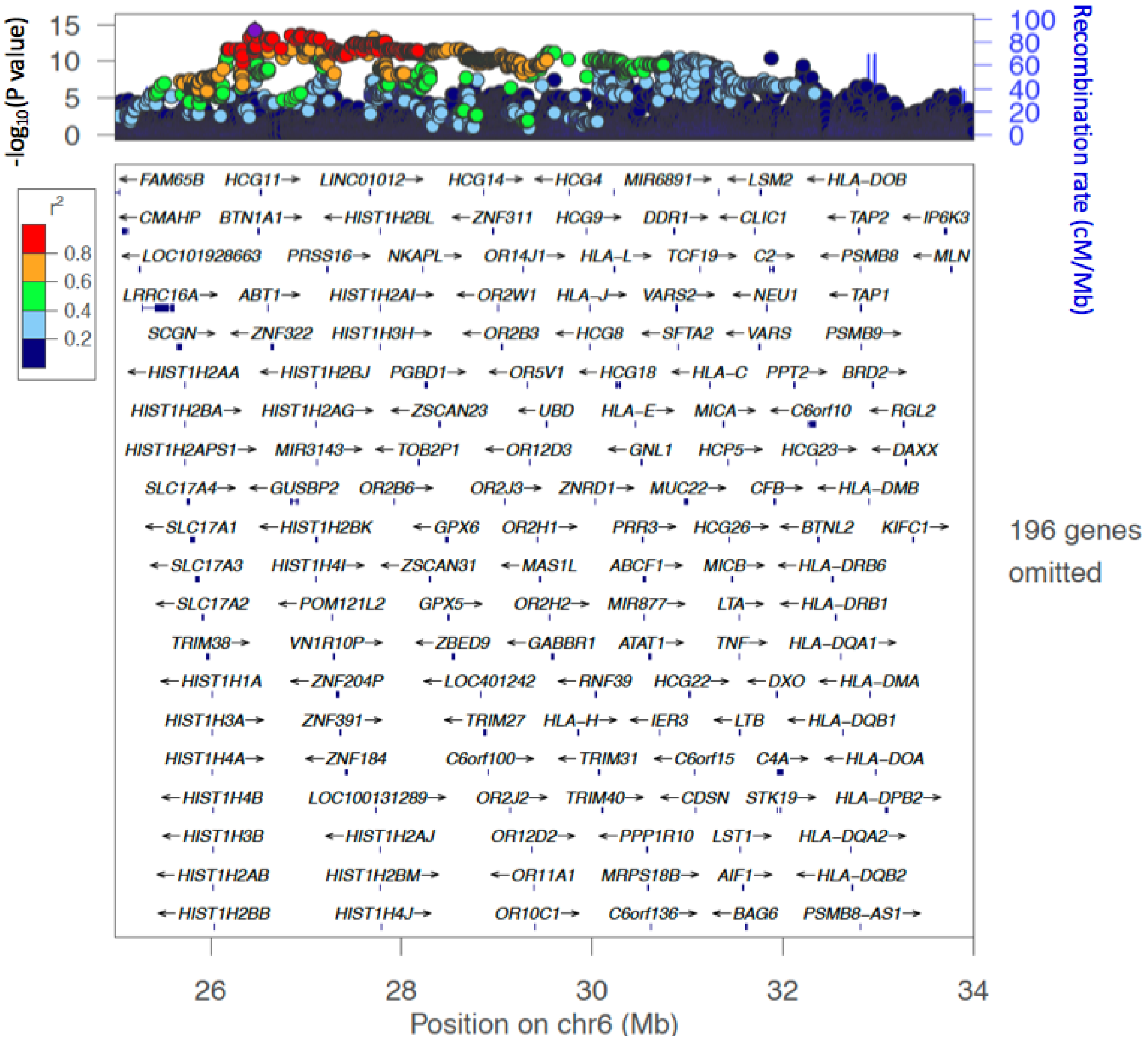


**Supplementary Figure 4: Regional plot of bipolar disorder association statistics in the extended major histocompatibility complex (MHC)**

The *x* axis shows genomic position and the *y* axis shows statistical significance as $-\log_{10}(P \text{ value})$. P values are based on an inverse variance weighted fixed effects meta-analysis of 41,917 bipolar disorder cases and 371,549 controls. P values are uncorrected and two-sided. SNPs are colored by linkage disequilibrium ($r^2$) to the top lead SNP rs13195402, which is shown as a purple diamond.

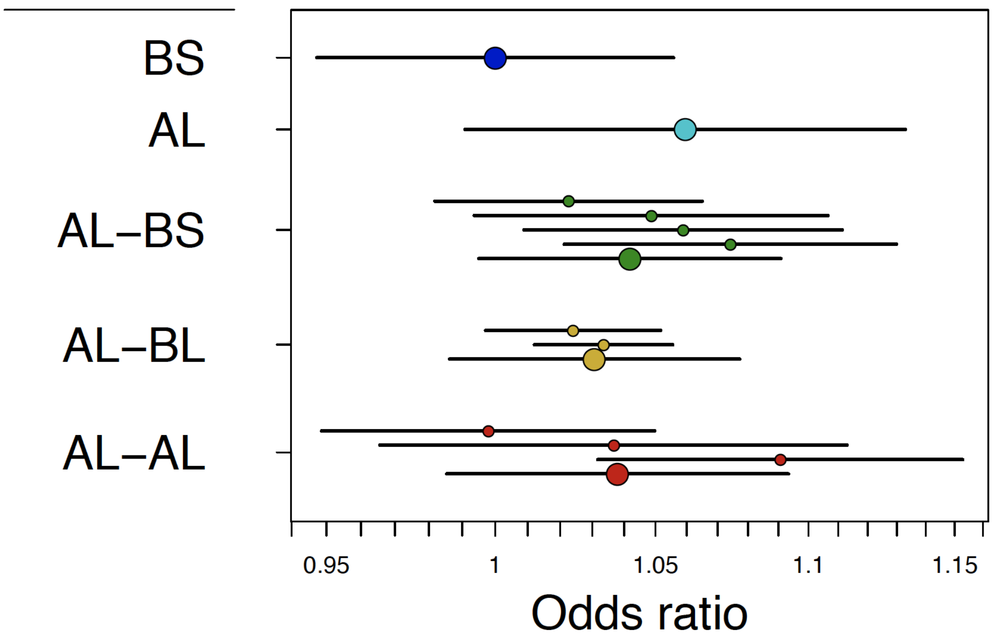


**Supplementary Figure 5: Odds ratio for bipolar disorder for the five most common *C4* locus structures, in a joint analysis that includes lead SNP rs13195402**
*C4* alleles were imputed for 32,749 bipolar disorder cases and 53,370 controls. Odds ratios are calculated relative to the BS haplotype. Error bars represent the 95% confidence interval around the effect size estimate for each allele. Because many *C4* alleles have arisen on multiple SNP haplotype backgrounds, results are shown for each specific haplogroup (small circles) as well as their combined association (large circles). There is no clear difference in bipolar disorder risk levels across these *C4* haplotypes.

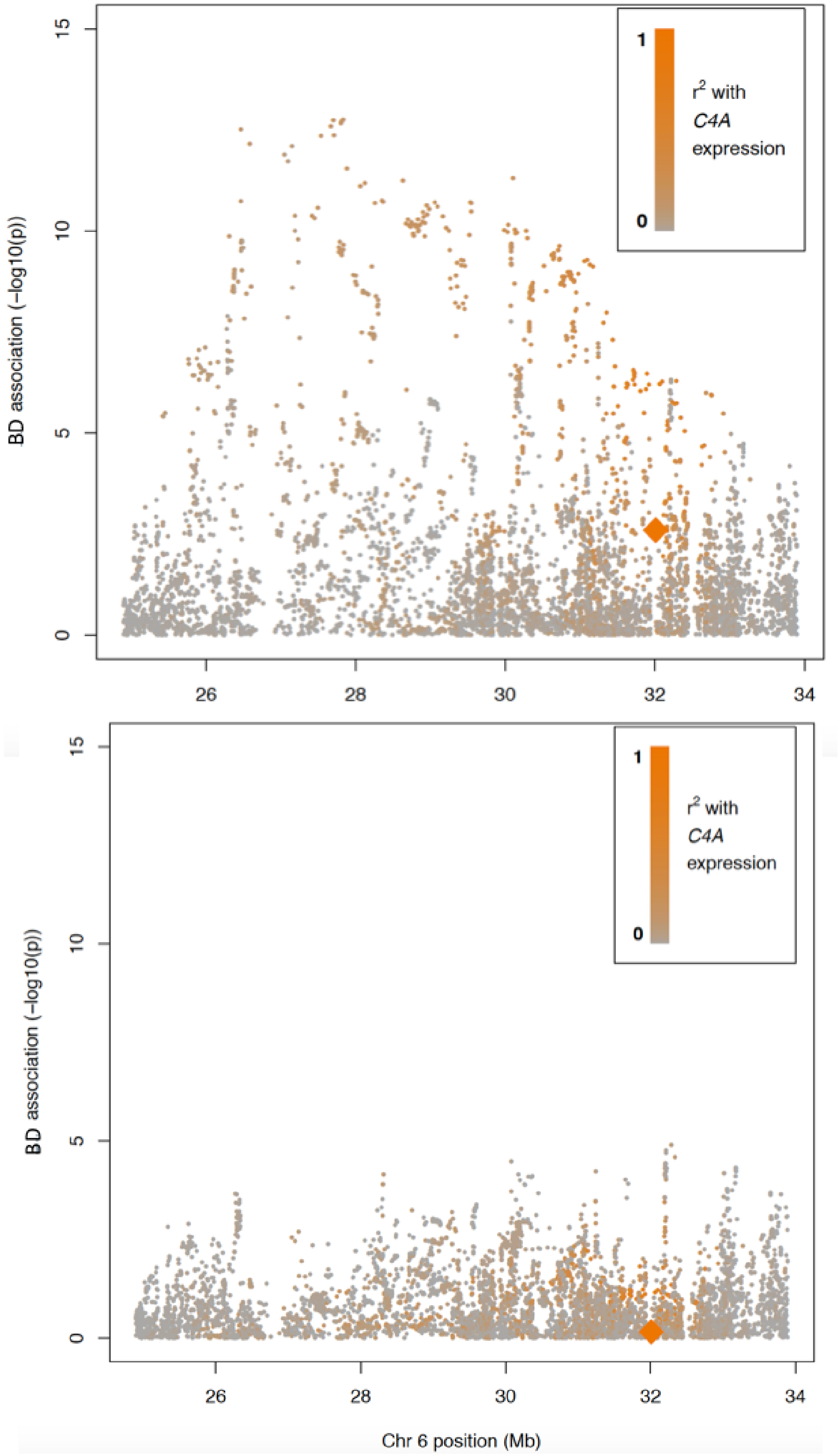


**Supplementary Figure 6: Association of bipolar disorder to chromosome 6 variation around and within the major histocompatibility complex (MHC) locus, including genetically predicted expression of *C4A***

The height of each point represents the statistical strength ($-\log_{10}$(*P* value)) of association with bipolar disorder (BD). Genetically predicted *C4A* expression is represented by the orange diamond. SNPs are colored by their level of correlation to genetically predicted *C4A* expression level. Above: unconditioned analysis. Below: Analysis conditional on rs13195402 (lead SNP in this region of chromosome 6). P values are based on logistic regression, are uncorrected and two-sided. Analyses were conducted in 32,749 bipolar disorder cases and 53,370 controls.

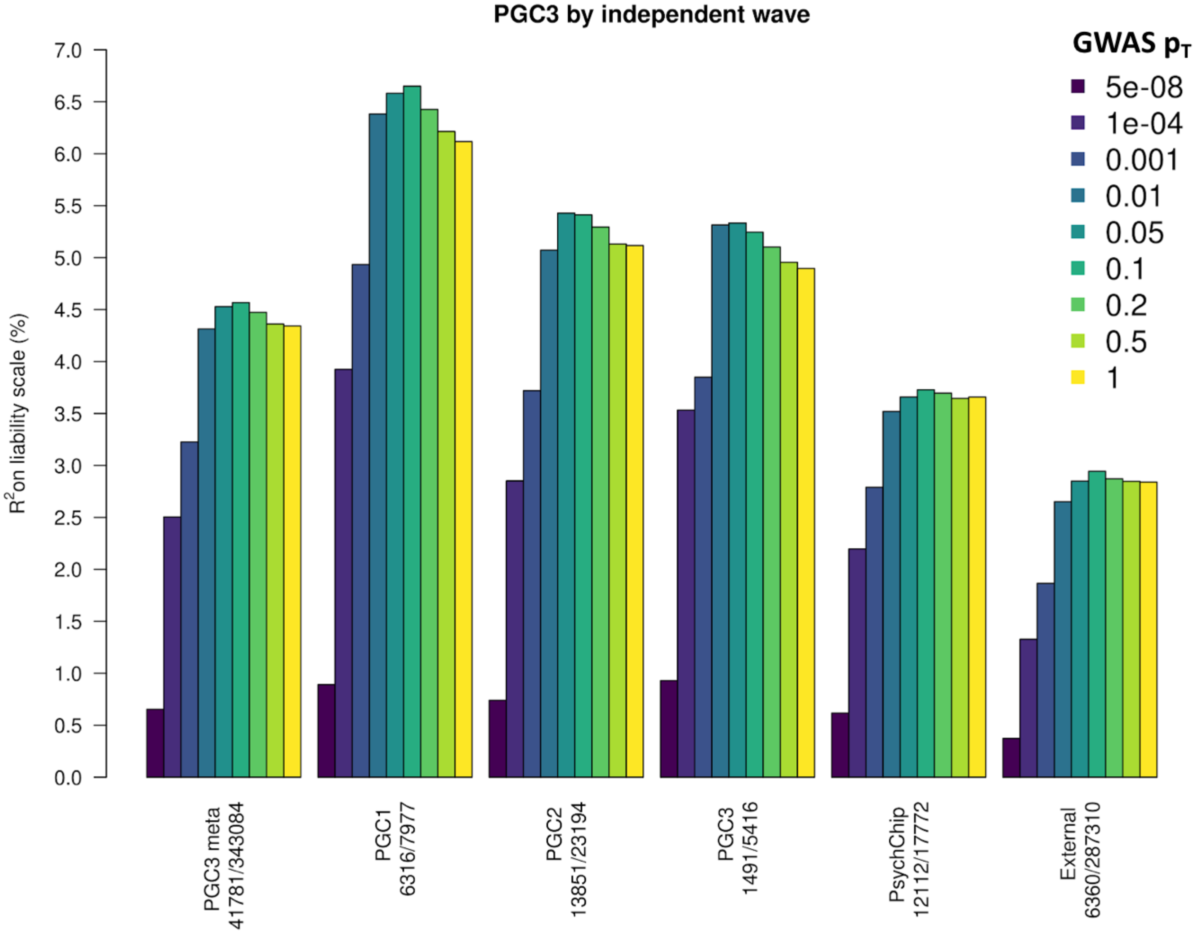


**Supplementary Figure 7: Phenotypic variance in bipolar disorder explained by polygenic risk scores per independent wave of ascertainment to the PGC**

Variance explained is presented on the liability scale, assuming a BD population prevalence of 2%. Results are based on logistic regression. The results shown are the weighted average $R^2$ values within each wave, calculated weighted by the effective N per cohort. The numbers of cases and controls are shown under the barplot for each wave from left to right. The color of the bars represents the P value threshold used to select SNPs from the discovery GWAS. The leftmost barplot ("PGC3 meta") shows the combined results of all waves (PGC1, PGC2, PGC3, PsychChip, External and follow-up [not shown in plot] datasets) and matches the European ancestry barplot in Figure 2.

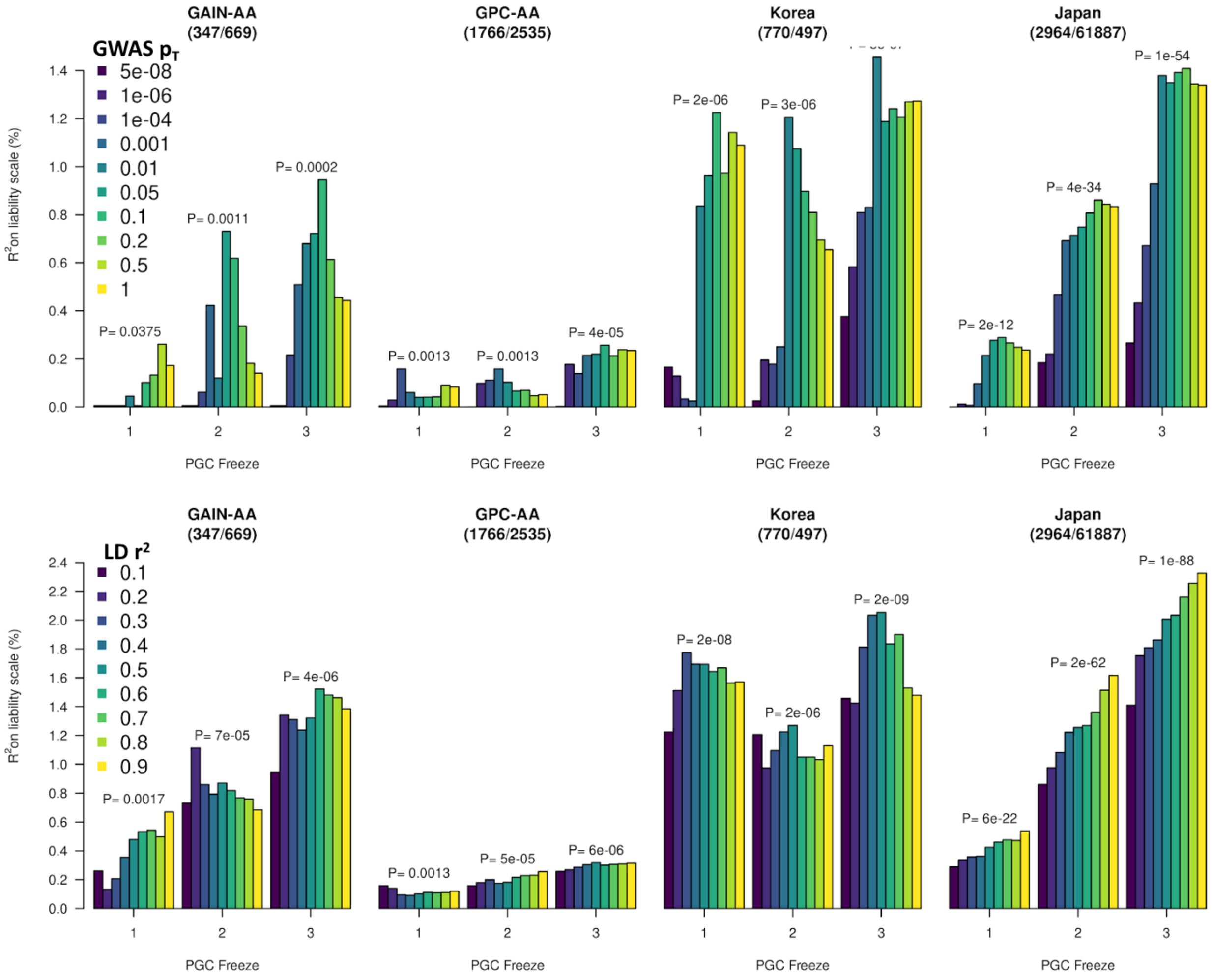


**Supplementary Figure 8: Increase of phenotypic variance in bipolar disorder explained by polygenic risk scores in independent non-European datasets, as European discovery sample size increases**

Variance explained is presented on the liability scale, assuming a BD population prevalence of 2%. The numbers of cases and controls are shown under the name of each test dataset from left to right. For each test dataset, we plot prediction performance as the PGC BD sample size has increased across freezes of the data. Top panel: Optimization of P value threshold in each dataset while setting the linkage disequilibrium (LD)-clumping threshold to 0.1. The color of the bars represents the P value threshold used to select SNPs from the discovery GWAS. Bottom panel: Optimization of LD-clumping threshold in each dataset while setting the P value threshold to the optimal for each dataset, selected in the top panel. The color of the bars represents the LD threshold used to clump SNPs from the discovery GWAS. P values for association of BD PRS with case-control status are shown for the best setting above each set of results. P values are based on logistic regression, are uncorrected and two-sided.

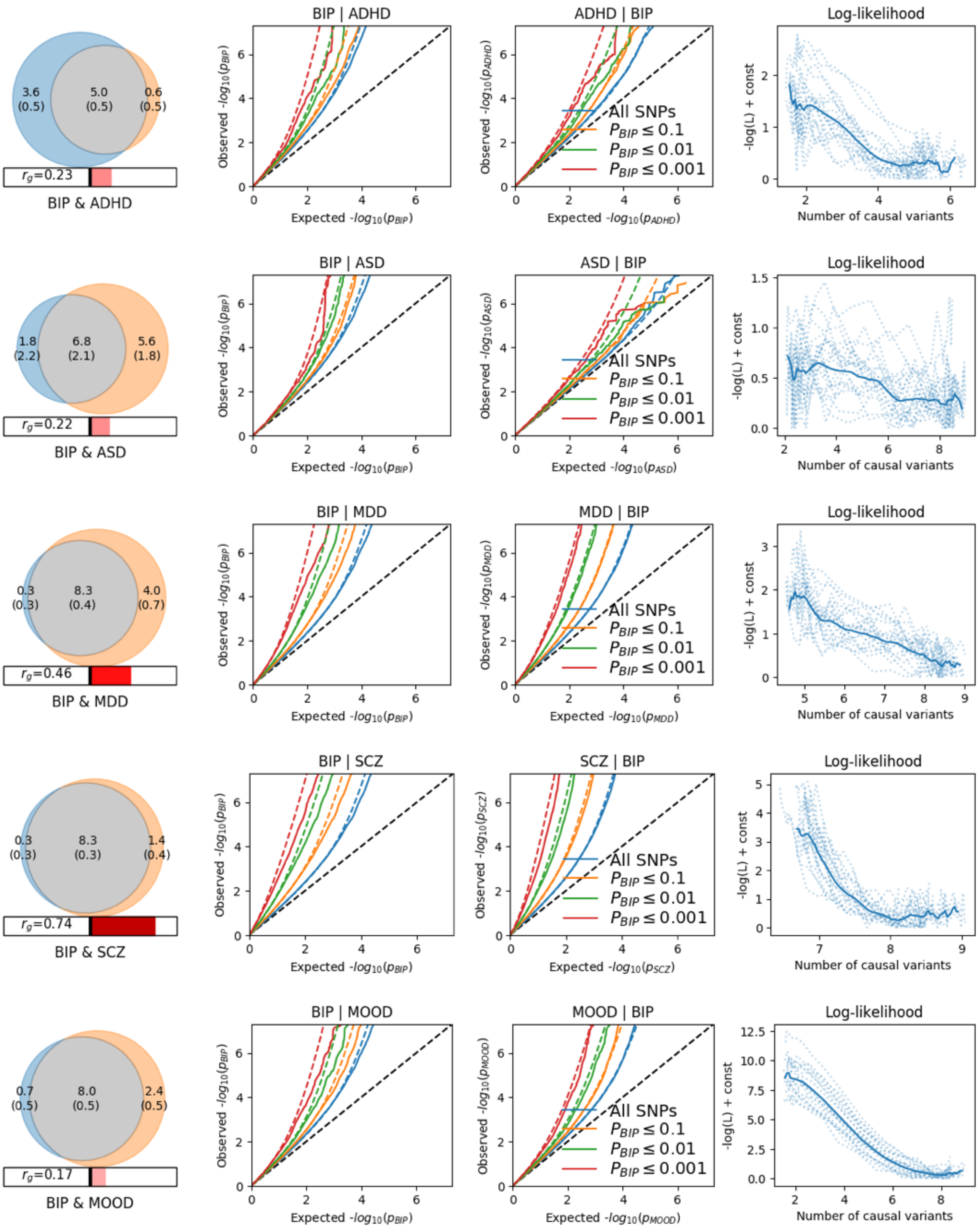
BIP & ADHD
3.6 (0.5)
5.0 (0.5)
0.6 (0.5)
$r_g$=0.23
BIP | ADHD
ADHD | BIP
Log-likelihood
BIP & ASD
1.8 (2.2)
6.8 (2.1)
5.6 (1.8)
$r_g$=0.22
BIP | ASD
ASD | BIP
BIP & MDD
0.3 (0.3)
8.3 (0.4)
4.0 (0.7)
$r_g$=0.46
BIP | MDD
MDD | BIP
BIP & SCZ
0.3 (0.3)
8.3 (0.3)
1.4 (0.4)
$r_g$=0.74
BIP | SCZ
SCZ | BIP
BIP & MOOD
0.7 (0.5)
8.0 (0.5)
2.4 (0.5)
$r_g$=0.17
BIP | MOOD
MOOD | BIP
All SNPs
$P_{BIP} \leq 0.1$
$P_{BIP} \leq 0.01$
$P_{BIP} \leq 0.001$
Observed $-log_{10}(p_{BIP})$
Expected $-log_{10}(p_{BIP})$
Observed $-log_{10}(p_{ADHD})$
Expected $-log_{10}(p_{ADHD})$
Observed $-log_{10}(p_{ASD})$
Expected $-log_{10}(p_{ASD})$
Observed $-log_{10}(p_{MDD})$
Expected $-log_{10}(p_{MDD})$
Observed $-log_{10}(p_{SCZ})$
Expected $-log_{10}(p_{SCZ})$
Observed $-log_{10}(p_{MOOD})$
Expected $-log_{10}(p_{MOOD})$
-log(L) + const
Number of causal variants

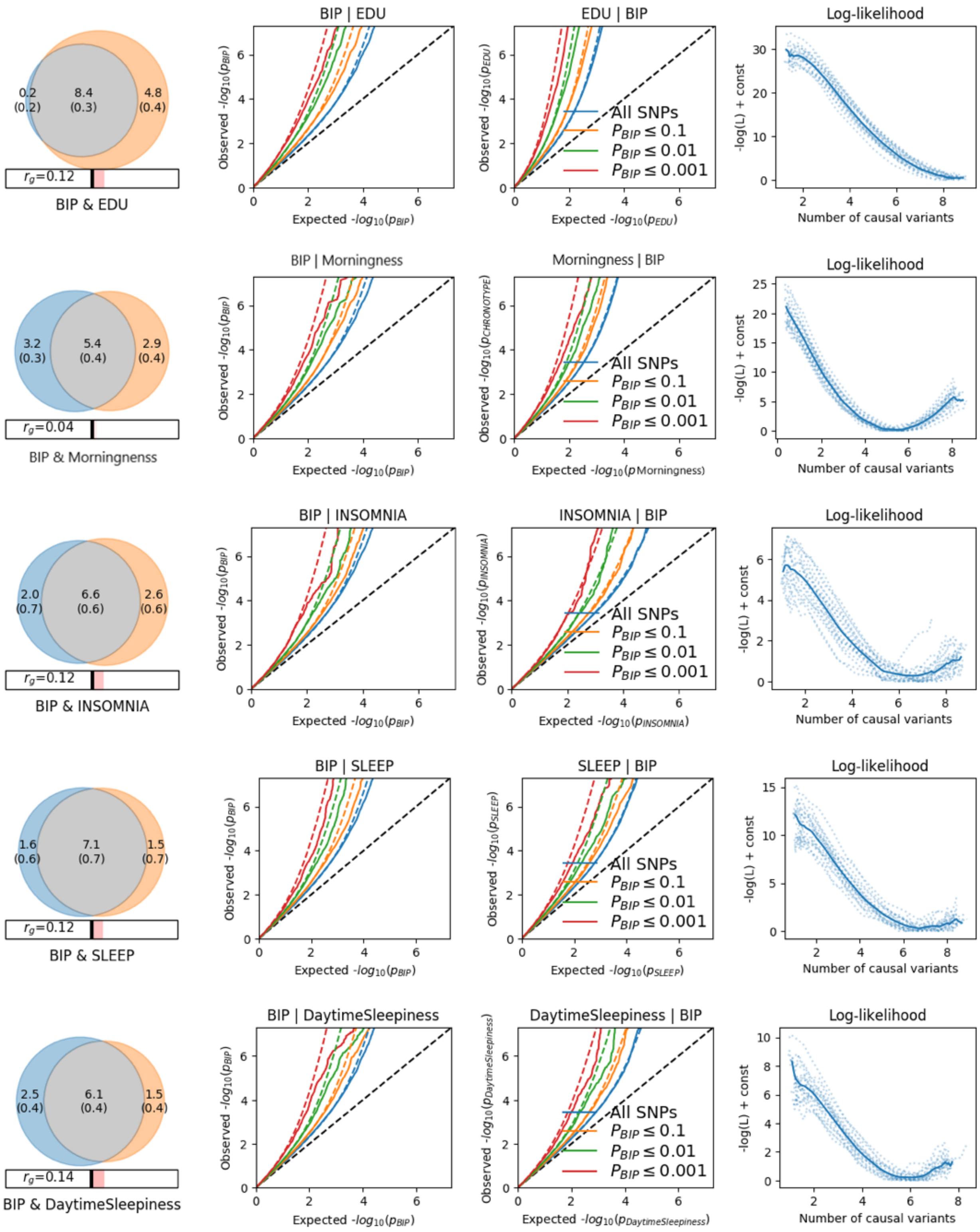
BIP & EDU
$r_g$=0.12
BIP | EDU
EDU | BIP
Log-likelihood
All SNPs
$P_{BIP} \leq 0.1$
$P_{BIP} \leq 0.01$
$P_{BIP} \leq 0.001$
BIP & Morningness
$r_g$=0.04
BIP | Morningness
Morningness | BIP
BIP & INSOMNIA
$r_g$=0.12
BIP | INSOMNIA
INSOMNIA | BIP
BIP & SLEEP
$r_g$=0.12
BIP | SLEEP
SLEEP | BIP
BIP & DaytimeSleepiness
$r_g$=0.14
BIP | DaytimeSleepiness
DaytimeSleepiness | BIP
Observed $-log_{10}(p_{BIP})$
Expected $-log_{10}(p_{BIP})$
Number of causal variants
-log(L) + const

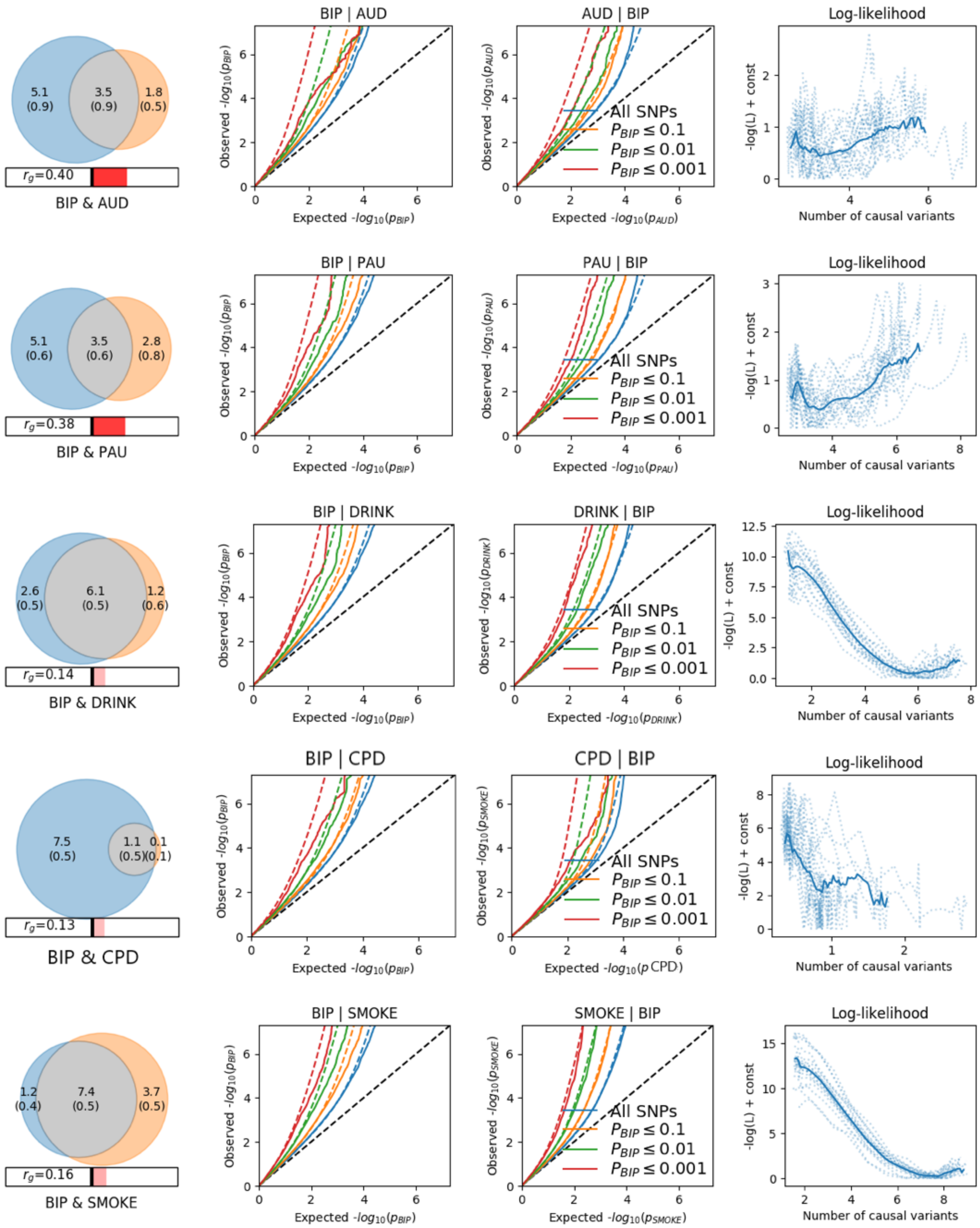
BIP & AUD
$r_g$=0.40
BIP | AUD
AUD | BIP
Log-likelihood
BIP & PAU
$r_g$=0.38
BIP | PAU
PAU | BIP
BIP & DRINK
$r_g$=0.14
BIP | DRINK
DRINK | BIP
BIP & CPD
$r_g$=0.13
BIP | CPD
CPD | BIP
BIP & SMOKE
$r_g$=0.16
BIP | SMOKE
SMOKE | BIP
All SNPs
$P_{BIP} \leq 0.1$
$P_{BIP} \leq 0.01$
$P_{BIP} \leq 0.001$
Number of causal variants
-log(L) + const

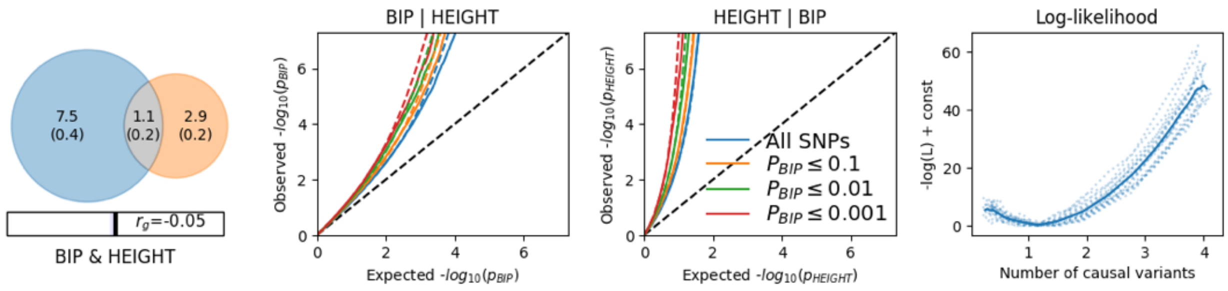


**Supplementary Figure 9: Bivariate MiXeR results comparing bipolar disorder (BD) to other traits of interest**

Venn diagrams depict the estimated number of influencing variants shared (grey) between BD and each trait of interest and unique (colors) to either of them. The number of causal variants and standard error in thousands is shown. The size of the circles reflects the polygenicity of each trait, with larger circles corresponding to greater polygenicity and vice versa. The estimated genetic correlation ($r_g$) for each pair of traits is also shown below the corresponding Venn diagram, with an accompanying scale (negative; blue shades, positive; red shades). Conditional quantile-quantile (Q-Q) plots are shown of observed versus expected −log10 P-values in the primary trait (e.g.BD) as a function of significance of association with a secondary trait (e.g. ADHD) at the level of P ≤ 0.1 (orange lines), P ≤ 0.01 (green lines), P ≤ 0.001 (red lines). P values are two-sided and based on logistic or linear regressions. Blue line indicates all SNPs. Dotted lines in blue, orange, green, and red indicate model predictions for each stratum. Black dotted line is the expected Q–Q plot under null (no SNPs associated with the phenotype). Log-likelihood curves highlight the goodness of model fit, by plotting the negative log-likelihood function (lower values correspond to better model fit) against the $\pi_{12}$ parameter (number of influencing variants shared between two traits). The remaining parameters of the model were constrained to their fitted values. The $\pi_{12}$ range on the log-likelihood plots goes from the smallest possible value $\pi_{12}=r_g*sqrt(\pi_1^u, \pi_2^u)$ that is still compatible with the estimated genetic correlation, up to the largest possible value $\pi_{12}=min(\pi_1^u, \pi_2^u)$ that corresponds to the minimum total polygenicity among the two traits. The minimum point indicates the best-fitting model estimate of the number of influencing variants shared between two traits. ASD, autism spectrum disorder. EDU, educational attainment. AUD, alcohol use disorder. PAU, problematic alcohol use. DRINK, drinks per week. CPD, cigarettes per day, . MOOD, mood instability. SLEEP, sleep duration. SMOKE, smoking initiation.

**Supplementary Note**
**Sample descriptions**
We performed a GWAS meta-analysis of 57 studies from 21 countries in Europe, North America and Australia **(Supplementary Table 1)**, totaling 41,917 cases and 371,549 controls of European descent. For 52 cohorts, raw genotype and phenotype data were shared with the Psychiatric Genomics Consortium (PGC). Cases were required to meet international consensus criteria (DSM-IV, ICD-9, or ICD-10) for a lifetime diagnosis of bipolar disorder (BD) established using structured diagnostic instruments from assessments by trained interviewers, clinician-administered checklists, or medical record review. Controls in most samples were screened for the absence of lifetime psychiatric disorders, as indicated. For five external cohorts, GWAS summary statistics for BD were shared with the PGC (iPSYCH, deCODE genetics, Estonian Biobank, HUNT and UK Biobank). Cases in these cohorts were largely defined using ICD codes ascertained from medical records. All samples in previous PGC BD GWAS papers were included, and cohorts were added to the PGC in five waves (PGC1[1], PGC2[2], PGC PsychChip, PGC3 and External Studies).

Below we describe the ascertainment and diagnosis of the participants in each individual cohort comprising this report. Most cohorts have been published on individually, and the primary report can usually be found using the PubMed identifiers provided. The lead PI of each sample warranted that their protocol was approved by their local Ethical Committee and that all participants provided written informed consent. **Supplementary Table 1** provides additional detail, including sample sizes and genotyping array. As the lifetime prevalence of BD is around 1-2%, some cohorts use controls that are not screened for BD[3,4]. The boldfaced first line for each sample indicates study PI, PubMed ID if published, country (study name), and the PGC internal tag or study identifier.

**======== PGC1 Samples ========**
**Rietschel, M; Nöthen, MM, Cichon, S | 21926972 [PGC1] | BOMA-Germany I | bip_bonn_eur**
Cases for the BOMA-Bipolar Study were ascertained from consecutive admissions to the inpatient units of the Department of Psychiatry and Psychotherapy at the University of Bonn and at the Central Institute for Mental Health in Mannheim, University of Heidelberg, Germany. DSM-IV lifetime diagnoses of bipolar I disorder were assigned using a consensus best-estimate procedure, based on all available information, including a structured interview with the SCID and SADS-L, medical records, and the family history method. In addition, the OPCRIT[5] checklist was used for the detailed polydiagnostic documentation of symptoms. Controls were ascertained from three population-based studies in Germany (PopGen, KORA, and Heinz-Nixdorf-Recall Study). The control subjects were not screened for mental illness. Study protocols were reviewed and approved in advance by Institutional Review Boards of the participating institutions. All subjects provided written informed consent.
**Corvin, A | 18711365 [PGC1] | Ireland | bip_dub1_eur**
Samples were collected as part of a larger study of the genetics of psychotic disorders in the Republic of Ireland, under protocols approved by the relevant IRBs and with written informed consent that permitted repository use. Cases were recruited from Hospitals and Community psychiatric facilities in Ireland by a psychiatrist or psychiatric nurse trained to use the SCID. Diagnosis was based on the structured interview supplemented by case note review and collateral history where available. All diagnoses were reviewed by an independent reviewer. Controls were ascertained with informed consent from the Irish GeneBank and represented blood donors who met the same ethnicity criteria as cases. Controls were not specifically screened for psychiatric illness.
**Blackwood, D | 18711365 [PGC1] | Edinburgh, UK | bip_edi1_eur**
This sample comprised Caucasian individuals contacted through the inpatient and outpatient services of hospitals in South East Scotland. A BD-I diagnosis was based on an interview with the patient using the

SADS-L supplemented by case note review and frequently by information from medical staff, relatives and caregivers. Final diagnoses, based on DSM-IV criteria were reached by consensus between two trained psychiatrists. Ethnically-matched controls from the same region were recruited through the South of Scotland Blood Transfusion Service. Controls were not directly screened to exclude those with a personal or family history of psychiatric illness. The study was approved by the Multi-Centre Research Ethics Committee for Scotland and patients gave written informed consent for the collection of DNA samples for use in genetic studies.

**Kelsoe, J | 21926972 [PGC1] | USA (GAIN) | bip_gain_eur**

*Genetic Association Information Network (GAIN)/ The Bipolar Genome Study (BiGS)* The BD sample was collected under the auspices of the NIMH Genetics Initiative for BD (http://zork.wustl.edu/nimh/), genotyped as part of GAIN and analyzed as part of a larger GWAS conducted by the BiGS consortium. Approximately half of the GAIN sample was collected as multiplex families or sib pair families (waves 1-4), the remainder were collected as individual cases (wave 5). Subjects were ascertained at 11 sites: Indiana University, John Hopkins University, the NIMH Intramural Research Program, Washington University at St. Louis, University of Pennsylvania, University of Chicago, Rush Medical School, University of Iowa, University of California, San Diego, University of California, San Francisco, and University of Michigan. All investigations were carried out after the review of protocols by the IRB at each participating institution. At all sites, potential cases were identified from screening admissions to local treatment facilities and through publicity programs or advocacy groups. Potential cases were evaluated using the DIGS[6], FIGS[7], and information from relatives and medical records. All information was reviewed through a best estimate diagnostic procedure by two independent and non-interviewing clinicians and a consensus best-estimate diagnosis was reached. In the event of a disagreement, a third review was done to break the tie. Controls were from the NIMH Genetic Repository sample obtained by Dr. P. Gejman through a contract to Knowledge Networks, Inc. Only individuals with complete or near-complete psychiatric questionnaire data who did not fulfill diagnostic criteria for major depression and denied a history of psychosis or BD were included as controls for BiGS analyses. Controls were matched for gender and ethnicity to the cases.

**Scott, L; Myer, RM; Boehnke, M | 19416921 [PGC1] | Michigan, USA (Pritzker and NIMH) | bip_mich_eur**

The Pritzker Neuropsychiatric Disorders Research Consortium (NIMH/Pritzker) case and control samples were from the NIMH Genetics Initiative Genetics Initiative Repository. Cases were diagnosed according to DMS-III or DSM-IV criteria using diagnostic interviews and/or medical record review. Cases with low confidence diagnoses were excluded. From each wave 1-5 available non-Ashkenazi European-origin family, two BD1 siblings were included when possible and the proband was preferentially included if available (n=946 individuals in 473 sibling pairs); otherwise a single BD1 case was included (n=184). The bipolar sibling pairs were retained within the NIMH/Pritzker sample when individuals in more than one study were uniquely assigned to a study set. Controls had non-Ashkenazi European-origin, were aged 20-70 years and reported no diagnosis with or treatment for BD or schizophrenia, and that they had not heard voices that others could not hear. Individuals with suspected major depression were excluded based on answers to questions related to depressive mood. NIMH controls were further selected as the best match(es) to NIMH cases based on self-reported ancestry.

**Sklar, P; Smoller, J | 18317468 [PGC1] | USA (STEP1) | bip_stp1_eur**

The Systematic Treatment Enhancement Program for Bipolar Disorder (STEP-BD) was a seven-site, national U.S., longitudinal cohort study designed to examine the effectiveness of treatments and their impact on the course of BD that enrolled 4,361 participants who met DSM-IV criteria for BD1, BD2, bipolar not otherwise specified (NOS), schizoaffective manic or bipolar type, or cyclothymic disorder based on diagnostic interviews. From the parent study, 2,089 individuals who were over 18 years of age with BD1 and BD2 diagnoses consented to the collection of blood samples for DNA. BD samples with a consensus diagnosis of BD1 were selected for inclusion in STEP1. Two groups of controls samples from the NIMH repository were used. One comprised DNA samples derived from US Caucasian anonymous cord blood donors. The second were controls who completed the online self-administered psychiatric screen and were ascertained as described above, by Knowledge Networks Inc. For the second sample of controls only those without a history of schizophrenia, psychosis, BD or major depression with functional impairment were used.

**Sklar, P; Smoller, J | 18711365 [PGC1] | USA (STEP2) | bip_stp2_eur**

The STEP2 sample included BD-1 and BD-2 samples from the STEP-BD study described above along with BD-2 subjects from UCL study also described above. The controls samples for this study were from the NIMH repository as described above for the STEP1 study.

**Andreassen, OA | PMID:21926972 [PGC1], PMID:20451256 | Norway (TOP) | bip_top7_eur**

In the TOP study (Tematisk omrade psykoser), cases of European ancestry, born in Norway, were recruited from psychiatric hospitals in the Oslo region. Patients were diagnosed according to the SCID[8] and further ascertainment details have been reported. Healthy control subjects were randomly selected from statistical records of persons from the same catchment area as the patient groups. The control subjects were screened by interview and with the Primary Care Evaluation of Mental Disorders (PRIME-MD)[9]. None of the control subjects had a history of moderate/severe head injury, neurological disorder, mental retardation or an age outside the age range of 18-60 years. Healthy subjects were excluded if they or any of their close relatives had a lifetime history of a severe psychiatric disorder. All participants provided written informed consent and the human subjects protocol was approved by the Norwegian Scientific-Ethical Committee and the Norwegian Data Protection Agency.

**McQuillin, A; Gurling, H | 18317468 [PGC1] | UCL (University College London), London, UK | bip_uclo_eur**

The UCL sample comprised Caucasian individuals who were ascertained and received clinical diagnoses of bipolar 1 disorder according to UK National Health Service (NHS) psychiatrists at interview using the categories of the International Classification of Disease version 10. In addition bipolar subjects were included only if both parents were of English, Irish, Welsh or Scottish descent and if three out of four grandparents were of the same descent. All volunteers read an information sheet approved by the Metropolitan Medical Research Ethics Committee who also approved the project for all NHS hospitals. Written informed consent was obtained from each volunteer. The UCL control subjects were recruited from London branches of the National Blood Service, from local NHS family doctor clinics and from university student volunteers. All control subjects were interviewed with the SADS-L to exclude all psychiatric disorders.

**Craddock, N, Jones, I, Jones, L | 17554300 | WTCCC | bip_wtcc_eur_sr-qc**

Cases were all over the age of 17 yr, living in the UK and of European descent. Recruitment was undertaken throughout the UK and included individuals who had been in contact with mental health services and had a lifetime history of high mood. After providing written informed consent, participants were interviewed by a trained psychologist or psychiatrist using a semi-structured lifetime diagnostic psychiatric interview (Schedules for Clinical Assessment in Neuropsychiatry) and available psychiatric medical records were reviewed. Using all available data, best-estimate life-time diagnoses were made according to the RDC[12]. In the current study we included cases with a lifetime diagnosis of RDC bipolar 1 disorder, bipolar 2 disorder or schizo-affective disorder, bipolar type.

Controls were recruited from two sources: the 1958 Birth Cohort study and the UK Blood Service (blood donors) and were not screened for history of mental illness.

All cases and controls were recruited under protocols approved by the appropriate IRBs. All subjects gave written informed consent.

**======== PGC2 Samples ========**

**Adolfsson, R | Not published | Umeå, Sweden | bip_ume4_eur**
Clinical characterization of the patients included the Mini-International Neuropsychiatric Interview (MINI[10]), the Diagnostic Interview for Genetic Studies (DIGS[6]), the Family Interview for Genetic Studies (FIGS[7]) and the Schedules for Clinical Assessment in Neuropsychiatry (SCAN)[11]. The final diagnoses were made according to the DSM-IV-TR and determined by consensus of 2 research psychiatrists. The unrelated Swedish control individuals, consisting of a large population-based sample representative of the general population of the region, were randomly selected from the 'Betula study'.
**Alda, M; Smoller, J | Not published | Nova Scotia, Canada; I2B2 controls | bip_hal2_eur**
The case samples were recruited from patients longitudinally followed at specialty mood disorders clinics in Halifax and Ottawa (Canada). Cases were interviewed in a blind fashion with the Schedule of Affective Disorders and Schizophrenia-Lifetime version (SADS-L)[12] and consensus diagnoses were made according to DSM-IV[13] and Research Diagnostic Criteria (RDC)[14]. Protocols and procedures were approved by the local Ethics Committees and written informed consent was obtained from all patients before participation in the study. Control subjects were drawn from the I2B2 (Informatics for Integrating Biology and the Bedside) project[15]. The study consists of de-identified healthy individuals recruited from a healthcare system in the Boston, MA, US area. The de-identification process meant that the Massachusetts General Hospital Institutional Review Board elected to waive the requirement of seeking informed consent as detailed by US Code of Federal Regulations, Title 45, Part 46, Section 116 (46.116).
**Andreassen, OA | Not published | Norway (TOP) | bip_top8_eur**
The TOP8 bipolar disorder cases and controls were ascertained in the same way as the bip_top7_eur (TOP7) samples described above, and recruited from hospitals across Norway.
**Biernacka, JM; Frye, MA | 27769005 | Mayo Clinic, USA | bip_may1_eur**
Bipolar cases were drawn from the Mayo Clinic Bipolar Biobank[16]. Enrolment sites included Mayo Clinic, Rochester, Minnesota; Lindner Center of HOPE/University of Cincinnati College of Medicine, Cincinnati, Ohio; and the University of Minnesota, Minneapolis, Minnesota. Enrolment at each site was approved by the local Institutional Review Board, and all participants consented to use of their data for future genetic studies. Participants were identified through routine clinical appointments, from in-patients admitted in mood disorder units, and recruitment advertising. Participants were required to be between 18 and 80 years old and be able to speak English, provide informed consent, and have DSM-IV-TR diagnostic confirmation of type 1 or 2 bipolar disorder or schizoaffective bipolar disorder as determined using the SCID. Controls were selected from the Mayo Clinic Biobank[17]. Potential controls with ICD9 codes for bipolar disorder, schizophrenia or related diagnoses in their electronic medical record were excluded.
**Breen, G; Vincent, JB | 24387768; 19416921; 21926972 [PGC1] |London, UK; Toronto, Canada [BACC] |bip_bac1_eur**
The total case/control cohort (N=1922) includes 871 subjects from Toronto, Canada (N=431 cases (160 male; 271 female); N=440 controls (176 male; 264 female)), 1051 subjects from London, UK (N=538 cases (180 male; 358 female); N=513 controls (192 male; 321 female)). A summary of mean and median age at interview, age of onset (AOO), diagnostic subtypes (BD 1 versus BD 2), presence of psychotic symptoms, suicide attempt and family history of psychiatric disorders has been provided previously for both the Toronto and London cohorts[18]. From the Toronto site (Centre for Addiction & Mental Health (CAMH)), BD individuals and unrelated healthy controls matched for age, gender and ethnicity were recruited. Inclusion criteria for patients: a) diagnosed with DSMIV/ICD 10 BD 1 or 2; b) 18 years old or over; c) Caucasian, of Northern and Western European origin, and three out of four grandparents also N.W. European Caucasian. Exclusion criteria include: a) Use of intravenous drugs; b) Evidence of intellectual disability; c) Related to an individual already in the study; d) Manias that only ever occurred in relation to or resulting from alcohol or substance abuse/dependence, or medical illness; e) Manias resulting from non-psychotropic substance usage. The SCAN interview (Schedule for Clinical Assessments in Neuropsychiatry) was used for subject assessment[19]. Using the SCAN interview along with case note

review, each case was assigned DSM-IV and ICD 10 diagnoses by two independent diagnosticians, according to lifetime consensus best-estimate diagnosis. Lifetime occurrence of psychiatric symptoms was also recorded using the OPCRIT checklist, modified for use with mood disorders. Similar methods and criteria were also used to collect a sample of 538 BD cases and 513 controls for the London cohort (King's College London; KCL)[20].

Both studies were approved by respective institutional research ethics committees (the CAMH Research Ethics Board (REB) in Toronto, and the College Research Ethics Committee (CREC) at KCL), and informed written consent was obtained from all participants. GWAS results have previously been published for the entire KCL/CAMH cohort[21].

**Rietschel, M; Nöthen, MM; Schulze, TG; Reif, A; Forstner, AJ | 24618891 | BOMA-Germany II | bip_bmg2_eur**

Cases were recruited from consecutive admissions to psychiatric in-patient units at the University Hospital Würzburg. All cases received a lifetime diagnosis of BD according to the DSM-IV criteria using a consensus best-estimate procedure based on all available information, including semi-structured diagnostic interviews using the Association for Methodology and Documentation in Psychiatry[22], medical records and the family history method. In addition, the OPCRIT system was used for the detailed polydiagnostic documentation of symptoms.

Control subjects were ascertained from the population-based Heinz Nixdorf Recall (HNR) Study[23]. The controls were not screened for a history of mental illness. Study protocols were reviewed and approved in advance by Institutional Review Boards of the participating institutions. All subjects provided written informed consent.

**Rietschel, M; Nöthen, MM; Schulze, TG; Bauer, M; Forstner, AJ; Müller-Myhsok, B | 24618891 | BOMA-Germany III | bip_bmg3_eur**[24]

Cases were recruited at the Central Institute of Mental Health in Mannheim, University of Heidelberg, and other collaborating psychiatric hospitals in Germany. All cases received a lifetime diagnosis of BD according to the DSM-IV criteria using a consensus best-estimate procedure based on all available information including structured diagnostic interviews using the AMDP, Composite International Diagnostic Screener (CID-S)[25], SADS-L and/or SCID, medical records, and the family history method. In addition, the OPCRIT system was used for the detailed polydiagnostic documentation of symptoms.

Controls were selected randomly from a Munich-based community sample and recruited at the Max-Planck Institute of Psychiatry. They were screened for the presence of anxiety and mood disorders using the CID-S. Only individuals without mood and anxiety disorders were collected as controls. Study protocols were reviewed and approved in advance by Institutional Review Boards of the participating institutions. All subjects provided written informed consent.

**Hauser, J; Lissowska, J; Forstner, AJ | 24618891 | BOMA-Poland | bip_bmpo_eur**

Cases were recruited at the Department of Psychiatry, Poznan University of Medical Sciences, Poznan, Poland. All cases received a lifetime diagnosis of BD according to the DSM-IV criteria on the basis of a consensus best-estimate procedure and structured diagnostic interviews using the SCID. Controls were drawn from a population-based case-control sample recruited by the Cancer-Center and Institute of Oncology, Warsaw, Poland and a hospital-based case-control sample recruited by the Nofer Institute of Occupational Medicine, Lodz, Poland. The Polish controls were produced by the International Agency for Research on Cancer (IARC) and the Centre National de Génotypage (CNG) GWAS Initiative for a study of upper aerodigestive tract cancers. The controls were not screened for a history of mental illness. Study protocols were reviewed and approved in advance by Institutional Review Boards of the participating institutions. All subjects provided written informed consent.

**Rietschel, M; Nöthen, MM; Rivas, F; Mayoral, F; Kogevinas, M; others | 24618891 | BOMA-Spain | bip_bmsp_eur**

Cases were recruited at the mental health departments of the following five centers in Andalusia, Spain:

University Hospital Reina Sofia of Córdoba, Provincial Hospital of Jaen; Hospital of Jerez de la Frontera (Cádiz); Hospital of Puerto Real (Cádiz); Hospital Punta Europa of Algeciras (Cádiz); and Hospital Universitario San Cecilio (Granada). Diagnostic assessment was performed using the SADS-L; the OPCRIT; a review of medical records; and interviews with first and/or second degree family members using the Family Informant Schedule and Criteria (FISC)[26]. Consensus best estimate BD diagnoses were assigned by two or more independent senior psychiatrists and/or psychologists, and according to the RDC, and the DSM-IV. Controls were Spanish subjects drawn from a cohort of individuals recruited in the framework of the European Community Respiratory Health Survey (ECRHS, http://www.ecrhs.org/). The controls were not screened for a history of mental illness. Study protocols were reviewed and approved in advance by Institutional Review Boards of the participating institutions. All subjects provided written informed consent.

**Fullerton, J.M.; Mitchell, P.B.; Schofield, P.R.; Martin N.G.; Cichon, S. | 24618891 | BOMA-Australia | bip_bmau_eur**

Cases were recruited at the Mood Disorder Unit, Prince of Wales Hospital in Sydney. All cases received a lifetime diagnosis of BD according to the DSM-IV criteria on the basis of a consensus best-estimate procedure[19] and structured diagnostic interviews using the DIGS, FIGS, and the SCID. Controls were parents of unselected adolescent twins from the Brisbane Longitudinal Twin Study. The controls were not screened for a history of mental illness. Study protocols were reviewed and approved in advance by Institutional Review Boards of the participating institutions. All subjects provided written informed consent.

**Grigoroiu-Serbanescu, M; Nöthen, MM | 21353194 | BOMA-Romania | bip_rom3_eur**

Cases were recruited from consecutive admissions to the Obregia Clinical Psychiatric Hospital, Bucharest, Romania. Patients were administered the DIGS[27] and FIGS[7] interviews. Information was also obtained from medical records and close relatives. The diagnosis of BP-I was assigned according to DSM-IV criteria using the best estimate procedure. All patients had at least two hospitalized illness episodes. Population-based controls were evaluated using the DIGS to exclude a lifetime history of major affective disorders, schizophrenia, schizoaffective disorders, and other psychoses, obsessive-compulsive disorder, eating disorders, and alcohol or drug addiction.

**Kelsoe, J; Sklar, P; Smoller, J | [PGC1 Replication] | USA (FAT2; FaST, BiGS, TGEN) | bip_fat2_eur**

Cases were collected from individuals at the 11 U.S. sites described for the GAIN sample. Eligible participants were age 18 or older meeting DSM-IV criteria for BD-I or BD-II by consensus diagnosis based on interviews with the Affective Disorders Evaluation (ADE) and MINI. All participants provided written informed consent and the study protocol was approved by IRBs at each site. Collection of phenotypic data and DNA samples were supported by NIMH grants MH063445 (JW Smoller); MH067288 (PI: P Sklar), MH63420 (PI: V Nimgaonkar) and MH078151, MH92758 (PI: J. Kelsoe). The control samples were NIMH controls that were using the methods described in that section. The case and control samples were independent of those included in the GAIN sample.

**Kirov, G | 25055870 | Bulgarian trios | bip_butr_eur**

All cases were recruited in Bulgaria from psychiatric inpatient and outpatient services. Each proband had a history of hospitalisation and was interviewed with an abbreviated version of the SCAN. Consensus best-estimate diagnoses were made according to DSM-IV criteria by two researchers. All participants gave written informed consent and the study was approved by local ethics committees at the participating centers.

**Kirov, G | 25055870 | UK trios | bip_uktr_eur**

The BD subjects were recruited from lithium clinics and interviewed in person by a senior psychiatrist, using the abbreviated version of the SCAN. Consensus best-estimate diagnoses were made based on the interview and hospital notes. Ethics committee approval for the study was obtained from the relevant research ethics committees and all individuals provided written informed consent for participation.

**Landén, M; Sklar, P | [ICCBD] | Sweden (ICCBD) | bip_swa2_eur**
The BD subjects were identified using the Swedish National Quality Register for Bipolar Disorders (BipoläR) and the Swedish National Patient Register (using a validated algorithm[28] requiring at least two hospitalizations with a BD diagnosis). A confirmatory telephone interview with a diagnostic review was conducted. Additional subjects were recruited from the St. Göran Bipolar Project (Affective Center at Northern Stockholm Psychiatry Clinic, Sweden), enrolling new and ongoing patients diagnosed with BD using structured clinical interviews. Diagnoses were made according to the DSM-IV criteria (BipoläR and St. Göran Bipolar Project) and ICD-10 (National Patient Register). The control subjects used were the same as for the SCZ analyses described above. All ascertainment procedures were approved by the Regional Ethical Committees in Sweden.
**Landén, M; Sklar, P | [ICCBD] | Sweden (ICCBD) | bip_swei_eur**
The cases and controls in the bip_swei_eur sample were recruited using the same ascertainment methods described for the bip_swa2_eur sample.
**Leboyer, M |[29]; [PGC1 replication] | France | bip_fran_eur**
Cases with BD1 or BD2 and control samples were recruited as part of a large study of genetics of BD in France (Paris-Creteil, Bordeaux, Nancy) with a protocol approved by relevant IRBs and with written informed consent. Cases of French descent for more than 3 generations were assessed by a trained psychiatrist or psychologist using structured interviews supplemented by medical case notes, mood scales and self-rating questionnaire assessing dimensions.
**Li, Q | 24166486; 27769005 | USA (Janssen), SAGE controls | bip_jst5_eur**
The study included unrelated patients with bipolar 1 disorder from 6 clinical trials (IDs: NCT00253162, NCT00257075, NCT00076115, NCT00299715, NCT00309699, and NCT00309686). Participant recruitment was conducted by Janssen Research & Development, LLC (formerly known as Johnson & Johnson Pharmaceutical Research & Development, LLC) to assess the efficacy and safety of risperidone. Bipolar cases were diagnosed according to DSM-IV-TR criteria. The diagnosis of bipolar disorder was confirmed by the Schedule for Affective Disorders and Schizophrenia for School-Age Children-Present and Lifetime Version (K-SADS-PL) in NCT00076115, by the SCID in NCT00257075 and NCT00253162, or by the MINI in NCT00299715 and NCT00309699, and NCT00309686, respectively. Additional detailed descriptions of these clinical trials can be found at ClinicalTrials.gov. Only patients of European ancestry with matching controls were included in the current analysis. Controls subjects were drawn from the Study of Addiction: Genetics and Environment (SAGE, dbGaP Study Accession: phs000092.v1.p1). Control subjects did not have alcohol dependence or drug dependence diagnoses; however, mood disorders were not an exclusion criterion.
**Craddock, N; Jones, I; Jones, L | [ICCBD] | Cardiff and Worcester, UK (ICCBD-BDRN) | bip_icuk_eur**
Cases were all over the age of 17 yr, living in the UK and of European descent. Cases were recruited via systematic and not systematic methods as part of the Bipolar Disorder Research Network project (www.bdrn.org), provided written informed consent and were interviewed using a semi-structured diagnostic interview, the Schedules for Clinical Assessment in Neuropsychiatry. Based on the information gathered from the interview and case notes review, best-estimate lifetime diagnosis was made according to DSM-IV. Inter-rater reliability was formally assessed using 20 randomly selected cases (mean κ Statistic = 0.85). In the current study we included cases with a lifetime diagnosis of DSM-IV bipolar disorder or schizo-affective disorder, bipolar type. The BDRN study has UK National Health Service (NHS) Research Ethics Committee approval and local Research and Development approval in all participating NHS Trusts/Health Boards.Controls were part of the Wellcome Trust Case Control Consortium common control set, which comprised healthy blood donors recruited from the UK Blood Service and samples from the 1958 British Birth Cohort. Controls were not screened for a history of mental illness. All cases and controls were recruited under protocols approved by the appropriate IRBs. All subjects gave written informed consent.

**Ophoff, RA | Not Published | Netherlands | bip_ucla_eur**
The case sample consisted of inpatients and outpatients recruited through psychiatric hospitals and institutions throughout the Netherlands. Cases with DSM-IV bipolar disorder, determined after interview with the SCID, were included in the analysis. Controls were collected in parallel at different sites in the Netherlands and were volunteers with no psychiatric history after screening with the (MINI[10]). Ethical approval was provided by UCLA and local ethics committees and all participants gave written informed consent.
**Paciga, S | [PGC1] | USA (Pfizer) | bip_pf1e_eur**
This sample comprised Caucasian individuals recruited into one of three Geodon (ziprasidone) clinical trials (NCT00141271, NCT00282464, NCT00483548). Subjects were diagnosed by a clinician with a primary diagnosis of Bipolar 1 Disorder, most recent episode depressed, with or without rapid cycling, without psychotic features, as defined in the DSM-IV-TR (296.5x) and confirmed by the MINI (version 5.0.0). Subjects also were assessed as having a HAM-D-17 total score of >20 at the screening visit. The trials were conducted in accordance with the protocols, International Conference on Harmonization of Good Clinical Practice Guidelines, and applicable local regulatory requirements and laws. Patients gave written informed consent for the collection of blood samples for DNA for use in genetic studies.
**Pato, C | [ICCBD] | Los Angeles, USA (ICCBD-GPC)| bip_usc2_eur**
Genomic Psychiatry Consortium (GPC) cases and controls were collected via the University of Southern California healthcare system, as previously described[30]. Using a combination of focused, direct interviews and data extraction from medical records, diagnoses were established using the OPCRIT and were based on DSM-IV-TR criteria. Age and gender-matched controls were ascertained from the University of Southern California health system and assessed using a validated screening instrument and medical records.

**======== PGC2 Followup Samples ========**
**Kelsoe, J | [PGC1] | USA (BiGS/TGEN1) | TGEN1_eur**
Cases and controls for this sample were ascertained using the same procedures applied for the bip_gain_eur sample described above. These samples formed a distinct PCA cluster from the samples described above and were therefore analysed separately.
**Li, Q | 24166486 | various Eastern Europe, shared T. Esku controls | JJ_EAST_eur**
The cases were drawn from the same six clinical studies described for bip_jst5_eur except that only patients of east European ancestry with matching controls were included in this cohort. Most of the Eastern European controls were from the Estonian Biobank project (EGCUT)[31] and were ancestrally matched with cases.
**Schulze, T | [ConLiGen] | Germany | BIP_KFO_eur**
The KFO sample was derived from the Clinical Research Group 241 (KFO241 consortium; www.kfo241.de) and the PsyCourse consortium (www.psycourse.de). The samples form part of a multi-site German/Austrian longitudinal study. Diagnoses were made according to DSM-IV. German Red Cross controls were collected by the Central Institute for Mental Health in Mannheim, University of Heidelberg, Germany. Volunteers who gave blood to the Red Cross were asked whether they would be willing to participate in genetic studies of psychiatric disorders. Control subjects were not selected on the basis of mental health screening.

**======== External studies ========**
**Mortensen, P; Borglum, A | Not published | [iPsych] | NA**
The iPSYCH bipolar disorder sample is a nationwide population based case-cohort sample derived from the Danish Bloodspot resource[32]. In 1981, Denmark began storing neonatal bloodspots and collected samples have been subsequently linked to the Danish Psychiatric Central Research Register (DPCRR). The

iPSYCH sample includes practically all individuals diagnosed with bipolar disorder who were born in Denmark between 1981 and 2005. Cases were diagnosed clinically by a psychiatrist at in- or out-patient psychiatric hospitals according to ICD10 as recorded in DPCRR (ICD10 codes F30-F31). Diagnoses were given in 2013 or earlier for persons not less than 10 years old. Controls were randomly selected from the same national birth cohort and not diagnosed with bipolar disorder.

DNA was prepared as described previously[33] and genotyping was done using the PsychChip array from Illumina (CA, San Diego, USA) according to the manufacturer's protocols. Genotypes were processed using the Ricopili pipeline and imputation using the 1000 genomes phase 3 reference panel. Genetic outliers were excluded based on principal component analysis. Due to the large number of study subjects in the overall iPSYCH cohort, the sample was genotyped and processed in 23 waves with each wave treated as a separate sample. Only waves with at least 100 bipolar cases were included in the analysis, and controls were down-sampled from each included wave (Ncontrols = 4 x N cases). After this processing, genotypes from 839 cases and 2938 controls were included for analysis. Due to the nature of the analyses and the overall lower number of cases we decided to relax the per wave sample size requirement for the sex-specific analysis and the analysis of chromosome X data. At least 50 female or male bipolar cases were required for a wave in order to be included in the analyses (with N controls = 4 x N cases). Please note that this still resulted in a nominal "loss of waves" that were included in the analyses when compared to the analysis of the full dataset. A total of 697 female cases and 1867 female controls as well as 111 male cases and 512 male controls were included, respectively. Processing and analysis of genotype data were performed at the secured, national high performance-computing cluster *GenomeDK* (http://genome.au.dk). The study was approved by the Danish Data Protection Agency and the Scientific Ethics Committee in Denmark.

**Stefánsson, H | [PGC1 replication] | Iceland (deCODE genetics) | deCODE**

The Icelandic sample consisted of 2,908 subjects with BD (1661 SNP typed) and 344,848 controls (141,854 SNP typed). DNA was isolated from blood samples provided by patients and controls that were recruited throughout Iceland. Approval for the study was granted by the National Bioethics Committee of Iceland and the Icelandic Data Protection Authority and informed consent was obtained for all participants providing a sample for the study. Diagnoses were assigned according to Research Diagnostic Criteria[34] through the use of the SADS-L[35] for 303 subjects. DSM-IV BD diagnoses were obtained through the use of the Composite International Diagnostic Interview (CIDI-Auto) for 82 subjects. The remaining BD subjects were diagnosed by ICD 9 or ICD 10 at Landspitali University Hospital in the years 1987-2018. Controls were recruited as a part of various genetic programs at deCODE and were not screened for psychiatric disorders. Whole genome sequencing was performed on samples from 541 BD cases and 26,014 controls. Two types of imputations were performed; into SNP-typed individuals based on long-range phasing, followed by a familial imputation step into un-typed relatives of SNP-typed individuals[36]. Cases of bipolar I disorder were defined using ICD-10 codes 31.1 and 31.2 and ICD-9 codes 296.0 and 296.2. Cases of bipolar II disorder were defined using the ICD-10 code 31.0 in the absence of ICD-10 codes F31.1 and F31.2 and ICD-9 codes 296.0 and 296.2.

**Milani L | 24518929 | Estonia (Estonian Biobank) | EstonianBiobank**

The Estonian Biobank (EstBB) is a population-based cohort of 200,000 participants with a rich variety of phenotypic and health-related information collected for each individual[31]. At recruitment, all participants signed a consent to allow follow-up linkage of their electronic health records (EHR), thereby providing a longitudinal collection of phenotypic information. Health records have been extracted from the national Health Insurance Fund Treatment Bills (from 2004), Tartu University Hospital (from 2008), and North Estonia Medical Center (from 2005). The diagnoses are coded in ICD-10 format and drug dispensing data include drug ATC codes, prescription status and purchase date (if available). For the current study, cases of bipolar disease were determined by searching the EHRs for data on F31* ICD-10 diagnosis. All remaining participants who did not have any ICD-10 F* group diagnoses were defined as controls. Cases with bipolar I disorder were those with ICD codes of F31.1 and F31.2.

**Zwart JA | Unpublished | Norway (the Nord-Trøndelag Health Study) | HUNT**

The HUNT sample consisted of 905 subjects with BD and 41,914 population controls[37]. Patients and controls were of European ancestry and were recruited from the Nord-Trøndelag County, Norway. Diagnoses were assigned according to ICD-9 or ICD-10. The controls included individuals not diagnosed with substance use disorders, schizophrenia, bipolar disorder, major depressive disorder, anxiety disorders, eating disorders, personality disorders, or ADHD in hospitals (ICD-9 or ICD-10) or general practice (ICPC2). They also were >40 years of age, had low self-reported levels of anxiety and depression (HADS-A and HADS-D $\leq$ 11), and reported no use of antidepressants, anxiolytics, or hypnotics. Approval for the study was granted by the Data Inspectorate of Norway, the Health Directorate and the Regional Committee for Medical and Health Research Ethics. Cases of bipolar I disorder were those with ICD codes of F31.1, F31.2 or F31.6 and ndividuals with an ICD-9 code of 295 or ICD-10 codes F20-F29 were excluded. Cases of bipolar II disorder were those with ICD codes of F31.8 and individuals with an ICD-9 code of 295 or ICD-10 codes F20-F29, F31.1-.2 or F31.6 were excluded.

**Breen G | 30305743 | UK (UK Biobank) | UKBiobank**

The UK Biobank is a prospective cohort study of 501,726 individuals, recruited at 23 centres across the United Kingdom[38]. Extensive phenotypic data are available for UK Biobank participants from health records and questionnaires. Participants were classified as having bipolar disorder if they had a reported clinical diagnosis of bipolar disorder (all primary and secondary ICD10 F31 code diagnoses in hospital inpatient records data; UK Biobank category 2002; http://biobank.ctsu.ox.ac.uk/showcase/label.cgi?id=2002; N = 777) or if they self-reported bipolar disorder during an interview with a nurse at baseline recruitment (UK Biobank data-field 20002; http://biobank.ctsu.ox.ac.uk/showcase/field.cgi?id=20002; N = 1,116; union N = 1,454). The selection of control participants has been described previously[39]. Control participants did not meet case criteria, did not report the use of any psychiatric medication at baseline (UK Biobank data-field 20003; http://biobank.ctsu.ox.ac.uk/showcase/field.cgi?id=20003), and did not self-report any history of mental health disorder in the online mental health questionnaire (UK Biobank category 136; http://biobank.ctsu.ox.ac.uk/showcase/label.cgi?id=136; N = 58113).

***======== PGC PsychChip Samples ========***

**Pato, C | Not published | [PGC Psychchip] | gpcw1**

The cases and controls in this study were ascertained in the same manner as those described above for bip_usc2_eur.

**Reif, A | Not published | [PGC Psychchip] | germ1**

Cases were recruited in the same manner as those described above for BOMA-Germany II | bip_bmg2_eur. Control subjects were healthy participants who were recruited from the community of the same region as cases. They were of Caucasian descent and fluent in German. Exclusion criteria were manifest or lifetime DSM-IV axis I disorder, severe medical conditions, intake of psychoactive medication as well as alcohol abuse or abuse of illicit drugs. Absence of DSM-IV axis I disorder was ascertained using the German versions of the Mini International Psychiatric Interview. IQ was above 85 as ascertained by the German version of the Culture Fair Intelligence Test 2[40]. Study protocols were reviewed and approved by the ethical committee of the Medical Faculty of the University of Würzburg. All subjects provided written informed consent.

**Serretti, A, Ribases M | Not published | [PGC Psychchip] | spsp3**

The sample includes 267 BD subjects (Spanish Wave2 Serretti PsychChip QC Summary), of which 180 Spanish and 87 Italian. Spanish sample: 180 subjects were enrolled in a naturalistic cohort study, consecutively admitted to the out-patient Bipolar Disorders Unit, Hospital Clinic, University of Barcelona. This is a systematic cross-sectional analysis deeply described in a previous paper on the same sample investigating rs10997870 SIRT1 gene variant[41]. Inclusion criteria were a diagnosis of Bipolar Disorder (type 1 or 2) according to DSM-IV TR criteria and age of 18 years or older. The study was approved by the local ethical committee and carried out in accordance with the ethical standards laid down in the Declaration of Helsinki. Signed informed consent was obtained from all participants after a detailed and extensive description of the study and patient's confidentiality was preserved. The current and lifetime diagnoses of mental disorders were formulated by independent senior psychiatrists (diagnostic concordance: Kappa=0.80) according to DSM-IV TR clinical criteria and confirmed through the semi-structured interviews for Axis I disorders according to DSM IV TR criteria (SCID I). Furthermore, all available clinical data coming from follow-up at our unit and collateral information concerning illness history were cross-referred in order to ensure accuracy and obtain complete clinical information. Specific psychopathological dimensions were assessed by means of rating scales and clinical questionnaires administered by clinicians, adequately trained to enhance inter-rater reliability. Mood episodes were defined according to DSM-IV TR criteria and their severity was measured through the administration of the 21-item Hamilton Depression Rating Scale (HDRS-21, Spanish version). The most severe depressive episode was defined on the basis of the severity at the HDRS (total score > 14) and clinical judgment. Italian sample: 87 subjects with bipolar depression were enrolled into the study when admitted at the Department of Psychiatry, University of Bologna, Italy. A description of the subjects has been previously reported when analyzing clinical features[42]. Inclusion criteria were: a diagnosis of bipolar disorder, most recent episode depressive as assessed by DSM-IV-TR criteria; Young Mania Rating Scale (YMRS) score <12; Hamilton Depression Rating Scale (HAM-D) <12. Exclusion criteria were: presence of a bipolar disorder, most recent episode manic or hypomanic; presence of severe medical conditions; presence of moderate to severe dementia (Mini Mental State Examination score <20). The following scales were administered biweekly during the hospitalization: HAM-D, Hamilton Anxiety Rating Scale (HAM-A), YMRS and Dosage Record and Treatment Emergent Symptom Scale (DOTES). Written informed consent was obtained for each patient recruited. The study protocol was approved by the local Ethical Committee and it has been performed in accordance with the ethical standards laid down in the 1975 Declaration of Helsinki.

The Spanish controls were part of the Mental-Cat clinical sample or the INSchool population-based cohort. A total of 1,774 controls from the Mental-Cat cohort (60.5% males) were evaluated and recruited prospectively from a restricted geographic area at the Hospital Universitari Vall d'Hebron of Barcelona (Spain) and consisted of unrelated healthy blood donors. The INSchool sample consisting of 771 children (76.2% males) from schools in Catalonia were involved for screening using the Achenbach System of Empirically Based Assessment (ASEBA) with the Child Behavior Checklist CBCL/4-18 (completed by parents or surrogates), the Teacher Report Form TRF/5-18 (completed by teachers and other school staff) and the Youth Self-Report YSR/11-18 (completed by youths); the Strengths and Difficulties Questionnaire (SDQ) and the Conner's ADHD Rating Scales (Parents and Teachers). Genomic DNA samples were obtained either from peripheral blood lymphocytes by the salting out procedure or from saliva using the Oragene DNA Self-Collection Kit (DNA Genotek, Kanata, Ontario Canada). DNA concentrations were determined using the Pico- Green dsDNA Quantitation Kit (Molecular Probes, Eugene, OR) and genotyped with the Illumina Infinium PsychArray-24 v1.1 at the Genomics Platform of the Broad Institute. The study was approved by the Clinical Research Ethics Committee (CREC) of Hospital Universitari Vall d'Hebron, all methods were performed in accordance with the relevant guidelines and regulations and written informed consent was obtained from participant parents before inclusion into the study. Detailed information has been published previously[43].

**Perlis, R; Sklar, P; Smoller, J, Goes F, Mathews CA, Waldman I | Not published | [PGC Psychchip] | usaw4**

Perlis, R; Sklar, P; Smoller, J: EHR data were obtained from a health care system of more than 4.6 million patients[44] spanning more than 20 years. Experienced clinicians reviewed charts to identify text features and coded data consistent or inconsistent with a diagnosis of bipolar disorder. Natural language processing was used to train a diagnostic algorithm with 95% specificity for classifying bipolar disorder. Filtered coded data were used to derive three additional classification rules for case subjects and one for control subjects. The positive predictive value (PPV) of EHR-based bipolar disorder and subphenotype diagnoses was calculated against diagnoses from direct semistructured interviews of 190 patients by trained clinicians blind to EHR diagnosis. The PPV of bipolar disorder defined by natural language processing was 0.86. Coded classification based on strict filtering achieved a value of 0.84, but classifications based on less stringent criteria performed less well. No EHR-classified control subject received a diagnosis of bipolar disorder on the basis of direct interview (PPV=1.0). For most subphenotypes, PPV exceeded 0.80. The EHR-based classifications were used to accrue bipolar disorder cases and controls for genetic analyses. Samples were genotyped on the Psychchip array.

Goes, FS: Cases represented independent probands from a European-American family sample that was collected at Johns Hopkins University from 1988-2010. Families had at least 2 additional relatives with a major mood disorder (defined as bipolar disorder type 1, bipolar type 2 or recurrent major depressive disorder). Diagnostic interviews were performed using the Schedule for Affective Disorders and Schizophrenia-Lifetime Version (N=81) and the Diagnostic Instrument for Genetics Studies (N=161). All cases underwent best-estimate diagnostic procedures. After genotyping quality control there were 242 cases, of which 240 were diagnosed as Bipolar Disorder type 1 and 2 as Schizoaffective Disorder, bipolar type. Diagnoses were based on DSM-III and DSM-IV criteria. Probands from this sample have been previously studied in family based linkage and exome studies.[45–47]

Mathews CA: Control samples were ascertained as part of ongoing genetic and neurophysiological studies of hoarding, obsessive compulsive and tic disorders. Controls reported no current or lifetime history of mania or hypomania at the time of ascertainment. Sixty-two of the 104 controls were screened for psychiatric illness using the Structured Clinical Interview for DSM-IV TR diagnoses and diagnoses of bipolar disorder, lifetime or current, were ruled out through a best estimate consensus diagnosis. Other psychiatric diagnoses were not excluded. The remaining 42 participants were not formally screened, but reported no lifetime or current history of bipolar disorder, obsessive compulsive, hoarding, or tic disorders. Samples were genotyped on the Psychchip array. Ethical approvals were obtained from the University of Florida Human Subjects Review Board.

Waldman I: Control samples were ascertained as part of an ongoing genetic study of ADHD and other Externalizing disorders (I.e., Oppositional Defiant Disorder and Conduct Disorder). Controls reported no current diagnoses of Externalizing or Internalizing disorders at the time of ascertainment. Controls were assessed for psychiatric conditions using the Emory Diagnostic Rating Scale (EDRS)[48], a questionnaire that assessed parent ratings of symptoms of common DSM-IV Externalizing and Internalizing disorders (e.g., Major Depressive Disorder and various anxiety disorders). Samples were genotyped on the Psychchip array. Ethical approvals were obtained from the Emory University and University of Arizona Human Subjects Review Boards.

**Baune, BT; Dannlowski, U | Not published | [PGC Psychchip] | bdtrs**

The Bipolar Disorder treatment response Study (BP-TRS) comprises BD inpatient cases and screened controls of Caucasian background. Psychiatric diagnosis of Bipolar Disorders was ascertained using SCID or MINI 6.0 using DSM-IV criteria in a face-to-face interview by a trained psychologist / psychiatrist for both cases and controls. Healthy controls were included if no current or lifetime psychiatric diagnosis was identified. Cases were included if current or lifetime diagnosis of bipolar disorder was ascertained by structured diagnostic interview. Cases and controls are of similar age range (>=18 yrs of age) and were collected from the same geographical areas. Other assessments including symptom ratings, psychiatric history, treatment history, treatment response were based on interview, and carried out by trained psychologists/psychiatrists. Samples were genotyped on the Psychchip array. Ethical approval was obtained from the University of Münster Human Ethics Committee, Münster, Germany.

**Ophoff R, Posthuma D, Lochner C, Franke B | Not published | [PGC Psychchip] | dutch**

Ophoff R: Cases and controls were collected using the same protocol as described above for the "ucla" sample.

Lochner C: Controls include population based-controls ascertained from blood banks and controls recruited through university campuses and newspaper advertisements, who underwent a psychiatric interview and had no current or lifetime psychiatric disorder [49,50].

Franke B: The controls included are healthy individuals from the Dutch part of the International Multicenter ADHD Genetics (IMAGE) project[51,52].

Posthuma D: Data were provided for 960 unscreened Dutch population controls from the Netherlands Study of Cognition, Environment and Genes (NESCOG)[53]. The study was approved by the institutional review board of Vrije Universiteit Amsterdam and participants provided informed consent.

**Gawlik M | Not published | [PGC Psychchip] | gawli**

Patients were recruited at the Department of Psychiatry, Psychosomatics and Psychotherapy, University of Würzburg, Germany. Diagnosis according to DSM-IV (Diagnostic and Statistical Manual of Mental Disorders-fourth edition) was made by the best estimate lifetime diagnosis method, based on all available information, including medical records, and the family history method.

**Fullerton J, Mitchell PB, Schofield PR, Green MJ, Weickert CS, Weickert TW, The Australian Schizophrenia Research Bank | Not published | [PGC Psychchip] | neuc1**

The NeuRA collection comprised BD cases from three cohorts ascertained in Australia: the bipolar high risk study[54] (n=97), the Imaging Genetics in Psychosis Study (IGP; n=47)[55] and a clinic sample (n=109) recruited via the Sydney Bipolar Disorders Clinic[56]. The clinic sample used the same ascertainment procedures as described for the bip_bmau_eur sample. The bipolar high risk study is a collaborative study with 4 US and one Australian groups, with young participants aged 12-30. The IGP sample was recruited from outpatient services of the South Eastern Sydney-Illawarra Area Health Service (SESIAHS), the Sydney Bipolar Disorders Clinic and the Australian Schizophrenia Research Bank. Healthy controls were sourced from the high risk, IGP and the Cognitive and Affective Symptoms of Schizophrenia Intervention (CASSI) trial[57] studies, and were recruited from the community, had no personal lifetime history of a DSM-IV Axis-I diagnosis as determined by psychiatric interview, and no history of psychotic disorders among first-degree biological relatives. Additional controls were recruited as part of the strategy to develop an Australian Schizophrenia Research Biobank for studies into the genetics of this disease. The ascertainment of these controls has been previously described[58].

**Landen M, Hillert J, Alfredsson L | Not published | [PGC Psychchip] | swed1**

The cases in the swed1 sample were recruited using the same ascertainment methods described for the bip_swa2_eur sample. Population-based healthy controls, randomly selected from the Swedish national population register, were collected as part of two case-control studies of multiple sclerosis: GEMS (Genes and Environment in Multiple Sclerosis) and EIMS (Epidemiological Investigation of Multiple Sclerosis)[59].

**Di Florio A, McQuillin A, McIntosh A, Breen G | Not published | [PGC Psychchip] | ukwa1**

McQuillin A: BD cases were recruited using the same protocol as the bip_uclo_eur described above. A subset (n=448) of the control subjects were random UK blood donors obtained from the ECACC DNA Panels (https://www.phe-culturecollections.org.uk/products/dna/hrcdna/hrcdna.jsp). The remaining control subjects (n=814) had been screened for an absence of mental illness in using the same protocol as the bip_uclo_eur described above.

Di Florio A: Cases were recruited across the United Kingdom in the same manner as described for the bip_wtcc_eur and bip_icuk_eur samples.

McIntosh AM: BD cases were recruited from the clinical case loads of treating psychiatrists from Edinburgh and across the central belt of Scotland. Controls were identified from non-genetic family members and from the extended networks of the participants themselves. All participants were of

European ancestry and diagnosis was confirmed using an established battery developed for ICCCBD. Breen G: Controls were drawn from blood donors to the UK Motor Neuron Disease Association DNA Biobank[60]

**Perlis, R; Sklar, P; Smoller, J, Nievergelt C, Kelsoe J | Not published | [PGC Psychchip] | usaw5**

Kelsoe, J: The Pharmacogenomics of Bipolar Disorder (PGBD) study was a prospective assessment of lithium response in BDI patients. The goal was to identify genes for lithium response. Subjects were recruited from clinics at 11 international sites and followed for up to 2.5 years. Diagnosis was obtained by DIGS interview and medical records reviewed by blind experienced clinicians. As the comparison was between lithium responders and non-responders, no controls were collected. All subjects provided written informed consent.

Perlis R: Cases of bipolar disorder were Individuals treated with lithium drawn from the Partners Healthcare electronic health record (EHR) database, which spans two large academic medical centers, Massachusetts General Hospital and Brigham and Women's Hospital in addition to community and specialty outpatient clinics[61]. Any patients aged 18 years or older with at least one lithium prescription between 2006 and 2013 based on e-prescribing data were included. The Partners Institutional Review Board approved all aspects of this study. Individuals with a diagnosis of schizophrenia based on ICD9 codes were excluded.

Smoller J: Cases and controls were recruited in the same manner as described above for "usaw4".

**======== PGC3 Samples ========**

**Rietschel M, Nöthen MM, Forstner AJ, Streit F, Babadjanova G |24618891| Russia (BOMA-Russia) | bmrus**

Patients were recruited from consecutive admissions to the psychiatric inpatient units of the Russian State Medical University, Moscow. Unrelated controls were recruited from the general population. All protocols and procedures were approved by the respective local Ethics Committees. Written informed consent was obtained from all study participants before the study participation. All patients were assigned a lifetime diagnosis of BPAD type I or type II. This was based on Diagnostic and Statistical Manual of Mental Disorders-IV criteria and a consensus best-estimate procedure, including a structured interview-I, review of medical records, the family history method and the Operational Criteria Checklist for Psychotic Illness OPCRIT system.

**Ferentinos P, Dikeos D, Patrinos G | Not published | Greece (Attikon General Hospital) | greek**

All adult patients with a DSM-IV-TR/DSM-5 diagnosis of Bipolar Disorder hospitalized at the inpatient unit or followed-up at the specialized 'Affective disorders and Suicide' outpatient clinic of the 2nd Department of Psychiatry, National and Kapodistrian University of Athens, Attikon General Hospital, Athens, Greece from 2012 to 2017 were recruited for the current study. Patients were referred to the specialized 'Affective disorders and Suicide' outpatient clinic either from the inpatient unit after hospitalization or from the community. Diagnosis was established and demographic (age, gender, family status, profession, employment status, education) and relevant clinical features (e.g. age at onset, polarity of first and most recent episode, number of lifetime depressive and manic/hypomanic episodes, number of hospitalizations, lifetime suicidality, lifetime psychosis) were extracted through a M.I.N.I.-5.0.0-based semi-structured diagnostic interview, which was administered during patients' initial clinical assessment and regularly updated ever since, interviews of primary caregivers and inspection of medical records. Lifetime presence of any DSM-IV-TR axis I psychiatric comorbidities (dysthymia, panic disorder, agoraphobia, social phobia, generalized anxiety disorder, obsessive-compulsive disorder, post-traumatic stress disorder, alcohol and substance abuse and dependence, anorexia nervosa, bulimia nervosa) was similarly extracted. Family history of major psychiatric disorders and suicidality in first and second degree relatives was recorded with a specific questionnaire based on the Family Interview for Genetic Studies. Medical comorbidities were recorded with the Cumulative Illness Rating Scale, completed on the basis of interview with patient and primary caregivers, inspection of patient's medical records and laboratory exams (basic or specific, if considered necessary). Presence of selected medical diseases was specifically recorded.

Control (unaffected) participants were a convenient sample drawn from the same geographic area as case participants, either within health care facilities or as community volunteers. All of them went through a brief clinical interview including items on psychiatric and medical history, psychiatric family history, past and current medical or psychiatric therapies, and a brief mental state examination. Only participants found to be free of lifetime major mental disorders (MDD, BD, schizophrenia, or other psychotic disorders) and with no family history of major mental disorder in their first-degree relatives were recruited as controls.

All cases and controls were native Greek speakers. All participants provided written informed consent before being included in the study and the study protocol was approved by the Research Ethics Committee of Attikon General Hospital.

**Andreassen, OA | Not published | Norway (TOP) | norgs**

The NORGS bipolar disorder cases and controls were ascertained in the same way as the bip_top7_eur (TOP7) samples described above, and recruited from hospitals across Norway.

**Andreassen, OA | Not published | Norway (TOP) | noroe**

The NOROE bipolar disorder cases and controls were ascertained in the same way as the bip_top7_eur (TOP7) samples described above, and recruited from hospitals across Norway.

**Reininghaus E | Not published| Austria (Medical University of Graz) | graza**

Assoz.Prof. DDr. Eva Reininghaus, Priv.Doz. DDr. Susanne Bengesser, Priv.Doz. Dr. Nina Dalkner, Priv.Doz. Armin Birner and further team members of the special outpatients department for bipolar affective disorders at the Department of Psychiatry and Psychotherapeutic Medicine, Medical University of Graz, Austria: Cases with bipolar affective disorder (type I and II) and healthy controls were recruited at the Department of Psychiatry and Psychotherapeutic Medicine at the Medical University of Graz (MUG), Austria. Study protocols were approved by the ethics committee of the Medical University of Graz. Patients and healthy controls gave written informed consent and the study was conducted according to the declaration of Helsinki. All patients received a clinical interview by a psychiatrist or psychologist and a diagnosis according to DSM-IV with the SCID-I (Structured clinical interview). Healthy controls did not have a history of a psychiatric disorder. Furthermore, healthy controls did not have any first or second degree relatives with a psychiatric disorder. The PGC-Graz sample (n= 244; 114 males, 130 females) includes 167 cases with bipolar disorder and 77 healthy controls genotyped with Omniexpress 1.2 by Illumina.

**Grigoroiu-Serbanescu M | 31791676; 26806518| | Romania (BOMA-Romania) | bmrom**

This sample includes the BOMA-Romania sample and additional cases from the ConLiGen-Romania sample. For the BOMA-Romania sample, unrelated BP-I patients were recruited from consecutive admissions in the Obregia Psychiatric Hospital of Bucharest, Romania. All participants provided written informed consent following a detailed explanation of the study aims and procedures. The study was performed in accordance with the Code of Ethics of the World Medical Association (Declaration of Helsinki). All participants were of Romanian descent according to self-reported ancestry. Genealogical information about parents and all four grandparents was obtained through direct interview of the subjects.

The patients were investigated with the Diagnostic Interview for Genetic Studies (DIGS)[27] and the Family Interview for Genetic Studies (FIGS)[7] The diagnosis of BP-I was assigned according to DSM-IV criteria on the basis of both the DIGS and medical records. Patients were included in the sample if they had at least two documented hospitalized illness episodes (one manic/mixed and one depressive or two manic episodes) and no residual mood incongruent psychotic symptoms during remissions. This information was also confirmed by first degree relatives for 64% of the cases. The illness age-of-onset was defined as the age at which the proband first met DSM-IV criteria for a manic, mixed, or major depressive episode. Family history of psychiatric illness was obtained with FIGS administered both to the patients and to all available relatives.

Cases in the ConLiGen-Romania study were ascertained in the same manner as for BOMA-Romania. Cases were required to have taken lithium for at least two years and lithium treatment response was evaluated with the Alda scale[62].

Population-based controls were evaluated using the DIGS and FIGS to screen for a lifetime history of major affective disorders, schizoaffective disorders, SCZ and other psychoses, obsessive-compulsive disorder, eating disorders, and alcohol or drug addiction. Unaffected individuals were included as controls in the present study.

**Quality control, imputation and analysis of cohorts external to the PGC**

For external cohorts, quality control (QC), imputation and GWAS were conducted by the collaborating research teams using comparable procedures as used for the PGC cohorts. These are outlined below. SNPs were retained in the GWAS summary statistics using the filtering method described for the PGC cohorts.

**iPSYCH**

For the iPSYCH cohort, QC, imputation and GWAS was performed using RICOPILI, as described for the PGC cohorts[63].

**deCODE genetics**

QC, imputation and association analyses were performed in the deCODE sample as previously described [36,64].

**Estonian Biobank**

A more detailed description of the genotyping, quality control and imputation procedures for the Estonian Biobank (EstBB) is reported elsewhere[65,66]. As a short description, of all the studied EstBB participants at the time of this study, 33,277 have been genotyped using the Global Screening Array v1, 8137 on the HumanOmniExpress beadchip, 2641 on the HumanCNV370-Duo BeadChips and 7,832 on the Infinium CoreExome-24 BeadChips from Illumina. Furthermore, 2,056 individuals' whole genomes have been sequenced at the Genomics Platform of the Broad Institute. Sequenced reads were aligned against the GRCh37/hg19 version of the human genome reference using BWA-MEM1 v0.7.7. The genotype data was phased using Eagle2 (v. 2.3) and imputed using BEAGLE (v. 4.1) software, implementing a joint Estonian and Finnish reference panel (described in [65]).

The GWAS was performed among 17,616 unrelated individuals (PiHat < 0.2) of whom 408 were cases of bipolar disorder and 17,209 were controls. The GWAS was run with the EPACTS software on variants with an allele frequency of at least 0.01% using an additive genetic logistic model (b.wald). To minimize the effects of population admixture and stratification, the analyses only included samples with European ancestry based on principal component analysis (PCA) and were adjusted for the first ten principal components (PCs) of the genotype matrix, as well as for birth year, birth year squared, gender and genotyping array. By the time of the analysis of the BDI phenotype, all 200,000 EstBB participants had been genotyped with the Global Screening Array and imputed using the Estonian reference panel. The BDI GWAS with 147 cases and 65,952 controls was performed using SAIGE, including related individuals and adjusting for the first ten PCs, as well as for birth year, birth year squared and sex.

**HUNT**

Participants were genotyped with Illumina HumanCoreExome arrays (HumanCoreExome12 v1.0, HumanCoreExome12 v1.1, or UM HUNT Biobank v1.0). In quality control, genotypes with call rates <99%, contamination >2.5%, large CNVs, lower call rate of technical duplicate pair or twins, uncommon sex chromosome constellations, and discrepancies with reported sex were removed. Variants with call rates <99%, higher call rates genotyped in another assay, probe sequences not mapping to the reference genome, cluster separation <0.3, gentrain score < 0.15, or HWE deviation from unrelated samples of European ancestry (p<0.0001) were also removed. Imputation was performed against a customized merged reference panel of 2,201 low-coverage whole-genome sequenced samples from the HUNT study and the Haplotype Reference consortium release 1.1 (excluding 1,023 samples from the HUNT study). The Scalable and Accurate Implementation of GEneralized mixed model (SAIGE) was used for association testing to account for case-control imbalance and relatedness[67].

**UK Biobank**

Genotypic data were available for 488,380 individuals and were imputed to the HRC, UK10K and 1,000 Genomes Phase 3 reference panels using IMPUTE4 to identify ≈ 93M variants for 487,409 individuals[68]. Variants for analysis were limited to those with minor allele frequency >= 0.01, imputation INFO-score >= 0.4, and which were either genotyped or imputed to the HRC reference panel, leaving a total of 7794483 SNPs for analysis. Using the genotyped SNPs, individuals were removed if: recommended by the UK Biobank core analysis team for unusual levels of missingness or heterozygosity; SNP genotype call rate < 98%; related to another individual in the dataset (KING r < 0.044, equivalent to removing up to third-degree relatives inclusive); phenotypic and genotypic gender information was discordant (X-chromosome homozygosity (FX) < 0.9 for phenotypic males, FX > 0.5 for phenotypic females). Removal of relatives was performed using a greedy algorithm, which minimises exclusions (for example, by excluding the child in a mother-father-child trio). All analyses were limited to individuals of White Western European ancestry, as defined by 4-means clustering on the first two genetic principal components provided by the UK Biobank[68]. Principal component analysis was also performed on the European-only subset of the data using the software flashpca2[69]. A genome-wide association study was performed using BGenie v.1.2[68], covarying for 6 PCs, and factors capturing site of recruitment and genotyping batch.

**Sample descriptions and polygenic risk scoring in non-European cohorts**

Polygenic risk scores (PRS) generated from the GWAS meta-analysis were tested for association with BD in four non-European cohorts, to investigate the cross-ancestry utility of PRS. The BD PRS were computed using summary statistics from the PGC1[1], PGC2[2], and PGC3 GWAS of BD to assess prediction performance in diverse ancestry samples, as the size of the European ancestry training sample increased. Analyses were conducted using PRSice-2[70], with P value informed clumping based on the LD structure of the target dataset. Following the PRS strategy of Bigdeli *et al*[71], subsets of SNPs were selected from the results at nine increasingly liberal P value thresholds ($P_T$) ($P_T$< 5E-08, $P_T$< 1E-04, $P_T$< 1E-03, $P_T$< 0.01, $P_T$< 0.05, $P_T$< 0.1, $P_T$< 0.2, $P_T$< 0.5, $P_T$< 1) as well as eight different LD-clumping $r^2$ parameters (clump-r2 = 0.1, 0.2, 0.3, ... , 0.8). The phenotypic variance explained by the PRS ($R^2$) was calculated on the liability scale using a BD population prevalence of both 1% and 2%. Each of the non-European samples are described below.

**Japan (advanced COSMO and Biobank Japan) | PMID: 28115744**

A detailed description of the sample information, genotyping, quality control and imputation procedures is reported elsewhere[72]. In brief, 2,964 BD and 61,887 comparison subjects from the Japanese population were included in this dataset (genotyped by Illumina OmniExpressExome v1.0 or v1.2 BeadChips). After the imputation and stringent QC, a total of 6,195,093 imputed SNPs were analysed for the association analysis. The diagnosis for each case subject followed the DSM-IV-TR criteria for BD and schizoaffective disorder and was reached by the consensus of at least two experienced psychiatrists, based on unstructured interviews with the subject and their family, as well as a review of the subject's medical records. For the comparison subjects, we used GWAS data for subjects in the BioBank Japan project collected as case subjects for non-psychiatric disorders. These subjects were not psychiatrically evaluated.

**Korea**

We genotyped 807 patients with bipolar disorder, 726 patients with schizophrenia and 497 healthy control subjects using the Affymetrix Axiom® Korea Biobank Array 1.0 (K-CHIP). K-CHIP was designed by the Center for Genome Science at the Korea National Institute of Health, including 833K SNPs. A more detailed description of the genotyping procedure is reported elsewhere[73]. We performed sample-level and variant-level QC of genotype data. We excluded variants with missing rate > 1%, Hardy-Weinberg equilibrium $P < 10^{-6}$, or minor allele frequency < 1%, and samples with missing rate > 5%, relatedness among the sample, mismatch between self-reported and inferred sex, or deviated heterozygosity rate. We confirmed homogeneity of the samples based on visual inspection of principal component analysis plots. Genotype imputation was conducted using the Haplotype Reference Consortium (HRC) reference panel. After the imputation and additional post-QC ($R^2 > 0.8$ and minor allele frequency > 1%), a total of 770 bipolar cases and 497 controls and 5,483,856 variants were analysed for polygenic risk score. All the patients met the DSM-IV-TR diagnostic criteria for bipolar I disorder and bipolar II disorder. For clinical diagnosis, a structured interview using the Korean version of the Diagnostic Interview for Genetic Studies (DIGS) or the Structured Clinical Interview for DSM-IV (SCID) was performed. The control group consisted of volunteers from the community who were free of any history of clinically significant psychiatric symptoms. Detailed assessment processes are described elsewhere[74].

**GAIN (admixed African American) (USA)**

Genetic Association Information Network (GAIN)/ The Bipolar Genome Study (BiGS) Data from the existing National Institutes of Health Genetic Association Information Network (GAIN) study of bipolar disorder was obtained through dbGap: phs000017.v3.p1. The GAIN study was multi-site and informed consent and institutional review board approval were obtained and details are described above for the GAIN-European data. Bipolar I diagnosis was confirmed with the structured Diagnostic Interview for Genetic Studies (DIGS) for the assessment of major mood and psychotic disorders and their spectrum conditions. The admixed African American (AA) bipolar patient data used in this study were from unrelated individuals in multiplex families and assessed with DIGS version 4. The genotyping has been described previously[75]. Briefly, genotyping of AA samples (347 BD cases; 669 controls) was carried out separately from European American (EA) samples, using the Affymetrix Genome-Wide Human SNP Array 6.0. Further quality controls were carried out to remove samples with low call rate (below 98.5% for EA and 97.8% for AA), excessively high or low heterozygosity (between 0.344 and 0.363 for EA and between 0.29 and 0.324 for AA), or incompatibility between reported gender and genetically determined gender. Samples were also checked for unexpected familial relationships using pairwise IBD estimation in PLINK. The total number of SNPs passing all initial QC tests was 845,814 for AA. Genotype imputation was conducted using the Consortium on Asthma among African ancestry Populations in the Americas (CAAPA) reference panel. After the imputation and additional post-QC (dosage $R^2 > 0.7$), a total of 347 bipolar cases and 669 controls and 10,762,719 variants were analysed for polygenic risk score.

**Genomic Psychiatry Cohort (GPC) (admixed African American) (USA)**

Details of ascertainment and diagnosis, genotyping and quality control have been described in detail previously[71]. Briefly, cases were ascertained using the Diagnostic Interview for Psychosis and Affective Disorders (DI-PAD), a semi-structured clinical interview administered by mental health professionals, which was developed specifically for the GPC study. Individuals reporting no lifetime symptoms indicative of psychosis or mania and who have no first-degree relatives with these symptoms are included as control participants. Genotyping of the AA-GPC was performed in 7 'batches' using Illumina Infinium arrays (Omni2.5, Multi-Ethnic Global Array, and Global Screening Array). Typed variants were aligned to the human reference genome (GRCh37), and within each genotyping batch, variants with missingness greater than 2% or Hardy-Weinberg Equilibrium $P$-value$<10^{-6}$ were excluded; all scripts for pre-processing GWAS array data are downloadable from https://github.com/freeseek/gwaspipeline.

Computational phasing and statistical genotype imputation were performed for each genotyping batch using Eagle (v2.3.5)[76] and Minimac3 (v2.0.1)[77], respectively, with default parameters and using publicly available reference haplotypes from the 1000 Genomes Project (1KGP) Phase 3[78]. Principal components analysis (PCA) was performed with GCTA (v1.2.4)[79], using a genome-wide genetic relatedness matrix (GRM) estimated for the full GPC dataset and reference samples from the 1KGP Phase 3 data based on 34,918 genotyped SNPs. For each individual, we estimated genome-wide average proportions of African (AFR), European (EUR), Admixed American (AMR), East Asian (EAS), and South Asian (SAS) ancestry from global ancestry PCs using a simple linear mixed model. PRS for BD were tested in two groups of cases and controls: 1766 cases and 2535 controls with >=25% African ancestry and 1636 cases and 2357 controls with >= 50% African ancestry. Associations between polygenic scores and case-control status were evaluated by logistic regression, with the first six global ancestry PCs and a batch indicator included as covariates.

**Selection of traits for Mendelian randomization**

BD has been linked to a range of other psychiatric, cognitive and behavioral phenotypes by clinical and epidemiological studies. On the basis of such studies, we selected 17 traits of interest for investigation of their genetic and potential causal relationships with BD. Traits were selected by a team of clinicians and biostatisticians, considering key clinical questions and the availability of GWAS summary statistics for the traits. Below we list the traits initially selected and the rationale for their inclusion. Only traits with at least 10 genome-wide significant loci were sufficiently powered to be investigated using Mendelian randomization, resulting in 10 traits tested (Supplementary Table 18).

**Sleep traits**

Reduced sleep duration is a diagnostic criterion for mania[102] and has been implicated both as a prodromal symptom[103] and trigger of illness episodes[104]. Hypersomnia and insomnia are commonly reported during major depressive episodes in bipolar disorder[105–107] and have been identified as residual symptoms associated with impairment[107–109]. Near-24-hour (circadian) oscillations are found in almost every human physiological process, including sleep-wake cycles[110]. Robust evidence associates bipolar disorder with a delayed sleep phase (i.e. evening chronotype)[111–113]. Interventions targeting sleep are common for the treatment of bipolar disorder, ranging from the use of sedating medication for the treatment of acute mania[114,115] to circadian manipulation[114,116] to prevent recurrences and improve outcomes. Until recently, there was no clear evidence supporting a genetic relationship between sleep and bipolar disorder. However, a recent study on over 20,000 participants found that the polygenic association between sleep and bipolar disorder differs across sleep traits and bipolar subtypes[117], with sleep duration associated to bipolar I disorder and insomnia to bipolar II disorder, but not vice versa. The study also did not find any evidence to support a causal relationship between sleep and bipolar phenotypes.

**Alcohol and substance use and misuse**

Of all psychotic and affective disorders, bipolar disorder has been reported to be the most strongly linked with alcohol or drug abuse. Compared to other primary psychiatric diagnoses, mania and hypomania may have one of the highest associations with alcohol use disorders, with a pooled lifetime prevalence around 35%[118]. Even when criteria for alcohol use disorders are not met, increased levels of alcohol use in bipolar disorder are associated with a less favourable illness course[119]. Lifetime co-occurrence rates for other substances are also high in bipolar disorder, with mean rates of 20% for cannabis use and 17% of any drug use disorders, according to a meta-analysis of clinical studies[120]. Conversely, individuals with substance use disorders have higher rates of bipolar disorder compared to non-users, with significant pooled odds ratios for both lifetime (OR 4.68,95% CI 3.39–6.47) and 12 months drug use disorders (OR 6.49, 95% CI 4.30–9.80), according to a meta-analysis of national surveys of general populations)[121]. In our research, we particularly focussed on cigarette smoking, as a recent Mendelian randomization study has suggested a causal link between smoking behaviors and bipolar disorder[122].

**Educational attainment and measures of intelligence**

The link between bipolar disorders and measures of intelligence or educational attainment is controversial. Evidence from a longitudinal whole population cohort study suggested that the association between educational attainment and risk of subsequent bipolar disorder follows a non-linear distribution: individuals with excellent school performance had the highest increased risk of later bipolar disorder compared with those with average grades (hazard ratio HR = 3.79, 95% CI 2.11–6.82). Yet, at the other end of the distribution, individuals with the poorest grades had also an increased risk of bipolar disorder (HR = 1.86, 95% CI 1.06–3.28), but the risk for them was not as high as for excellent students[123].

The association of bipolar disorder with educational attainment may differ from that with measures of intelligence. A Dutch study corroborated this hypothesis by finding associations in opposite directions for educational attainment (positive association with bipolar disorder) and measures of intelligence quotient (negative association with bipolar disorder)[124]. Moreover, it found that the association with educational attainment was specific for bipolar disorder and did not extend to schizophrenia.

Molecular genetic studies have also found an association between bipolar disorder and educational attainment ($r_{:}$0.25; $r_{LD}$:0.28)[125], but results for intelligence are equivocal[126–128]. Evidence from a population-based longitudinal study has suggested that the polygenic burden for bipolar disorder manifests as impaired cognitive performance in 8-year-old children from the general population[129]. The association, however, seemed to be driven by genetic variants shared with schizophrenia.

Such distinction between bipolar disorder and schizophrenia has also been supported by conditional false discovery rate genome-wide analyses[128]. Here, the majority of bipolar disorder risk alleles were associated with better cognitive performance, while the association was in the opposite direction (i.e. impaired cognitive performance) for schizophrenia. Among BD risk alleles identified at a lower significance threshold there was a balanced mix of bipolar disorder risk alleles associated with better or poorer cognitive performance[128]. This is in line with the non-significant genetic correlation between BD and intelligence[2], and the non-linear association between risk of bipolar disorder and school performance[123].

**Mood instability**

Mood instability does not have a shared, agreed definition[130,131]. Although it is present in many psychiatric phenotypes, the association with bipolar disorder is particularly striking. Chronic mood instability is present between illness episodes, with longitudinal studies suggesting that it is actually more common than discrete episodes[132–135]. Mood instability in bipolar disorder is of clinical relevance as it is associated with poor prognosis[132–134,136–139]. Although the mechanisms linking mood instability and bipolar disorder are not clear, a neurocomputational model has suggested that mood bias observed in bipolar disorder affects the striatal response to rewards, increasing reward prediction errors, and, in turn, causing expectations and mood to oscillate[140]. A recent UK biobank genome wide association study of mood instability has, however, found only a weak genetic association between bipolar disorder and mood instability ($r_g$=0.09; s.e.=0.037)[141]. Authors have suggested that the "mood instability" construct elicited in the general population by the question "Does your mood often go up and down?" is different from that experienced in the context of bipolar disorder, supporting the importance of phenotype definitions and the heterogeneity of mood instability.

**Brain volumes**

Neuroimaging research in bipolar disorder has been hindered by the lack of statistical power of small studies. The ENIGMA Bipolar Disorder Working Group[142] has overcome the problem by integrating data from 28 international cohorts in the largest brain magnetic resonance imaging study of bipolar disorder to date. They compared cortical grey matter thickness and surface in 1837 BD individuals and 2582 controls and found significant associations between reduced cortical surface area and history of psychosis (but not mood state at the time of scanning) and between cortical thickness and duration of illness. They also found an age-by-diagnosis interaction and an association with medication use.

**Physical activity**

Guidelines suggest physical activity for patients with bipolar disorder, especially for those taking antipsychotics and long-term medication[143]. In a systematic review of 15,587 patients with bipolar disorder, the prevalence of sedentary lifestyle varied from 40% to 64.9%[144]. Despite the high burden, the review concluded that the evidence was “insufficient to establish a cause-effect relationship between mood and physical exercise”. A recent 2 sample Mendelian randomization study on 5 SNPs associated with overall physical activity was also inconclusive[145]. One of the methods employed, however, suggested a protective causal association from overall physical activity to BD (OR, 0.491; 95% CI: 0.314–0.767; p=0.002).

**Childhood-onset psychiatric disorders**

Youths with BD have higher rates of attention deficit/hyperactivity disorder (ADHD)[146] and childhood ADHD has been found to prospectively predict later BD[147]. It has been reported that a clinically significant proportion of youth with bipolar I disorder also suffer from comorbid autism spectrum disorder[148]

**Full acknowledgements**

We thank the participants who donated their time, life experiences and DNA to this research, and the clinical and scientific teams that worked with them. We are deeply indebted to the investigators who comprise the PGC. The PGC has received major funding from the US National Institute of Mental Health (PGC3: U01 MH109528, ; PGC2: U01 MH094421; PGC1: U01 MH085520). Statistical analyses were carried out on the NL Genetic Cluster Computer (http://www.geneticcluster.org ) hosted by SURFsara and the Mount Sinai high performance computing cluster (http://hpc.mssm.edu), which is supported by the Office of Research Infrastructure of the National Institutes of Health under award numbers S10OD018522 and S10OD026880. The content is solely the responsibility of the authors and does not necessarily represent the official views of the National Institutes of Health.

Cohort acknowledgements:

BACCS: This work was supported in part by the NIHR Maudsley Biomedical Research Centre ('BRC') hosted at King's College London and South London and Maudsley NHS Foundation Trust, and funded by the National Institute for Health Research under its Biomedical Research Centres funding initiative. The views expressed are those of the authors and not necessarily those of the BRC, the NHS, the NIHR or the Department of Health or King's College London. We gratefully acknowledge capital equipment funding from the Maudsley Charity (Grant Reference 980) and Guy's and St Thomas's Charity (Grant Reference STR130505).

BACCS-Canada: This work was supported through funding from the Canadian Institutes of Health Research, MOP-172013, to JBV at the Centre for Addiction & Mental Health, Toronto. The ascertainment of the case control cohorts was also supported by funding from GlaxoSmithKline to JBV.

BD_TRS: This work was funded by the German Research Foundation (DFG, grant FOR2107 DA1151/5-1 to UD; SFB-TRR58, Project C09 to UD) and the Interdisciplinary Center for Clinical Research (IZKF) of the medical faculty of Münster (grant Dan3/012/17 to UD).

BiGS, GAIN: FJM was supported by the NIMH Intramural Research Program, NIH, DHHS. BOMA-Australia: This work was supported by the Australian National Health and Medical Research Council, grant numbers: 1037196 (PBM, PRS), 1066177 (JMF, JIN), 1063960 (JMF, PRS); and the Lansdowne Foundation. BOMA-Germany I, BOMA-Germany II, BOMA-Germany III, PsyCourse: This work was supported by the German Ministry for Education and Research (BMBF) through the Integrated Network IntegraMent (Integrated Understanding of Causes and Mechanisms in Mental Disorders), under the auspices of the e:Med program (grant 01ZX1314A/01ZX1614A to MMN and SC, grant 01ZX1314G/01ZX1614G to MR, grant 01ZX1314K to TGS). This work was supported by the German Ministry for Education and Research (BMBF) grants NGFNplus MooDS (Systematic Investigation of the Molecular Causes of Major Mood Disorders and Schizophrenia; grant 01GS08144 to MMN and SC, grant 01GS08147 to MR). This work was also supported by the Deutsche Forschungsgemeinschaft (DFG), grant NO246/10-1 to MMN (FOR 2107), grant RI 908/11-1 to MR (FOR 2107), grant WI 3429/3-1 to SHW, grants SCHU 1603/4-1, SCHU 1603/5-1 (KFO 241) and SCHU 1603/7-1 (PsyCourse) to TGS. This work was supported by the Swiss National Science Foundation (SNSF, grant 156791 to SC). MMN is supported through the Excellence Cluster ImmunoSensation. TGS is supported by an unrestricted grant from the Dr. Lisa-Oehler Foundation. AJF received support from the BONFOR Programme of the University of Bonn, Germany. MH was supported by the Deutsche Forschungsgemeinschaft.

Fran (France): This research was supported by Assistance Publique des Hôpitaux de Paris (APHP Grant PHRC GAN12), by Institut National de la Santé et de la Recherche Médicale (INSERM grant C0829), by the Fondation FondaMental and by the Investissements d'Avenir Programs managed by the Agence nationale pour la Recherche (references ANR-11-IDEX-0004-02 and ANR-10-COHO-10-01).

Halifax: Halifax data were obtained with support from the Canadian Institutes of Health Research (grant #166098), Genome Canada and from Dalhousie Medical Research Foundation.

HUNT: The Nord-Trøndelag Health Study (The HUNT Study) is a collaboration between HUNT Research Centre (Faculty of Medicine and Health Sciences, NTNU, Norwegian University of Science and Technology), Trøndelag County Council, Central Norway Regional Health Authority, and the Norwegian Institute of Public Health. The genotyping was financed by the National Institute of health (NIH), University of Michigan, The Norwegian Research council, and Central Norway Regional Health Authority and the Faculty of Medicine and Health Sciences, Norwegian University of Science and Technology (NTNU). The genotype quality control and imputation has been conducted by the K.G. Jebsen center for genetic epidemiology, Department of public health and nursing, Faculty of medicine and health sciences, Norwegian University of Science and Technology (NTNU).

iPSYCH BP group: ADB and the iPSYCH team acknowledges funding from The Lundbeck Foundation (grant no R102-A9118, R155-2014-1724 and R248-2017-2003), the Stanley Medical Research Institute, NIMH (1U01MH109514-01), an Advanced Grant from the European Research Council (project no: 294838), and grants from Aarhus University to the iSEQ, CGPM (GenomeDK HPC facility) and CIRRAU centers. The Danish National Biobank resource was supported by the Novo Nordisk Foundation.

The Mayo Bipolar Disorder Biobank was funded by the Marriot Foundation and the Mayo Clinic Center for Individualized Medicine.

Michigan (NIMH/Pritzker Neuropsychiatric Disorders Research Consortium): We thank the participants who donated their time and DNA to make this study possible. We thank members of the NIMH Human Genetics Initiative and the University of Michigan Prechter Bipolar DNA Repository for generously providing phenotype data and DNA samples. Many of the authors are members of the Pritzker Neuropsychiatric Disorders Research Consortium which is supported by the Pritzker Neuropsychiatric Disorders Research Fund L.L.C. A shared intellectual property agreement exists between this philanthropic fund and the University of Michigan, Stanford University, the Weill Medical College of Cornell University, HudsonAlpha Institute of Biotechnology, the Universities of California at Davis, and at Irvine, to encourage the development of appropriate findings for research and clinical applications.

Neuc1 (NeuRA-CASSI-Australia): This work was funded by the NSW Ministry of Health, Office of Health and Medical Research. CSW was a recipient of National Health and Medical Research Council (Australia) Fellowships (#1117079, #1021970).

Neuc1 (NeuRA-IGP-Australia): MJG was supported by a NHMRC Career Development Fellowship (1061875).

Neuc1 (NeuRA-BDHR-Australia): This work was supported by the Australian National Health and Medical Research Council, grant numbers: 1037196 (PBM, PRS), 1066177 (JMF, JIN), 1063960 (JMF, PRS); and the Lansdowne Foundation. JMF would like to thank Janette M O'Neil and Betty C Lynch for their support.

Neuc1/ASGC1/ASRB: This study used samples and data from the Australian Schizophrenia Research Bank (ASRB), which is supported by the National Health and Medical Research Council of Australia, the Pratt Foundation, Ramsay Health Care, the Viertel Charitable Foundation and the Schizophrenia Research Institute. We thank and acknowledge the contribution of the ASRB Chief Investigators: V. Carr, U. Schall, R. Scott, A. Jablensky, B. Mowry, P. Michie, S. Catts, F. Henskens and C. Pantelis.

Norway: TE was funded by The South-East Norway Regional Health Authority (#2015-078) and a research grant from Mrs. Throne-Holst.

Span2: CSM was a recipient of a Sara Borrell contract (CD15/00199) and a mobility grant (MV16/00039) from the Instituto de Salud Carlos III, Ministerio de Economía, Industria y Competitividad, Spain. MR was a recipient of a Miguel de Servet contract (CP09/00119 and CPII15/00023) from the Instituto de Salud Carlos III, Ministerio de Economía, Industria y Competitividad, Spain. This investigation was supported by Instituto de Salud Carlos III (PI14/01700, PI15/01789, PI16/01505,PI17/00289, PI18/01788, PI19/00721, and P19/01224), and cofinanced by the European Regional Development Fund (ERDF), "la Marató de TV3" (092330/31), the Agència de Gestió d'Ajuts Universitaris i de Recerca-AGAUR, Generalitat de Catalunya (2014SGR1357 and 2017SGR1461) and the Pla estratègic de recerca i innovació en salut (PERIS), Generalitat de Catalunya (MENTAL-Cat; SLT006/17/287). This project has also received funding from the European Union's Horizon 2020 Research and Innovation Programme under the grant agreements No 667302 (CoCA) and 728018 (Eat2beNICE).

SWEBIC: We are deeply grateful for the participation of all subjects contributing to this research, and to the collection team that worked to recruit them. We also wish to thank the Swedish National Quality Register for Bipolar Disorders: BipoläR. Funding support was provided by the Stanley Center for Psychiatric Research, Broad Institute from a grant from Stanley Medical Research Institute, the Swedish Research Council, and the NIMH.

Sweden: This work was funded by the Swedish Research Council (M. Schalling, C. Lavebratt), the Stockholm County Council (M. Schalling, C. Lavebratt, L. Backlund, L. Frisén, U. Ösby) and the Söderström Foundation (L. Backlund) and the Swedish Brain Foundation (T. Olsson).

UK - BDRN: BDRN would like to acknowledge funding from the Wellcome Trust and Stanley Medical Research Institute, and especially the research participants who continue to give their time to participate in our research.

UNIBO / University of Barcelona, Hospital Clinic, IDIBAPS, CIBERSAM: EV thanks the support of the Spanish Ministry of Economy and Competitiveness (PI15/00283) integrated into the Plan Nacional de

I+D+I y cofinanciado por el ISCIII-Subdirección General de Evaluación y el Fondo Europeo de Desarrollo Regional (FEDER); CIBERSAM; and the Comissionat per a Universitats i Recerca del DIUE de la Generalitat de Catalunya to the Bipolar Disorders Group (2014 SGR 398).

WTCCC: The principal funder of this project was the Wellcome Trust. For the 1958 Birth Cohort, venous blood collection was funded by the UK Medical Research Council.

EstBB: This study has been supported by grants from the Estonian Research Council (grant numbers PRG184, PRG687, IUT20-60 and IUT24-6), and the CoMorMent project within the EU H2020 Framework Program (agreement no. 847776).

This work was funded in part by a NARSAD Young Investigator award to EAS. AHY is funded by the National Institute for Health Research (NIHR) Biomedical Research Centre at South London and Maudsley NHS Foundation Trust and King's College London. The views expressed are those of the authors and not necessarily those of the NHS, the NIHR, or the Department of Health.

This work was supported in part by the Million Veteran Program Merit Review Award #I01 BX003341 from the U.S. Department of Veterans Affairs, Biomedical Laboratory Research and Development Service and the VISN 4 Mental Illness Research, Education and Clinical Center. The views expressed in this article are those of the authors and do not necessarily represent the position or policy of the Department of Veterans Affairs or the U.S. Government.

**Funding Sources**

| Study | Lead investigator | Country, Funder, Award number |
|---|---|---|
| PGC | P Sullivan; EA Stahl | USA, NIMH MH109528; NIMH U01 MH109536 |
| PGC | D Posthuma | Netherlands, Scientific Organization Netherlands, 480-05-003 |
| PGC | D Posthuma | Dutch Brain Foundation and the VU University Amsterdam Netherlands |
| UK - BDRN (Cardiff) | PA Holmans | Medical Research Council (MRC) Centre (G0801418) and Program Grants (G0800509) |
| Analysis | NR Wray | NHMRC 1078901,108788 |
| ASRB | V Carr | Australia, National Health and Medical Research Council, grant number: (86500). |

| | | |
|---|---|---|
| ASRB | M Cairns | Australia, National Health and Medical Research Council, grant numbers: (1121474, 1147644). |
| ASRB | C Pantelis | Australia, National Health and Medical Research Council, grant numbers: (628386, 1105825). |
| BACCS | G Breen | GB, JRIC, HG, CL were supported in part by the NIHR Maudsley Biomedical Research Centre (‘BRC’) hosted at King’s College London and South London and Maudsley NHS Foundation Trust, and funded by the National Institute for Health Research under its Biomedical Research Centres funding initiative. |
| BD_TRS | U Dannlowski | Germany, DFG, Grant FOR2107 DA1151/5-1; Grant SFB-TRR58, Project C09 |
| BiGS, Uchicago | ES Gershon | R01 MH103368 |
| BiGS, NIMH | FJ McMahon | US, NIMH, R01 MH061613, ZIA MH002843 |
| BiGS, GAIN, UCSD | J Kelsoe | US, NIMH, MH078151, MH081804, MH59567 |
| BOMA-Australia | JM Fullerton | Australia, National Health and Medical Research Council, grant numbers: 1066177; 1063960 |
| BOMA-Australia | SE Medland | Australia, National Health and Medical Research Council, grant numbers: 1103623 |
| BOMA-Australia | PB Mitchell | Australia, National Health and Medical Research Council, grant numbers: 1037196 |
| BOMA-Australia | GW Montgomery | Australia, National Health and Medical Research Council, grant numbers: 1078399 |
| BOMA-Australia | PR Schofield | Australia, National Health and Medical Research Council, grant numbers: 1037196; 1063960 |
| Romania (BOMA-Romania) bip_rom3_eur and bmrom | M Grigoroiu-Serbanescu | Multiple grants from UEFISCDI, Romania |

| | | |
|---|---|---|
| BOMA-Germany I, II, III | S Cichon | Germany, BMBF Integrament, 01ZX1314A/01ZX1614A |
| BOMA-Germany I, II, III | S Cichon | Germany, BMBF NGFNplus MooDS, 01GS08144 |
| BOMA-Germany I, II, III | S Cichon | Switzerland, SNSF, 156791 |
| BOMA-Germany I, II, III | MM Nöthen | Germany, BMBF Integrament, 01ZX1314A/01ZX1614A |
| BOMA-Germany I, II, III | MM Nöthen | Germany, BMBF NGFNplus MooDS, 01GS08144 |
| BOMA-Germany I, II, III | MM Nöthen | Germany, Deutsche Forschungsgemeinschaft, Excellence Cluster ImmunoSensation |
| BOMA-Germany I, II, III | MM Nöthen | Germany, Deutsche Forschungsgemeinschaft, NO246/10-1 |
| BOMA-Germany I, II, III | SH Witt | Germany, Deutsche Forschungsgemeinschaft, WI 3429/3-2 |
| BOMA-Germany I, II, III, BOMA-Spain | M Rietschel | Germany, Deutsche Forschungsgemeinschaft, RI 908/11-1 |
| BOMA-Germany I, II, III, BOMA-Spain | M Rietschel | Germany, BMBF, ERA-Net Neuron “EMBED”, 01EW1904 |
| BOMA-Germany I, II, III, BOMA-Spain | M Rietschel | Germany, BMBF, ERA-Net Neuron “Synschiz”, 01EW1810 |
| BOMA-Germany I, II, III, PsyCourse, BiGS | TG Schulze | Germany, BMBF Integrament, 01ZX1314K |
| BOMA-Germany I, II, III, PsyCourse, BiGS | TG Schulze | Germany, DFG, SCHU 1603/4-1, SCHU 1603/5-1, SCHU 1603/7-1 |
| BOMA-Germany I, II, III, PsyCourse, BiGS | TG Schulze | Germany, Dr. Lisa-Oehler Foundation (Kassel, Germany) |

| | | |
|---|---|---|
| Bulgarian Trios (Cardiff) | G Kirov | The recruitment was funded by the Janssen Research Foundation. Genotyping was funded by multiple grants to the Stanley Center for Psychiatric Research at the Broad Institute from the Stanley Medical Research Institute, The Merck Genome Research Foundation, and the Herman Foundation. |
| EstBB | L Milani | Estonia, Estonian Research Council (grant numbers PRG184, PRG687, IUT20-60 and IUT24-6), European Commission EU H2020 (CoMorMent project, agreement no. 847776). |
| France | M Leboyer, F Bellivier, B Etain, S Jamain | France, APHP, INSERM, ANR, Fondation Fondamental |
| Halifax | M Alda | CIHR grant #166098 and Dalhousie Medical Research Foundation |
| iPSYCH BP group | AD Børglum | Denmark, Lundbeck Foundation, R102-A9118 and R155-2014-1724 (iPSYCH) |
| iPSYCH BP group | AD Børglum | Denmark, Aarhus University, iSEQ and CIRRAU |
| iPSYCH BP group | AD Børglum | USA, Stanley Medical Research Institute |
| iPSYCH BP group | AD Børglum | EU, European Research Council, 294838 |
| Mayo Bipolar Disorder Biobank | JM Biernacka, MA Frye | Marriot Foundation and the Mayo Clinic Center for Individualized Medicine |
| Michigan | M Boehnke | US, NIMH, R01 MH09414501A1; US, NIMH, MH105653 |
| Million Veteran Program | H Kranzler | I01 BX003341 from the U.S. Department of Veterans Affairs, Biomedical Laboratory Research and Development Service and the VISN 4 Mental Illness Research, Education, and Clinical Center. |
| Mount Sinai | EA Stahl | NARSAD Young Investigator Award |
| Mount Sinai, STEP-BD, FAST | P Sklar, EA Stahl | US NIH R01MH106531, R01MH109536 |

| | | |
|---|---|---|
| NeuRA-CASSI-Australia (neuc1) | C Shannon Weickert | Australia, National Health and Medical Research Council, grant number: 568807 |
| NeuRA-CASSI-Australia (neuc1) | TW Weickert | Australia, National Health and Medical Research Council, grant number: 568807 |
| NeuRA-IGP-Australia (neuc1) | MJ Green | Australia, National Health and Medical Research Council, grant numbers: 630471, 1081603 |
| NeuRA-BDHR-Australia (neuc1) | PB Mitchell, PR Schofield, JM Fullerton | Australia, National Health and Medical Research Council, grant numbers: 1037196; 1066177; 1063960, & The Lansdowne Foundation |
| Norway | I Agartz | Sweden, Swedish Research Council |
| Norway | OA Andreassen | Norway, Research Council of Norway (#217776, #223273, #248778, #249711), KG Jebsen Stiftelsen, The South-East Norway Regional Health Authority (#2012-132, #2012-131, #2017-004) |
| Norway | T Elvsåshagen | Norway, The South-East Norway Regional Health Authority (#2015-078) and a research grant from Mrs. Throne-Holst. |
| Norway | I Melle | Norway, Research Council of Norway (#421716,#223273), KG Jebsen Stiftelsen, The South-East Norway Regional Health Authority (#2011085, #2013088, #2014102) |
| Norway | KJ Oedegaard | Norway, the Western Norway Regional Health Authority |
| Norway | OB Smeland | Norway, The South-East Norway Regional Health Authority (#2016-064, #2017-004) |
| Span2 | M Ribasés | Spain, Instituto de Salud Carlos III, Ministerio de Economía, Industria y Competitividad, CP09/00119 and CPII15/00023 |
| Span2 | C Sánchez-Mora | Spain, Instituto de Salud Carlos III, Ministerio de Economía, Industria y Competitividad, CD15/00199 and MV16/00039 |
| State University of New York, | C Pato, MT Pato, JA Knowles, H Medeiros | US, National Institutes of Health, R01MH085542 |

| | | |
|---|---|---|
| Downstate Medical Center (SUNY DMC) | | |
| SWEBIC | M Landén | The Stanley Center for Psychiatric Research, Broad Institute from a grant from Stanley Medical Research Institute; NIMH MH077139 (PFS), The Swedish Research Council: 2018-02653 (ML), the Swedish foundation for Strategic Research (KF10-0039), the Brain foundation (FO2020-0261), and the Swedish Federal Government under the LUA/ALF agreement (ALF 20170019, ALFGBG-716801). |
| UCL | A McQuillin | Medical Research Council (MRC) - G1000708 |
| UCLA-Utrecht (Los Angeles) | NB Freimer | US, National Institutes of Health, R01MH090553, U01MH105578 |
| UCLA-Utrecht (Los Angeles) | LM Olde Loohuis | US, National Institutes of Health, R01MH090553, U01MH105578 |
| UCLA-Utrecht (Los Angeles) | RA Ophoff | US, National Institutes of Health, R01MH090553, U01MH105578 |
| UCLA-Utrecht (Los Angeles) | APS Ori | US, National Institutes of Health, R01MH090553, U01MH105578 |
| UK - BDRN (Cardiff) | MC O'Donovan | Medical Research Council (MRC) Centre (G0801418) and Program Grants (G0800509) |
| UK - BDRN (Cardiff) | MJ Owen | Medical Research Council (MRC) Centre (G0801418) and Program Grants (G0800509) |
| UK - BDRN (Cardiff) | N Craddock, I Jones, LA Jones | UK, Wellcome Trust, 078901; USA, Stanley Medical Research Institute, 5710002223-01 |
| UK - BDRN (Cardiff) | A Di Florio | European Commission Marie Curie Fellowship, grant number 623932. |
| UNIBO / University of Barcelona, Hospital Clinic, IDIBAPS, CIBERSAM | E Vieta | Grants PI15/00283 (Spain) and 2014 SGR 398 (Catalonia) |

| | | |
|---|---|---|
| University of Pittsburgh | V Nimgaonkar | US, NIMH MH63480 |
| USC | JL Sobell | USA, National Institutes of Health, R01MH085542 |
| WTCCC | N Craddock; AH Young | Wellcome Trust. For the 1958 Birth Cohort, venous blood collection was funded by the UK Medical Research Council. AHY was funded by NIMH (USA); CIHR (Canada); NARSAD (USA); Stanley Medical Research Institute (USA); MRC (UK); Wellcome Trust (UK); Royal College of Physicians (Edin); BMA (UK); UBC-VGH Foundation (Canada); WEDC (Canada); CCS Depression Research Fund (Canada); MSFHR (Canada); NIHR (UK); Janssen (UK) |
| Greece | G.P.Patrinos | European Commission (H2020-668353; Ubiquitous Pharmacogenomics (U-PGx); Greek General Secretariat of Research and Technology (MIS 5002550) |
| University of California, Irvine | MP Vawter | USA, National Institutes of Health, MH113177, MH074307, MH085801, MH099440, RR000827, MH060068, MH060870.<br>Pritzker Neuropsychiatric Disorders Research Fund L.L.C. |
| Korea | HJ Lee | National Research Foundation of Korea (2019R1A2C2084158, 2017M3A9F1031220). Ministry of Health & Welfare of Korea (HM14C2606) |
| BIPGRAZ, Austria | EZ Reininghaus | Funding from the government of Styria, Austria. |
| BIPGRAZ, Austria | SA Bengesser | MEFO funding of the Medical University of Graz. |
| Korea | KS Hong | National Research Foundation of Korea (2019R1A2C1005346, 2019M3C7A1030617) |
| Japan | N Iwata, M Ikeda | Japan Agency for Medical Research and Development (JP20dm0107097, JP20km0405201, JP20km0405208) |

**Consortium authors and affiliations**

Named authors and their affiliations appear in the main manuscript.

*HUNT All-In Psychiatry*
Amy Martinsen[1,2,14], Anne Heidi Skogholt[2,3], Bendik S Winsvold[2,4], Børge Sivertsen[5,6,7], Cristen Willer[8,9,10,] Eystein Stordal[6,11], Grete Dyb[12], Gunnar Morken[6,13], Håvard Kallestad[6,13], Ingrid Heuch[4], John-Anker Zwart[2,4,14], Jonas Bille Nielsen[8,] Katrine Kveli Fjukstad[15,16], Lars Fritsche[8,] Laurent Thomas[2], Linda M Pedersen[4], Maiken Elvestad Gabrielsen[2], Marianne Bakke Johnsen[1,14], Marit Skrove[17], Marit Sæbø Indredavik[3], Ole Kristian Drange[6,13], Ottar Bjerkeset[6,18], Sigrid Børte[1,2,14], Synne Øien Stensland[1,12], Torunn Stene Nøvik[19], Wei Zhou[9,10]

1 Research and Communication Unit for Musculoskeletal Health (FORMI), Department of Research, Innovation and Education, Division of Clinical Neuroscience, Oslo University Hospital, Oslo, Norway.
2 K. G. Jebsen Center for Genetic Epidemiology, Department of Public Health and Nursing, Faculty of Medicine and Health Sciences, Norwegian University of Science and Technology, Trondheim, Norway.
3 Department of Clinical and Molecular Medicine, Norwegian University of Science and Technology, Trondheim, Norway.
4 Department of Research, Innovation and Education, Division of Clinical Neuroscience, Oslo University Hospital, Oslo, Norway.
5 Department of Health Promotion, Norwegian Institute of Public Health, Bergen, Norway.
6 Department of Mental Health, Faculty of Medicine and Health Sciences, Norwegian University of Science and Technology, Trondheim, Norway.
7 Department of Research and Innovation, Helse-Fonna HF, Haugesund, Norway.
8 Department of Internal Medicine, Division of Cardiovascular Medicine, University of Michigan, Ann Arbor, 48109, MI, USA.
9 Department of Computational Medicine and Bioinformatics, University of Michigan, Ann Arbor, MI, USA.
10 Department of Human Genetics, University of Michigan, Ann Arbor, MI 48109, USA; Center for Statistical Genetics, University of Michigan, Ann Arbor, MI 48109, USA.
11 Department of Psychiatry, Hospital Namsos, Nord-Trøndelag Health Trust, Namsos, Norway.
12 NKVTS, Norwegian Centre for Violence and Traumatic Stress Studies.
13 Division of Mental Health Care, St. Olavs Hospital, Trondheim University Hospital, Trondheim, Norway.
14 Institute of Clinical Medicine, Faculty of Medicine, University of Oslo, Oslo, Norway.
15 Department of Psychiatry, Nord-Trøndelag Hospital Trust, Levanger Hospital, Norway.
16 Department of Laboratory Medicine, Children’s and Women’s Health, Norwegian University of Science and Technology, Trondheim, Norway.
17 Regional Centre for Child and Youth Mental Health and Child Welfare, Department of Mental Health, Faculty of Medicine and Health Sciences, NTNU – Norwegian University of Science and Technology.
18 Faculty of Nursing and Health Sciences, NORD University, Levanger, Norway.
19 Department of Child and Adolescent Psychiatry, St. Olavs Hospital, Trondheim University Hospital, Trondheim, Norway.

102. American Psychiatric Association. *Diagnostic and Statistical Manual of Mental Disorders 5th edn*.

(American Psychiatric Association Publishing, 2013).

113. Bergink, V., Rasgon, N. & Wisner, K. L. Postpartum Psychosis: Madness, Mania, and Melancholia in

Motherhood. *Am. J. Psychiatry* **173**, 1179–1188 (2016).

143. National Institute for Health and Care Excellence. Bipolar disorder: assessment and management. https://www.nice.org.uk/guidance/cg185/chapter/1-R.
